# Polygenic Risk-Informed White Matter Integrity Improves Deep Learning-Based Prediction of Youth Depression

**DOI:** 10.1101/2025.03.27.25324746

**Authors:** Jungyoun Janice Min, Heehwan Wang, Eunji Lee, Bo-Gyeom Kim, Gakyung Kim, Seung Yun Choi, Kahyun Choi, Sung Hee Hong, Yumin Jang, Yu Jin Rah, Ji Yeon Kim, Seunghui Han, Kyung Hwa Lee, Junhyung Kim, Milenna T. van Dijk, Jae-Won Kim, Grace H. Chung, Dongil Chung, Sang Ah Lee, Yoonjung Yoonie Joo, Jiook Cha

## Abstract

Early detection of youth depression is crucial, given its rising prevalence and long-term consequences. Although genetic factors contribute significantly to youth depression, their integration with neuroimaging remains limited. We present a deep learning framework using polygenic scores (PGS) to pretrain a 3D convolutional neural network on diffusion MRI (track-weighted fractional anisotropy), capturing gene–brain associations from a multi-ethnic cohort in the Adolescent Brain Cognitive Development Study (N=4,741). Fine-tuned for predicting depression, the model improved cross-sectional (N=266; AUC=0.62) and two-year predictions of depression and suicidality (AUC=0.61–0.66). It outperformed unimodal models, increasing accuracy by up to 24% over PGS-only and 5.8% over brain-only models. Explainable artificial intelligence identified key white matter tracts—superior longitudinal fasciculus, cingulum and corpus callosum—as predictive features. Decision curve analysis showed greater clinical utility. The model generalized to an independent Korean youth sample (N=108; AUC=0.67), supporting the cross-ethnic scalability of PGS-informed diffusion MRI for precision psychiatry

## Introduction

Youth depression is a growing global health concern [1–3], marked by early onset, high recurrence, resistance to treatments, and long-term consequences including suicide, which is a leading cause of death of youth [4–9]. These features underscore the urgent need for early identification to alter adverse developmental trajectories. Within the framework of precision psychiatry, which emphasizes individualized risk detection, biological markers reflecting latent vulnerability—such as genetic predispositions and brain-based features—represent a promising avenue for early prediction and intervention.

Youth depression exhibits stronger genetic associations than adult-onset depression [10], including higher polygenic risk scores (PGS) and increased genetic overlap with other psychiatric disorders [11–13]. Twin studies confirm high heritability [14, 15] with genetic factors impacting symptom persistence and stability [10, 16, 17]. Youths with high familial risk are more likely to experience early-onset depression, further shaped by stress and maternal psychopathology [18]. These genetic vulnerabilities interact strongly with neurobiological substrates, particularly white matter integrity, which provides the structural basis for long-range neural connectivity [19–21]. Fractional Anisotropy (FA), derived from diffusion MRI (dMRI), serves as a key marker of white matter integrity [22]. It demonstrates high heritability with genetic factors explaining up to 88% of its variance [19, 23, 24]. Higher polygenic risk for major depressive disorder (MDD) has been linked to reductions in white mater integrity, suggesting that genetic predisposition may contribute to neurostructural vulnerabilities in depression [25]. Specific tracts, such as the corpus callosum, corona radiata, and superior longitudinal fasciculus, are strongly influenced by genetic factors, supporting the notion that white matter differences are under strong genetic control [21, 24].

Converging evidence from human and animal studies implicates disruptions in specific white matter tracts in the pathophysiology of depression. Human studies report reduced FA in major white matter tracts, including the corpus callosum, cingulum, superior longitudinal fasciculus, and uncinate fasciculus [26–32]. Consistent with these findings, animal models of depression show reduced FA in the corpus callosum and related tracts [33, 34]. Developmental alterations in white matter, particularly in the corpus callosum, fimbria, external capsule, and internal capsule, were observed in rodent model of depression [35]. These findings collectively underscore the critical role of changes in white matter integrity in the pathophysiology of depression.

The interplay between genetic predispositions and white matter integrity is critical for understanding the biological basis of depression. Supporting this, studies have identified significant genetic correlations between measures of diffusion tensor imaging and depression-related traits [23, 36]. Additionally, forebrain white matter-related dMRI measures have been associated with single nucleotide polymorphisms in genes, encoding proteins involved in extracellular matrix and epidermal growth factor signaling, which are critical for myelin repair and synaptic plasticity and closely linked to MDD [20]. These findings suggest shared genetic mechanisms influencing both white matter integrity and depression susceptibility.

Despite advancements, critical gaps remain in our understanding of youth depression. Most studies have examined genetic and neuroimaging factors in isolation, failing to capture their complex interplay [32, 37–39]. Research in other psychiatric disorders shows that integrating genetic and neuroimaging data through multi-modal approaches enhances predictive accuracy over single-modality models [40–42]. Deep learning methodologies enables multi-modal integration [43], by capturing complex non-linear relationships and learning shared representations across different modalities. However, their application in developmental psychiatric research is often constrained by small sample sizes, limiting model generalizability. [44, 45]. Optimized strategies are therefore needed to effectively utilize multi-modal information in youth depression research, where datasets are typically smaller.

Additionally, most existing studies in both neuroimaging and genetics have been conducted on predominantly Caucasian populations, leaving other ethnic groups underrepresented [46, 47]. Addressing this disparity is critical to developing globally applicable predictive models for depression. Further investigation is needed to assess model applicability in underrepresented groups, such as through transfer learning approaches.

To address these challenges, this study introduces a novel deep learning framework that leverages PGS to *pretrain* a model on dMRI data (specifically, track-weighted fractional anisotropy or TW-FA) from a large, multi-ethnic cohort (the Adolescent Brain Cognitive Development [ABCD] Study), then fine-tune for classification of depression. This approach aims to: (1) improve the accuracy of *both* cross-sectional and longitudinal prediction of youth depression by integrating genetic and neuroimaging information; (2) identify key white matter tracts associated with depression risk; and (3) demonstrate the *cross-ethnic generalizability* of our model by validating it in an independent Korean cohort.

## Results

We analyzed 5,248 ABCD participants with multiple ancestral backgrounds meeting strict quality criteria for dMRI and genetic data. The dataset was divided into three groups: Pretrain (N=4,741), Finetune (N=266), and 2-Year Follow-up Prediction (N=236) sets with additional subsets for suicidality analyses (Figure. S1). We pretrained a 3DCNN on depression PGS and composite PGS alongside white matter imaging to learn gene-brain associations using the pretraining set, then fine-tuned for depression classification via 10-fold cross-validation (Figure. 1). Propensity score matching (age, sex, and study site) minimized selection bias in each fold. To ensure robust evaluation of zero-shot prediction in the 2-Year Prediction subsets, we generated 10 matched subsets with diverse controls. Further details on PGS calculations are provided in the Methods section. Full demographics and performance metrics are in Supplementary Materials (Table. S1, S12–S33).

**Figure 1.**
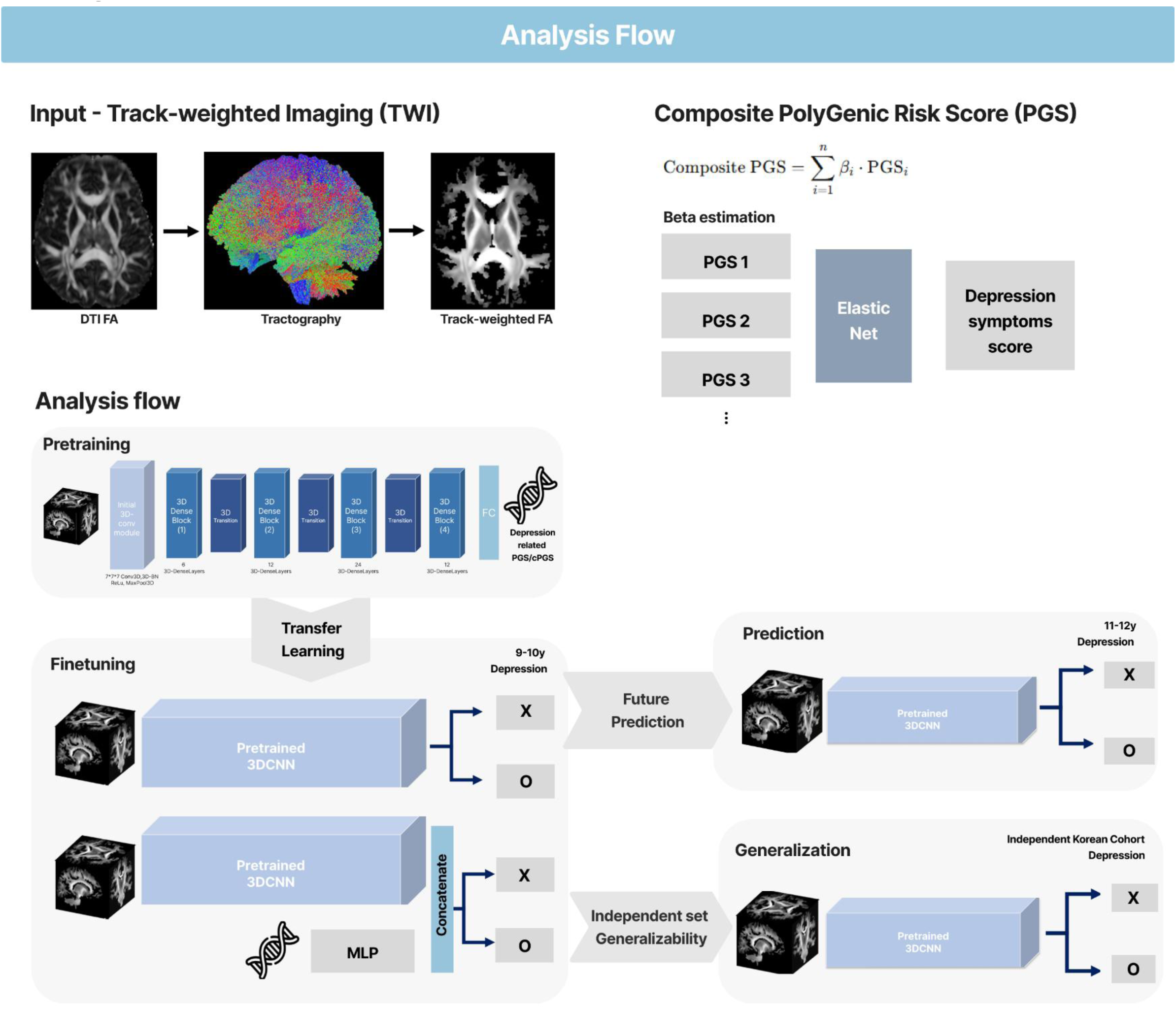
Analysis Flows. This study integrates Track-Weighted FA and PGS to enhance youth depression prediction. The model is pretrained on white matter imaging and depression-related PGS using a 3D convolutional neural network. Fine-tuned model evaluates 9-10y depression classification, 2-year follow-up depression prediction followed by validation in an independent Korean cohort for generalizability (Multilayer perceptron layer (MLP)).

### PGS-Pretrained 3DCNN Outperforms Comparison Models in Cross-sectional MDD classification

The PGS-pretrained 3DCNN using track-weighted FA (TW-FA) consistently outperformed unimodal and multimodal baselines for both cross-sectional and 2-year follow-up MDD prediction. For cross-sectional classification, the composite PGS-pretrained model achieved an AUROC of 0.62±0.121 and an F1 score of 0.636±0.072, surpassing models trained with only composite PGS (AUROC = 0.50±0.118, F1 = 0.444±0.149), only TW-FA (AUROC = 0.586±0.133, F1 = 0.278±0.212), and multimodal models trained without pretraining (AUROC = 0.562±0.118, F1 = 0.449±0.217) (Figure. 2A, B). Compared to composite PGS-only models, the pretrained model improved AUROC by 24%, while outperforming brain-only and non-pretrained multimodal models by 5.8% and 10.3%, respectively. F1 score differences were statistically significant across all comparisons (p < 0.05) (Table. S13).

**Figure 2.**
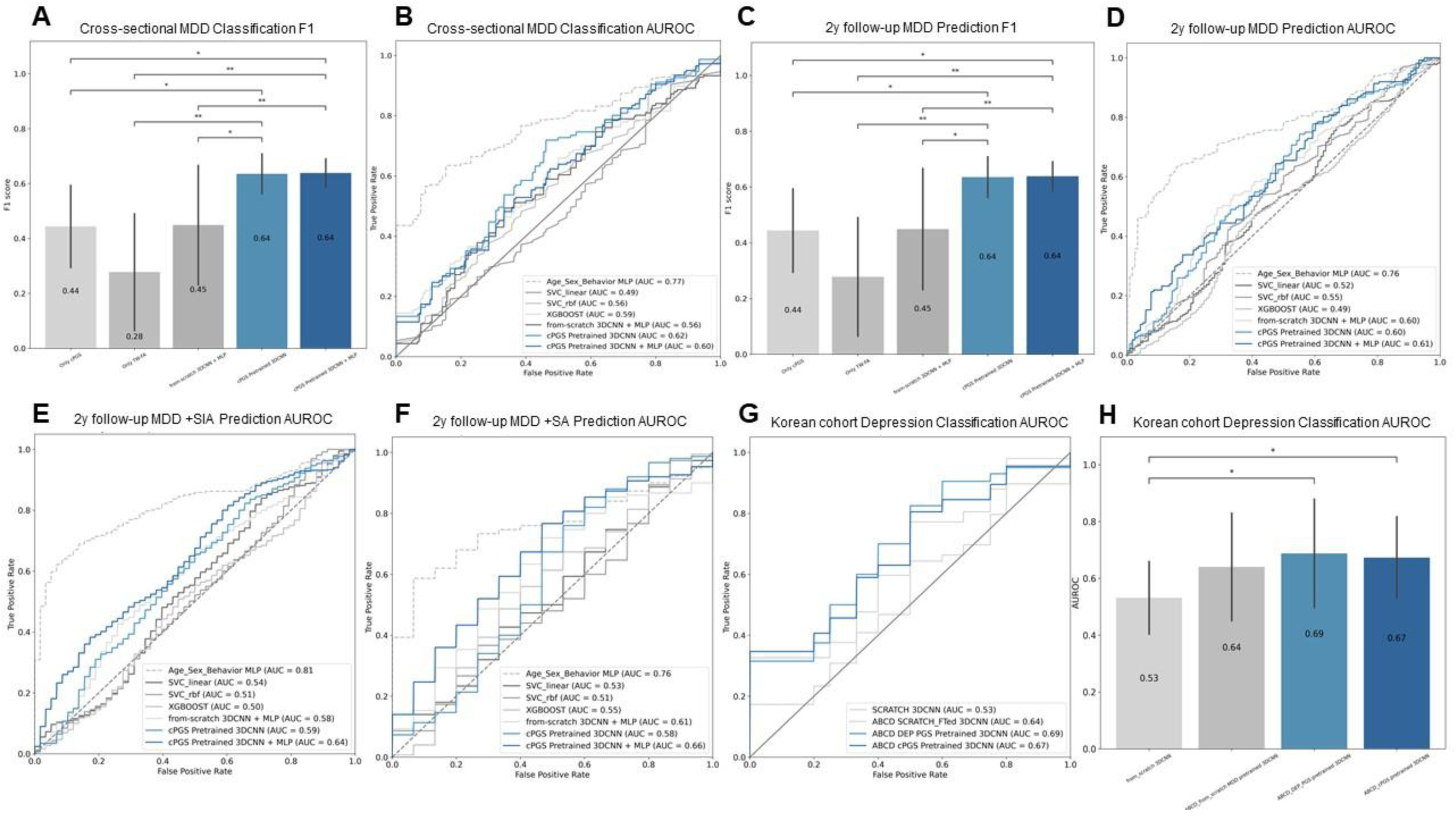
Depression Classification Task Performance using Composite PGS. (A) F1 score comparison for cross-sectional MDD classification, showing statistically significant performance differences across models. (B) Receiver operating characteristic (ROC) curves displaying AUROC values for cross-sectional MDD classification models. (C) F1 score comparison for 2-year follow-up MDD prediction, highlighting model performance differences. (D) ROC curves for 2-year follow-up MDD prediction. (E) ROC curves for 2-year follow-up MDD and suicidal active ideation prediction. (F) ROC curves for 2-year follow-up MDD and suicidal attempt prediction. (A-F) ABCD cohort. (G) ROC curves for depression classification in the independent Korean cohort, demonstrating external validation performance (H) AUROC comparison for depression classification, highlighting statistical significance in generalizability across models. (G-H) Korean cohort. (*: p-value < 0.05, **: p-value <0.01, ***p-value <0.001).

The benchmark for psychiatric risk prediction was established using symptom-based model that incorporated behavior (CBCL internalizing problems), age, and biological sex, achieving an AUROC of 0.767±0.07. Our PGS-pretrained model retained 80.5% of this benchmark performance, demonstrating that genetic and neuroimaging biomarkers can serve as robust predictive features.

A similar pattern was observed with depression PGS, where the pretrained model (AUROC = 0.597±0.12, F1 = 0.629±0.085) outperformed depression PGS-only (AUROC = 0.546±0.114, F1 = 0.517±0.155) and non-pretrained multimodal models (AUROC = 0.54±0.146, F1 = 0.484±0.223) (Figure. S2A, B; Table. S12). Additionally, the pretrained 3DCNN surpassed traditional machine learning models, including support vector classifiers and XGBOOST (Table. S3), further underscoring the effectiveness of integrating PGS-based pretraining with white matter imaging.

### PGS-Pretraining Enhances 2-Year Follow-Up MDD Prediction

In the 2-year follow-up MDD prediction, conducted as a zero-shot evaluation on unseen data, our PGS-pretrained model again demonstrated superior predictive accuracy. The 3DCNN+Multi-layer Perceptron(3DCNN+MLP) model, which processes brain images via a composite PGS–pretrained 3DCNN and integrates composite PGS through an additional MLP, achieved an AUROC of 0.61±0.011 and an F1 score of 0.652±0.007, significantly surpassing models using only composite PGS (AUROC = 0.562±0.011, F1 = 0.474±0.016), only TW-FA (AUROC = 0.589±0.012, F1 = 0.534±0.016), and from-scratch multimodal models (AUROC = 0.596±0.008, F1 = 0.604±0.011) (p < 0.05) (Figure. 2C, D; Table. S17). Specifically, compared to composite PGS-only models, our pretrained model improved AUROC by 8.5%, while outperforming brain-only and from-scratch multimodal models by 3.6% and 2.3%, respectively.

A similar trend was observed for depression PGS, where the depression PGS-pretrained 3DCNN+MLP model (AUROC = 0.619±0.014, F1 = 0.623±0.012) significantly outperformed depression PGS-only (AUROC = 0.538±0.01, F1 = 0.473±0.017) and from-scratch multimodal models (AUROC = 0.607±0.014, F1 = 0.565±0.02) (p < 0.05) (Figure. S2C, D; Table. S16).

Importantly, traditional machine learning models showed poor generalizability, with AUROC and F1 scores below 0.55 (Table. S4). In contrast, both PGS-pretrained + MLP models exhibited significant improvements over conventional machine learning models (p < 0.01) (Table. S18, S19).

These findings highlight the utility of PGS integration strategies across tasks. While PGS-based pretraining alone improved cross-sectional MDD prediction, likely by embedding stable gene–brain associations, combining pretraining with direct PGS input conferred additional advantages in 2-year follow-up prediction. This suggests that incorporating polygenic risk at both representational and decision levels may be particularly beneficial when forecasting longer-term psychiatric outcomes.

### Decision Curve Analysis (DCA) Results for Clinical Utility

To evaluate the potential clinical utility of our pretrained models, we conducted a Decision Curve Analysis (DCA) comparing the default strategies including, “Treat All” and “Treat None” with our best-performing PGS-pretrained 3DCNN. Our model yielded higher net benefit values compared to both comparison strategies across a range of clinically relevant threshold probabilities (0.4-0.5) (Figure. S3 A, B). This indicates superior performance in identifying children who would progress to MDD while minimizing unnecessary interventions.

### Leveraging PGS in 2-Year Follow-Up MDD and Suicidality Prediction

The PGS-pretrained 3DCNN demonstrated superior performance in predicting MDD with suicidality at the 2-year follow-up across all tested models. Using composite PGS, the PGS-pretrained 3DCNN + MLP model achieved the best predictive accuracy. For MDD with suicidal active ideation, our model attained an AUROC of 0.64±0.02 and F1 score of 0.675±0.007, showing a 1.9% improvement over composite PGS-only models (AUROC = 0.628±0.015) and a 13.9% improvement over unimodal TW-FA models (AUROC = 0.562±0.021) (Figure. 2E; Table. S5). While symptom-based models reached an AUROC of 0.815±0.021, our model retained 79% of this benchmark, highlighting its utility even in the absence of behavioral indicators. For MDD with suicidal attempts, the PGS-pretrained model achieved an AUROC of 0.665±0.071 (F1 = 0.702±0.034), improving upon from-scratch 3DCNN + MLP models by 9% (AUROC = 0.611±0.061) (Figure. 2F; Table. S7). Compared to the symptom-based model (AUROC = 0.756±0.03), our model retained 88% of its predictive accuracy, further supporting its clinical relevance.

A similar pattern was observed with depression PGS, where the best-performing model using TW-FA and depression PGS reached an AUROC of 0.651±0.021 (F1 = 0.664±0.008), exceeding depression PGS-only (AUROC = 0.614±0.016) by 6% and TW-FA-only models (AUROC = 0.562±0.021) by 15.8% (Figure. S2 E, F; Table. S5). For MDD with suicidal attempts, the depression PGS-pretrained model (AUROC = 0.67±0.076, F1 = 0.651±0.034) outperformed from-scratch multimodal models (AUROC = 0.661±0.074) by 7.4% and TW-FA-only models (AUROC = 0.573±0.062) by 16.9% (Figure. S2 G, H; Table. S7). Across all comparisons, both PGS-pretrained models significantly outperformed alternative approaches in F1 scores (p < 0.01) (Table. S20, S21).

Traditional machine learning models failed to generalize in the unseen dataset for MDD and suicidality prediction, with AUROC and F1 scores below 0.55 (Table. S6, S8). In contrast, PGS-pretrained + MLP models exhibited statistically significant improvements over conventional machine learning models (p < 0.05) (Table. S22, S23, S26, S27). Notably, our approach maintained robust performance across all comparison models in predicting MDD onset at the 2-year follow-up, even when excluding cases with suicidality.

We conducted a control analysis to evaluate the specificity of PGS-based pretraining. We compared our approach against alternative pretraining approaches using family history of depression, cognitive variable (NIH Toolbox), and behavioral measure (CBCL Total Problems) as pretraining outcomes. The composite PGS-pretrained model showed significantly higher AUROC than the family history and cognitive variable models (p<0.05) (Table. S28), while no statistically significant difference was observed compared to the behavioral variable-based pretraining. Additional control analyses, including microstructural versus macrostructural feature evaluations, MDD subgroup analyses and performance assessments of multi-modal models using TW-FA, PGS, age, sex, and behavioral variables are provided in the Supplementary Materials (Figure. S4-S6; Table. S9-S11).

### Machine Learning Interpretation

Using Integrated Gradients [48] and SmoothGrad [49], we generated voxel-level saliency maps to identify key white matter features associated with MDD (Figure. 3A). Lower fractional anisotropy (FA) across widespread white matter regions is associated with a higher probability of MDD. To improve interpretability, we performed fasciculus-level analyses, revealing that the superior longitudinal fasciculus, cingulum, corticospinal tract, and corpus callosum contained a high proportion of voxels with reduced FA, significantly differing from controls (Figure. 3B). In contrast, relatively few voxels showed a positive association with MDD (Figure. 3C). Children who developed depressive symptoms and suicidal ideation two years later exhibited similar FA reductions in these tracts, which strongly contributed to their symptom prediction (Figure. 3D–F).

**Figure 3.**
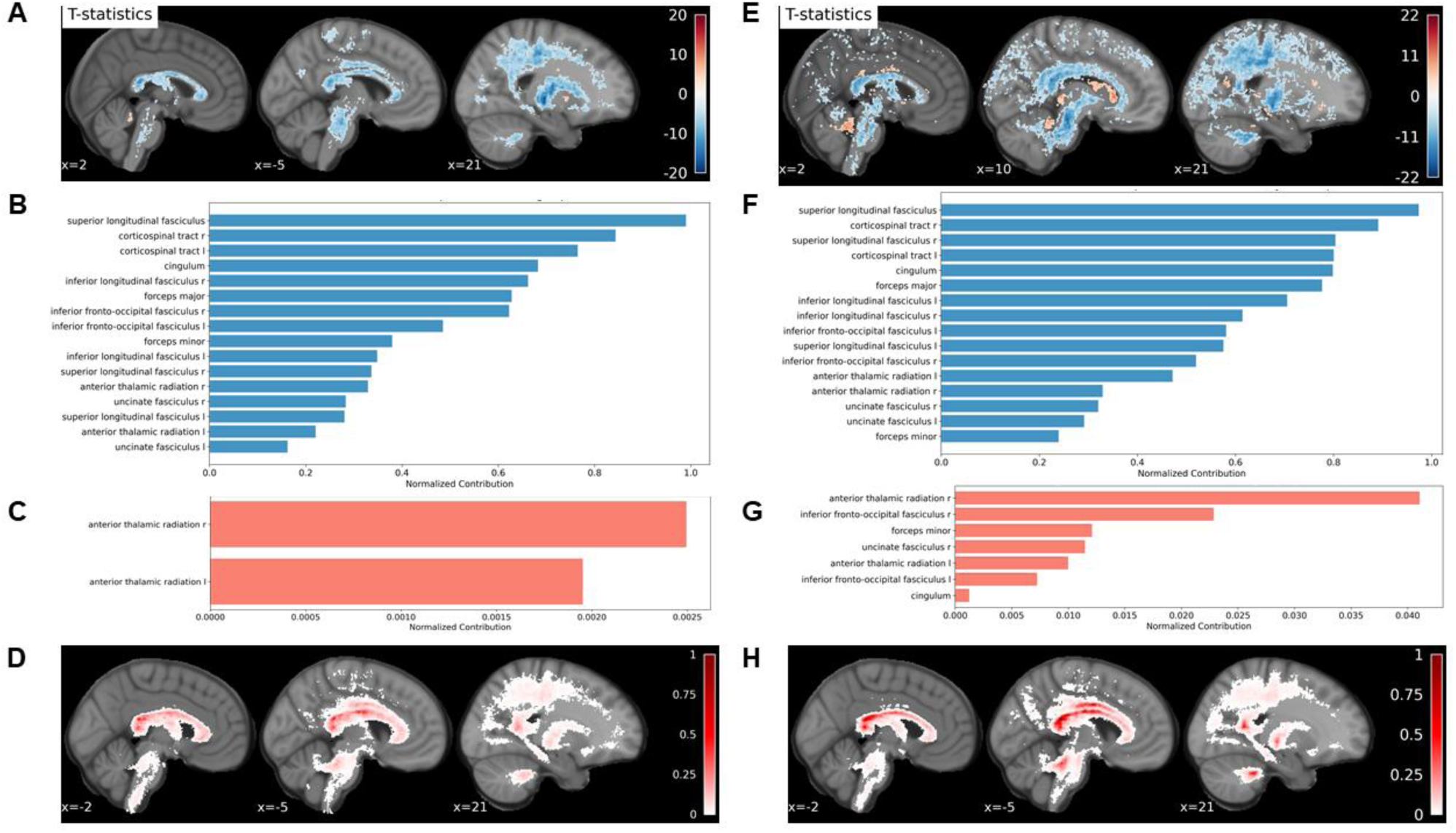
Machine Learning Interpretations Reveal Reduced White Matter FA Associated with Youth MDD in ABCD Cohort. (A) Cross-sectional (9-10y) MDD classification explainable AI (XAI) map. Voxel-wise t-tests were performed with FDR correction (*p* < 0.001, cluster threshold = 100). Blue regions indicate voxels where lower fractional anisotropy (FA) values are associated with a higher probability of MDD. (B) Fasciculus-level contributions of voxels with significantly lower FA in the MDD group. Normalized contribution was calculated as the proportion of significant voxels (negative associations) within each fasciculus. (C) Fasciculus-level contributions of voxels where higher FA is associated with a higher probability of MDD, using the same normalization method as in (B). (D) Variability map of individual attribute score within True Positive participants. (E) XAI map for MDD prediction at 2-year follow-up in individuals with suicidal active ideation. Voxel-wise t-tests with FDR correction (*p* < 0.001, cluster threshold = 100) were applied. (F) Fasciculus-level contributions of voxels where lower FA is associated with a higher probability of MDD+SIA at 2-year follow-up, following the same analysis as (B). (G) Fasciculus-level contributions of voxels where higher FA is associated with a higher probability of MDD+SIA at 2-year follow-up, following the same analysis as (C). (H) Variability map of individual attribute score within True Positive participants.

### Generalizability testing on Independent Korean Cohorts

To assess cross-ethnic generalizability, we evaluated the PGS-pretrained 3DCNN on an independent Korean adolescent sample. Fine-tuning the PGS-pretrained 3DCNN models on this dataset significantly outperformed the from-scratch model trained solely on Korean data in AUROC (p-value < 0.05) (Table. S33). The composite PGS-pretrained model achieved an AUROC of 0.673±0.154 and F1 score of 0.61±0.227, compared to the from-scratch model’s AUROC of 0.532±0.127 and F1 score of 0.5±0.215 (Figure. 2G, H; Table. 3). These observations underscore the value of combining PGS with neuroimaging data to improve cross-population applicability.

## Discussion

This study demonstrates that integrating PGS with white matter neuroimaging significantly enhances the prediction of youth depression. Using a deep learning framework, we achieved robust predictive performance in cross-sectional and longitudinal datasets. PGS-pretrained models significantly outperformed alternative pretraining outcomes (e.g. family history, cognitive variables), underscoring the specificity. Our model successfully identified key white matter tracts, including the superior longitudinal fasciculus, cortico-spinal tract, corpus callosum, and cingulum, as critical predictors of depression risk. Furthermore, the model demonstrated strong generalizability, accurately classifying children and adolescents at risk for depression in the independent Korean cohort. To our knowledge, this is the first study to pretrain white matter representations using polygenic risk and apply them to psychiatric outcome prediction, with validation conducted longitudinally and cross-cohort. Pretraining the deep learning model with PGS likely enhanced the model’s capacity to detect biologically meaningful white matter features shaped by genetic risk for depression.

Compared to auxiliary pretraining targets such as family history of depression or cognitive variables, our approach yielded the higher classification performance, underscoring the specificity of genetic information in guiding the white matter representation learning.

Depression related PGS captures the cumulative effects of thousands of common variants associated with depression risk, many of which influence neurodevelopmental processes relevant to white matter, including myelination, axonal integrity, and synaptic signaling. Variants such as 5-HTTLPR [50, 51], BDNF [50, 52], SLC6A15 [53], FKBP5-T [54], and COMT [55] have been associated with alterations in white matter microstructure. Beyond individual variants, PGS for depression correlates with FA values, supporting broader genetic influences beyond single-gene effects on white matter integrity [27]. Epigenetic modifications in genes like SLC6A4, COMT, and BDNF [33, 93] may further shape neural pathways by regulating gene expression related to myelination and neural plasticity. During pretraining, the model may have learned to recognize subtle, spatially distributed FA patterns shaped by these genetic factors, enabling more sensitive detection of depression-related white matter deviations during fine-tuning. This approach bridges genetic vulnerability and neurodevelopmental architecture, offering a biologically grounded framework for early risk identification.

Our approach not only yielded improved predictions of depression but also more nuanced insights about neurological correlations of child depression. We observed a widespread distribution of voxels with reduced FA that were significant predictors of youth depression. Individuals with early-onset MDD, particularly those with recurrent or persistent symptoms, showed widespread reductions in FA, which are also broadly influenced by genetic factors [19, 51, 56, 57]. These results emphasize that whole-brain FA alterations may mediate genetic influences on the development of depression.

Despite the importance of whole-brain level white matter microstructures in MDD prediction, we specified several key fasciculi with a high proportion of reduced FA that were critical for predicting youth depression. The superior longitudinal fasciculus, with over 90% of its voxels exhibiting lower FA in youth with an increased likelihood of MDD compared to controls. As a tract connecting frontal, parietal, temporal, and occipital lobes, the superior longitudinal fasciculus has been consistently linked to depression severity, even among asymptomatic youth with familial risk [39, 58]. Meta-analyses further observed the association between the superior longitudinal fasciculus integrity with depression duration and treatment response [59]. Moreover, reduced superior longitudinal fasciculus integrity has been tied to cognitive symptoms related to depression, including deficits in working memory, executive functioning, and attention maintenance [60, 61]. Consistent with prior findings, our results identify the superior longitudinal fasciculus as the most important tract for predicting depression, both cross-sectionally and longitudinally, underscoring its pivotal role in the neurobiology of youth depression.

The cingulum was another significant predictor, with a high proportion of lower FA values contributing critically to the MDD classification. This tract connects the anterior cingulate cortex to the hippocampus and is central to emotional regulation, self-initiative processes, and spatial memory [62, 63], function closely tied to depressive symptoms such as anhedonia, rumination and memory deficits. Disrupted connectivity in the anterior cingulate cortex is a hallmark of adolescent depression [64]. Furthermore, reduced FA in the cingulum has also been consistently associated with suicidal thoughts and behaviors, implicating fronto-limbic dysfunction in both mood dysregulation and self-harm risks [65, 66].

The corpus callosum is a well-established white matter marker in depression, with reduced FA frequently linked to the future symptom severity [32, 67, 68]. Among its subregions, the genu and body show the most pronounced reductions in FA, reflecting impaired interhemispheric connectivity in the frontal lobes, which is critical for emotional and cognitive regulation [69–71]. Animal studies further support this, with reduced FA observed in mice lacking the serotonin transporter gene SLC6A4, which is strongly associated with depressive behaviors [34, 72]. These findings, corroborated by human studies [73], emphasize the central role of the corpus callosum in depression-related white matter alterations.

While our model’s AUROCs in the 0.61-0.66 range fall within the moderate performance band, they consistently surpassed unimodal and conventional machine learning baselines. In the context of low-prevalence, high-stakes conditions such as youth depression [7], even modest improvements in future outcome prediction can provides incremental predictive value. Decision curve analysis (DCA) [74] further demonstrated that our models offer higher net benefit over default strategies such as “treat-all” or “treat-none” approaches. These findings support the clinical utility of biologically informed prediction models, not as stand-alone diagnostics, but as components of multi-tiered screening frameworks.

Our core thesis is that polygenic risk–informed white matter analyses represent a pivotal step forward in the early detection of youth depression, surpassing conventional unimodal approaches in both effect size and cross-ethnic scalability. To aid interpretation, we converted AUROC values to Cohen’s d using the formula *AUROC* = ϕd/√2 (under the equal variance assumption) [75]. We observed estimated cohen’s D of 0.43 (cross-sectional), 0.40 (2-year MDD), 0.51 (suicidal ideation), 0.60 (suicidal attempt), and 0.63 (Korean cohort). These values exceed those reported in large-scale studies where univariate brain features alone often yield modest or non-replicating effects. For example, Van Velzen et al. (2020) reported subtle white matter alterations in adult MDD with d = 0.12–0.26, but adolescent findings did not survive correction [29]. Similarly, large-scale ENIGMA meta-analysis have reported cortical surface area reductions in adolescents with MDD, with effect sizes ranging from *d* = –0.26 to –0.57, as well as subcortical volume differences, including *d* = –0.20 for the hippocampus [76, 77]. In contrast, our gene-informed white matter framework yields medium-to-large effect sizes across cohorts and outcomes, comparable to or exceeding those reported in prior large-scale studies. This supports its potential as a robust tool for identifying youth at risk of depression.

Our moderate-to-large effect sizes persist even in the smaller independent Korean cohort (N=108), highlighting how transfer learning can bolster predictive power in underrepresented samples. Taken together, these findings underscore the promise of gene-informed white matter models for identifying youth at risk of depression – offering a more robust and generalizable signal than has typically been observed with single-modality or univariate approaches.

While dMRI is a relatively costly modality for large-scale screening, its use may be justified in targeted settings—particularly for youth already flagged by clinical or genomic indicators. By combining genetic predispositions and neurobiological markers, our framework demonstrates the feasibility of early, biology-informed identification. Future work incorporating behavioral assessments and environmental exposures may further improve individualized risk prediction and inform early intervention strategies for high-risk youth.

While this study provides valuable insights into youth depression, several limitations remain. First, our framework does not incorporate environmental factors like early stress and socio-economic status. Future work should incorporate these to explore gene-environmental interactions. Second, dMRI has known limitations in resolving crossing fibers [78], although we applied recommended preprocessing pipelines and utilized track-weighted imaging, which has demonstrated higher sensitivity compared to conventional diffusion tensor imaging in representing crossing fiber configurations [79, 80], to mitigate this. Third, our approach excluded other biological modalities, such as brain hippocampal volume and functional connectivity, limiting a comprehensive view of depression. Future research should address these limitations by integrating environmental measures and diverse imaging modalities to enhance prediction and early intervention strategies.

Despite these limitations, our multi-modal integration strategy advances understanding of pediatric depression by demonstrating how PGS-based pretraining can improve neuroimaging-based classification and prognosis in large-scale cohorts. By integrating genetic predispositions and white matter neuroimaging data, we provide a novel framework for identifying children with depression and further at-risk youth before symptom onset. Our findings highlight multiple white matter tracts—including the corpus callosum, superior longitudinal fasciculus, cingulum, and corticospinal tract—as key features for depression prediction. Furthermore, the model’s robust performance in an independent Korean population emphasizes its potential for cross-population generalizability. These findings support a precision psychiatry approach, integrating genetics and neuroimaging for early depression risk detection.

## Methods

### Participants

We used participants from the Adolescent Brain and Cognitive Development (ABCD) study [81] (N=11,868), and a Korean cohort (N=180) recruited through research collaboration with Seoul National University and Ulsan National Institute of Science and Technology.

Following rigorous quality control and exclusion criteria, the final analysis included 5,248 ABCD participants and 108 Korean participants. Detailed demographics and full inclusion/exclusion criteria are provided in Supplementary Materials (Table. S1).

### Track-weighted FA (TW-FA) Imaging Processing

Track-weighted FA (TW-FA) was used as the input in all experiments. dMRI provides measures of brain macrostructure (e.g. volume(mm^3^), length(mm), area(mm^2^)) and microstructure (e.g. white matter integrity), non-invasively [82]. The processing was conducted using software based on the MRtrix3 package [83] especially using the command *tckmap* after general MRI quality assessment. TW-FA maps were formed as described in previous studies [79, 80], so we will describe them briefly here. In summary, reference maps (here, FA maps) values were computed at each point along the streamline. For each voxel, a Gaussian-weighted mean of the reference map values was determined within the neighborhood along individual fiber streamlines (yielding the track-wise statistic). The voxel intensity for the track-weighted imaging map was then calculated as the average of these track-wise values across all tracts passing through the voxel, resulting in the voxel-wise statistic. We utilized full-width-half-maximums (FHWM) of 15mm for gaussian neighborhood weightings, which is known to be more sensitive to focal abnormalities [84]. The TW-FA framework applies directional smoothing to tractograms by assigning each streamline a weight derived from the average FA value along its path, effectively creating a track-informed version of the FA map [84, 85]. TW-FA processing was conducted using the same protocol for both cohorts.

### Demographic and Behavioral Variables

#### Psychiatric Variables of ABCD Cohort

For assessing the children’s lifetime MDD and depressive symptoms, our analysis employed the computerized version of the Kiddie-Structured Assessment for Affective Disorders and Schizophrenia (KSADS-COMP) based on child and parent reports and Child Behavioral Checklist (CBCL) [86], which relies on parent reports. KSADS-COMP has been well validated from previous study showing good to high reliability (AUC 0.89-1.0) compared against clinician administered [87]. For the current analysis, we used a lifetime MDD diagnosis variable encompassing past, present and remitted MDD diagnoses, based on summary ratings from the KSADS-COMP, which considers both child and parent reports. We excluded participants who were taking anti-depressant medication and who met lifetime psychotic (e.g. schizophrenia) or neurodevelopmental disorder (e.g. autism spectrum disorder) diagnoses [88, 89]. For a control group, we included participants who met none of the diagnosis criteria in child nor parent report past or current time point and met healthy control criteria of CBCL (total problems T-score<60) from the baseline to 2-year follow-up. We utilized lifetime indicators of suicidality (including suicidal ideation and suicide attempts) based on KSADS-COMP diagnoses, derived from child and parent reports. The criteria for the control group were consistent with those described earlier.

Since we excluded all the MDD diagnosed participants from pretraining set to maximize the sample size of the finetune set, depressive symptoms score was used exclusively within the pretraining set to estimate the composite polygenic scores. We generated a depressive symptoms score based on a previous study [90], using *module 1* of the KSADS-COMP symptoms assessment, based on summation of the child and parent reports. Detailed methods for composite PGS calculations are described in the Methods section of *Genetic Data Preprocessing*.

In conclusion, 4741 participants were included in the pretraining dataset, and 266 participants (control:133) whose propensity scores were matched for age, biological sex, and study site were used for cross-sectional MDD classification in fine-tune set. In addition, to statistically evaluate the 2-year follow-up zero-shot prediction performance, we utilized individuals with MDD diagnosis at the 2-year follow-up and matched them with randomly sampled from participants not included in either the pretrained or finetune sets (control: 118).

This matching process was repeated 10 times to ensure robustness through different samples, which leads to 10 datasets of 236 participants. For suicidality we used the same strategy, resulting in 10 datasets of 116 (control: 58) participants with MDD and suicidal active ideation prediction at the 2-year follow-up and 30 (control: 15) participants with MDD and suicidal attempt prediction. However, we ensured that none of the participants overlapped with pretraining nor finetune sets. Detailed demographics statistics are provided in Supplementary Materials (Table. S1).

#### Psychiatric Variables of Korean Cohort

Validated Korean translations of depression measures were used to classify participants [91]. Depression was assessed using the child-reported Center for Epidemiological Studies-Depression Scale (CES-D) [92, 93] and parent-reported CBCL [94]. Based on prior meta-analyses, a CES-D cut-off of ≥21, which accounts for gender and racial variations [95, 96], was chosen to align with ABCD study criteria. The depressed group included individuals with CES-D ≥21, while the control group had CES-D <16 and CBCL total problems T-scores <60. The final sample consisted of 108 participants (41 depressed, 67 control). For deep learning, data were split into 10-folds using iterative stratification by age, sex, and CES-D score, followed by propensity score matching for validation and test sets.

#### Genetic Data Preprocessing

In this study, we used polygenic scores (PGS) to estimate individual-specific genetic propensities, which were previously calculated and validated in [97], where a detailed description of the methodology is provided. For further details on genotype preprocessing, see Supplementary Materials. Here, we briefly summarize the key aspects of the PGS calculation. The PGS were calculated using PLINK version 1.9 and the parameters were optimized in PRS-CSx with Bayesian approach [98]. Since our study focused on youth depression in the American population of multi-ancestry, we utilized PGS scores derived from publicly available multi-ancestry Genome-Wide Association Study (GWAS) summary statistics with robust statistical power. PGS related to depression, body mass index, post-traumatic stress disorder, alcohol dependency and schizophrenia were included, as these were constructed using multi-ancestry GWAS summary statistics to ensure applicability across diverse populations.

We included composite PGS in our analysis, guided by prior research, that demonstrated the increased predictive power of composite PGS in enhancing risk stratification [99, 100]. To address multicollinearity, we employed Elastic Net, a robust regularization technique, to estimate beta coefficients for each PGS using KSADS-COMP symptom scores as the outcome variable.

The model was fitted to maximize explained variance (R²) while minimizing the number of independent variables through a stepwise elimination process across 5,000 resampled train-validation-test splits. The final composite score was then computed by summing the selected PGS, each weighted by its corresponding beta coefficient from the best-performing model. We ensured that none of the participants in pretraining sets overlapped with finetune or prediction sets.

### Model Optimization and Evaluation

#### Deep Neural Network Training

We implemented a 3D-Convolution Neural Network (3DCNN) model with densenet3D121 as the backbone architecture across all our experiments. The model consists of an initial 3D convolutional module, four 3D dense-blocks, three 3D Transition layers and one fully connected classifier for each task. In the field of medical imaging, prior studies have demonstrated the superior performance of densenet3D121 model compared to traditional CNNs in tasks such as brain tumor classification [101], and Alzheimer’s disease diagnosis [102]. Additionally, we conducted experiments using a model that incorporated both each participant’s PGS and brain imaging data as input features. This was done by adding the PGS before passing through the fully connected layer of the 3DCNN model, followed by processing through a MLP (3DCNN+MLP). We optimized the model with Mean Squared Error loss for regression tasks and Cross Entropy for the classification tasks. Minimal augmentation was applied, including resizing images to 128×128×128, Random Rotation by 90 degrees (probability=0.5), Random Axis Flip (probability=0.5), and Random Affine Transform (probability=0.5). The deep neural networks were optimized using the AdamW optimizer with a cosine learning rate decay scheduler and a linear warm-up. Additionally, we implemented early stopping with a patience window of 30 epochs to prevent overfitting on the training data.

We employed two types of comparison models to evaluate our approach. The first set of comparison models focused on modality, including unimodal approaches that used either genetic or neuroimaging data alone and a multi-modal model that incorporated both but without pretraining. This comparison aimed to assess whether both genetic and neuroimaging information are essential for detecting children with MDD and to evaluate the benefit of PGS-based pretraining. Specifically, we implemented an MLP model using only PGS as input, a 3DCNN model using only TW-FA, and a from-scratch 3DCNN+MLP model that integrated both PGS and TW-FA as inputs without employing a pretraining strategy. The second set of comparison models included traditional machine learning models: support vector machine with a linear kernel (SVC_linear), support vector machine with an RBF kernel (SVC_rbf), and eXtreme Gradient Boosting (XGBOOST) using FA-weighted structural connectivity and PGS as their inputs. All experiments were conducted using 10-fold cross-validation, ensuring that the same training, validation, and test folds were used consistently across all experiments. Details on the training parameters for pretraining, cross-sectional and 2-year follow-up MDD prediction, along with transfer learning, are available in the Supplementary Materials.

#### Explainable Artificial Intelligence

To identify brain regions contributing most to the model’s predictions, we employed explainable AI (XAI) techniques, specifically Integrated Gradients [48] combined with SmoothGrad [49]. Integrated Gradients generate saliency maps by integrating the gradients of the model’s predictions with respect to the input features, providing a measure of each voxel’s contribution. SmoothGrad enhances this approach by adding Gaussian noise (σ = 0.05) to input images within brain regions, generating 50 noise-augmented images per participant and averaging their resulting saliency maps. This method was implemented using the Captum library (version 0.6.0).

We performed tract-level analyses and identified key tracts containing voxels that significantly contributed to depression classification and prediction. We analyzed only the voxels with statistically significant XAI attribute values between the control and depressed groups. We conducted voxel-wise t-tests with False Discovery Rate (FDR) corrections using the Benjamin-Hochberg method and applied a cluster-based threshold to identify statistically significant regions. Then we calculated the proportion of significant voxels for each tract, by dividing the total number of significant voxels by the total voxels in the tract. This allowed us to determine the positive and negative associations of each tract in the depression classification task.

#### Decision Curve Analysis

To assess the clinical utility of our model, we conducted Decision Curve Analysis (DCA) by computing net benefit across a range of clinically relevant threshold probabilities [74]. Net benefit quantifies the balance between true positives and false positives, incorporating the relative harm of unnecessary interventions. It is calculated as:

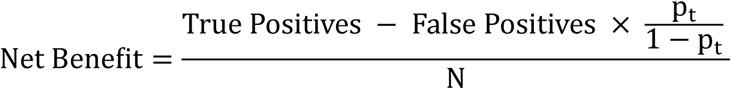

where *p* is the threshold probability, and *N* is the sample size. We compared our best-performing models against two baseline strategies: ‘Treat All’ (assuming all individuals are at risk and should receive intervention) and ‘Treat None’ (assuming no individuals require intervention). A higher net benefit across threshold probabilities indicates superior clinical applicability by improving the identification of at-risk individuals while minimizing unnecessary interventions.

## Data Availability

Multi-cohort data produced are available online at (https://nda.nih.gov/abcd)
Korean independent data produced in the present study are available upon reasonable request to the authors

## Acknowledgements

This work was supported by the National Research Foundation of Korea(NRF) grant funded by the Korea government(MSIT) (No. 2021R1C1C1006503, RS-2023-00266787, RS-2023-00265406, RS-2024-00421268, RS-2024-00342301), by Creative-Pioneering Researchers Program through Seoul National University(No. 200-20240057, 200-20240135), by Semi-Supervised Learning Research Grant by SAMSUNG(No.A0342-20220009), by Identify the network of brain preparation steps for concentration Research Grant by LooxidLabs(No.339-20230001), by Institute of Information & communications Technology Planning & Evaluation (IITP) grant funded by the Korea government(MSIT) [NO.RS-2021-II211343, Artificial Intelligence Graduate School Program (Seoul National University)] by the MSIT(Ministry of Science, ICT), Korea, under the Global Research Support Program in the Digital Field program(RS-2024-00421268) supervised by the IITP(Institute for Information & Communications Technology Planning & Evaluation), by the National Supercomputing Center with supercomputing resources including technical support(KSC-2023-CRE-0568) and by the Ministry of Education of the Republic of Korea and the National Research Foundation of Korea (NRF-2021S1A3A2A02090597, NRF-2023R1A2C2005587, RS-2023-00301976), Seoul National University Center for Happiness Studies (No. 0404-20220001), SNU Creative-Pioneering Researchers Program (339-20230014), Research Grant from Seoul National University(350-20230080) and by Artificial intelligence industrial convergence cluster development project funded by the Ministry of Science and ICT(MSIT, Korea) & Gwangju Metropolitan City.

---

**Table 1.** Cross-sectional MDD Classification 10-fold CV Average TEST Performance Comparison with Modalities.

| model | input | ACC | AUROC | F1 score | stdACC | stdAUROC | std F1 |
| --- | --- | --- | --- | --- | --- | --- | --- |
| ONLY_DEP<br>PGS | DEP PGS | 0.563 | 0.546 | 0.517 | 0.069 | 0.114 | 0.155 |
| Only TW-FA | TW-FA | 0.496 | 0.586 | 0.278 | 0.06 | 0.133 | 0.212 |
| from-scratch<br>3DCNN | TW-FA +<br>DEP PGS | 0.533 | 0.54 | 0.484 | 0.084 | 0.146 | 0.223 |
| DEP PGS<br>pretrained<br>3DCNN | TW-FA | **0.585** | **0.597** | **0.629** | 0.096 | 0.12 | 0.085 |
| DEP PGS<br>pretrained<br>3DCNN | TW-FA +<br>DEP PGS | 0.547 | 0.574 | 0.619 | 0.066 | 0.081 | 0.071 |
| model | input | ACC | AUROC | F1 score | stdACC | stdAUROC | std F1 |
| ONLY cPGS | cPGS | 0.473 | 0.5 | 0.444 | 0.086 | 0.118 | 0.149 |
| Only TW-FA | TW-FA | 0.496 | 0.586 | 0.278 | 0.06 | 0.133 | 0.212 |
| from-scratch<br>3DCNN | TW-FA +<br>cPGS | 0.503 | 0.562 | 0.449 | 0.071 | 0.118 | 0.217 |
| cPGS<br>pretrained<br>3DCNN | TW-FA | **0.577** | **0.62** | 0.636 | 0.094 | 0.121 | 0.072 |
| cPGS<br>pretrained<br>3DCNN | TW-FA +<br>cPGS | 0.564 | 0.6 | **0.638** | 0.033 | 0.127 | 0.052 |

**Table 2.** 2-Year-Follow-up MDD Prediction 10-fold CV Average TEST Performance Comparison with Modalities.

| model | input | ACC | AUROC | F1 score | stdACC | stdAUROC | std F1 |
| --- | --- | --- | --- | --- | --- | --- | --- |
| ONLY DEP PGS | DEP PGS | 0.529 | 0.538 | 0.473 | 0.01 | 0.01 | 0.017 |
| from-scratch 3DCNN | TW-FA | 0.575 | 0.589 | 0.534 | 0.01 | 0.012 | 0.016 |
| from-scratch 3DCNN | TW-FA + DEP PGS | **0.589** | 0.607 | 0.565 | 0.01 | 0.014 | 0.02 |
| DEP PGS pretrained 3DCNN | TW-FA | 0.583 | 0.595 | 0.605 | 0.014 | 0.014 | 0.018 |
| DEP PGS pretrained 3DCNN | TW-FA + DEP PGS | 0.581 | **0.619** | **0.623** | 0.01 | 0.014 | 0.012 |
| model | input | ACC | AUROC | F1 score | stdACC | stdAUROC | std F1 |
| ONLY composite PGS | composit e PGS | 0.536 | 0.562 | 0.474 | 0.009 | 0.011 | 0.016 |
| Only TW-FA | TW-FA | 0.575 | 0.589 | 0.534 | 0.01 | 0.012 | 0.016 |
| from-scratch 3DCNN | TW-FA + composit e PGS | 0.577 | 0.596 | 0.604 | 0.008 | 0.008 | 0.011 |
| composite PGS pretrained 3DCNN | TW-FA | 0.566 | 0.597 | 0.593 | 0.013 | 0.013 | 0.017 |
| composite PGS pretrained 3DCNN | TW-FA + composit e PGS | **0.585** | **0.61** | **0.652** | 0.006 | 0.011 | 0.007 |

**Table 3.**
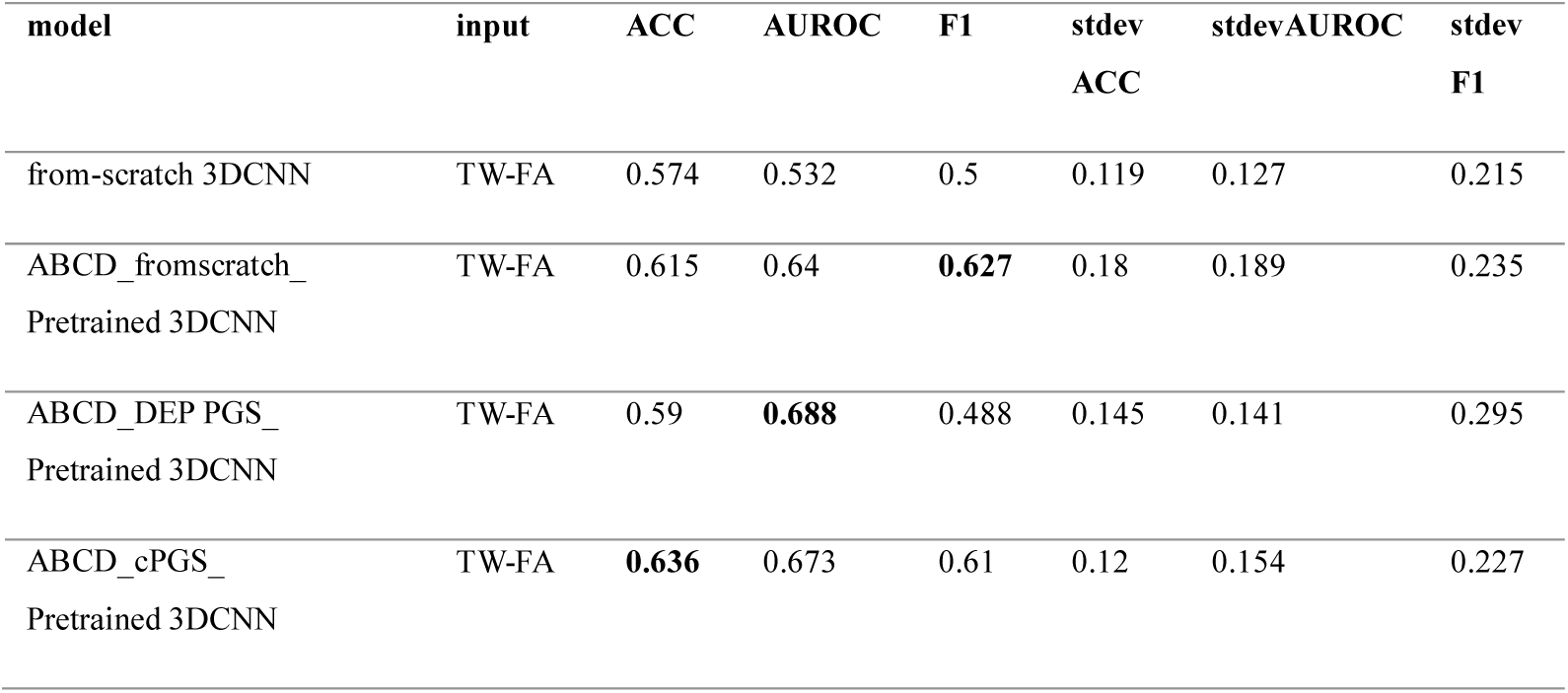
Korean Intendent Set Depression Classification 10-fold CV Average TEST Performance.

## Supplementary Materials

### MRI Acquisition

#### MRI Acquisition of ABCD Cohort

MRI data for the ABCD study was gathered from 21 research sites using Siemens Prisma, GE 750, and Philips Achieva and Ingenia 3T scanners. To maintain uniformity, scanning protocols were standardized across all sites. Detailed descriptions of the imaging protocols for both structural and dMRI have been previously published [1, 2]. The dMRI data were obtained in the axial plane with a 1.7mm isotropic resolution and a multiband acceleration factor of 3. The diffusion-weighted images included seven b=0 s/mm² frames and 96 non-collinear gradient directions, with 6 directions at b=500 s/mm², 15 directions at b=1000 s/mm², 15 directions at b=2000 s/mm², and 60 directions at b=3000 s/mm². T1-weighted images were captured using a 3D magnetization-prepared rapid acquisition gradient echo (MPRAGE) sequence at a 1mm isotropic resolution, without multiband acceleration.

The dMRI data from the ABCD study were acquired and preprocessed following a standardized protocol. For MRI quality assessment, we conducted the ABCD DAIRC’s pipeline. The detailed inclusion criteria are available in the ABCD Release Notes, under ‘MRI QC and Recommended Image Inclusion Criteria’. The DAIRC processed the images with several steps, including correction for eddy current distortion and head motion, adjustment of diffusion gradients, robust diffusion tensor estimation, B0 distortion correction and resampling.

Comprehensive details of the dMRI preprocessing are available in the ABCD Release Notes. We employed MRtrix3 [3] for calculating the diffusion tensor metric, FA, and generating tractography. FA reflects the magnitude and direction of water diffusivity, providing insights into the organization of axons and myelin sheaths [4, 5]. FA values range from 0 to 1, indicating the degree of anisotropy of water molecules and their directional diffusion within each voxel.

Probabilistic tractography was performed using constrained-spherical deconvolution (CSD) [6], with random seeding across the brain and a target streamline count of 20 million. The initial tractograms were filtered using spherical-deconvolution informed filtering (SIFT) (2:1 ratio) [7], yielding a final streamline count of 10 million, which was further down-sampled to 1 million. The computations were conducted using supercomputers at the Argonne Leadership Computing Facility (Theta) and the Texas Advanced Computing Center (Stampede2).

#### MRI Acquisition of Korean Cohort

Diffusion MRI data were acquired using Siemens MAGNETOM Trio Tim 3T scanners with the following parameters: voxel size of 2.3 × 2.3 × 2.3 mm, TR of 3200 ms, TE of 110 ms, and a multi-band acceleration factor of 3. The acquisition included 66 slices in the axial plane with interleaved multi-slice mode. Diffusion-weighted images were obtained using monopolar schemes with b-values of 0 and 3000 s/mm², specifically including 20 directions at b=1000 s/mm², 30 directions at b=2000 s/mm², and 64 directions at b=3000 s/mm². T1-weighted images were collected with a 3D magnetization-prepared rapid acquisition gradient echo (MPRAGE) sequence at 1 mm isotropic resolution without multiband acceleration, providing high-resolution structural data. Probabilistic tractography was performed using CSD, with random seeding across the brain and the target streamline was 1 million. All the preprocessing steps were conducted using the Brainlife platform [8].

#### Participants

We utilized a large-scale neuroimaging, cognitive, and behavioral dataset from 11,868 multiethnic children as part of the Adolescent Brain and Cognitive Development (ABCD) study [1], a nationwide, longitudinal investigation into brain development and child health. For this study, we accessed release 5.1 (http://abcdstudy.org) for demographic, cognitive, psychiatric, and behavioral assessments to correct missing or erroneous values from earlier releases. We used release 2.0 for de-identified diffusion MRI neuroimaging data. We analyzed baseline data (9–10 years old) and 2-year follow-up data (11–12 years old) for psychological assessments.

Participants were excluded based on several criteria: not meeting diffusion MRI quality control standards, not being fluent in English, having a history of severe traumatic brain injury, or being one of multiple births (twins or triplets) from which only one was randomly selected.

Additionally, we included participants who had both genetic and diffusion MRI data. Ultimately, a total of 5,248 participants were included in our analysis (Fig S1.A). The ABCD study was approved by institutional review boards, and informed consent and assent were obtained from all participants and their legal guardians.

We recruited a total of 180 Korean children and adolescents through collaboration with Seoul National University and Ulsan National Institute of Science and Technology. This dataset comprises multi-modal neuroimaging (T1, diffusion MRI, task-functional MRI), behavioral assessments from both participants and their caregivers and genomic data. Participants were excluded if they met any of the following criteria: (1) intellectual disabilities, (2) a history of being diagnosed with psychiatric disorders or learning disabilities, (3) lack of fluency in Korean, (4) color blindness, or (5) conditions that made normal MRI scanning difficult (e.g., presence of metallic implants or potential pregnancy). Among the recruited participants, 150 passed data quality control, which excluded individuals with issues such as excessive head motion or equipment-related problems. Of these, 108 participants who met the criteria for depression or health control groups were included in the final analysis (Fig. S1.B). Participants and their legal guardians provided written informed consent before the experiment, and all procedures were approved by the IRB of Seoul National University. (IRB No. 2112/004-006, IRB No. 2202/003-003)

### Demographic variables

#### Demographic, Biological variables of ABCD Cohort

We used age, biological sex, and ABCD study site data at the baseline year from ABCD release 5.1. To minimize the influence of these variables, we utilized iterative stratification and propensity score matching (PSM) using these variables to match participants in both the validation and test sets across all experimental designs. We used PsmPy for PSM analysis [9].

#### Genetic data preprocessing

In our study, quality control was stringently applied to the genotyping of 733,293 SNPs from saliva DNA samples using the Affymetrix NIDA Smoke Screen Array at Rutgers University Cell and DNA Repository RUCDR. We excluded SNPs and samples with less than 95% call rate and SNPs with a minor allele frequency below 1% using PLINK 1.90. Following this, genotypes were imputed with the 1000 Genomes phase 3 v5 panel [10] and further filtered for high-quality variants using strict criteria, including Hardy-Weinberg equilibrium. Additionally, we utilized PC-Air [11] and PC-Relate [12] to ensure the inclusion of only genetically unrelated individuals from the multi-ethnic ABCD study cohort, ultimately retaining 11,301,999 variants from 8,620 participants. For further detail on these processes, see Joo et al [13].

### Model Optimization and Evaluation

#### Pretraining with PGS

Participants of pretraining set were partitioned into train, validation, test datasets with 10-fold iterative stratification to ensure there was no significant difference in distribution of biological sex, ABCD study site and age (month) and the prediction target outcome, which was PGS. Train, validation, and test were divided in a ratio of 7:2:1. Hyperparameter tuning was performed within the following ranges: learning rate (lr) = {0.01, 0.001}, weight decay (wd) = {0.01, 0.001,0.001,0.00001}, dropout = {0.1, 0.2} and compared validation performance metric to select the best model.

#### Cross-sectional MDD Prediction

Participants were partitioned with 10-fold iterative stratification to ensure there was no significant difference in distribution of biological sex, ABCD study site and age (month) and the prediction target outcome. Train, validation, and test were divided in a ratio of 7:2:1. We conducted PSM to match the participants in the folds using same covariate variables.

Hyperparameter tuning was performed within the following ranges: learning rate (lr) = {0.01, 0.001, 0.0001, 0.00001}, weight decay (wd) = {0.01, 0.001}, dropout = {0.1, 0.2} and compared validation performance metric to select the best model. All the metrics are reported as average values across all repeated settings. The pretrain and finetune sets, while held out from one another, share similar data distributions, and the outcomes for pretraining and finetuning are closely aligned. Also, considering the relatively small size of the finetune set, we chose to freeze the pretrained model’s parameters, allowing only the final layer to be updated during finetuning for fully utilize pre-trained knowledge [14].

#### 2-year follow-up MDD / MDD & Suicidal Behavior Prediction

The dataset for 2-year follow-up predictions excluded participants from cross-sectional tasks. Predictions for future depression KSADS diagnosis were made using an ensemble of the 10 models generated in the cross-sectional MDD KSADS diagnosis classification task. For statistical performance comparison, as previously described, 10 randomly matched datasets were inferred, and the reported performance metrics represent the average across these datasets. The 2-year MDD KSADS diagnosis prediction involved 10 datasets comprising a total of 236 participants. For suicidal behavior prediction, the same strategy was applied, resulting in 10 datasets including 116 participants with MDD and suicidal active ideation at the 2-year follow-up, and 30 participants with MDD and a history of suicide attempts. However, we ensured that none of the participants overlapped with pretraining nor finetune sets.

#### Transfer Learning to Independent Dataset

Participants of Korean datasets were also partitioned through 10-fold iterative stratification to ensure there was no significant difference in distribution of biological sex, age (month) and the prediction target outcome. Train, validation, and test were divided in a ratio of 7:2:1. We conducted PSM to match the participants in the folds using same covariate variables. Hyperparameter tuning was performed within the following ranges: learning rate (lr) = {0.01, 0.001, 0.0001, 0.00001}, weight decay (wd) = {0.01, 0.001}, dropout = {0.1, 0.2} and compared validation performance metric to select the best model. All the metrics are reported in average value of all repeated settings. Considering the ethnic difference between the two datasets, we chose to unfreeze the ABCD MDD classifier model’s parameters.

#### Control Analyses

Additional control analyses confirmed the robustness of our approach. First, PGS-specific pretraining demonstrated superior classification accuracy compared to pretraining with behavioral, family history of depression, and cognitive variables (Table S9). Both PGS-pretrained models significantly outperformed the family history-pretrained model (p < 0.05) and the total intelligence-pretrained model in accuracy (p < 0.02). The cPGS-pretrained model also showed a marginally significant improvement over the CBCL total behavioral problems- pretrained model in F1 score (p < 0.06, Table S28, Fig. S4A–D).

Second, we examined the role of microstructural versus macrostructural information in MDD classification. Models using macrostructural-only inputs (e.g., white matter shape and volume) performed significantly worse in both cross-sectional and 2-year follow-up MDD prediction compared to TW-FA-based models, with statistically significant F1 score differences (p < 0.05, Table S9, Fig. S4E, F).

Third, we conducted comparisons between MDD subgroups, examining differences between those with and without suicidal behaviors and healthy controls (Table S10, S11, Fig. S5A, B). Lastly, we performed additional comparisons incorporating behavioral, age, and sex variables alongside neuroimaging and genetic data, evaluating their combined predictive performance relative to symptom-based models alone (Fig. S6A–D).

## Supplementary Figures

**Supplementary figure 1.**
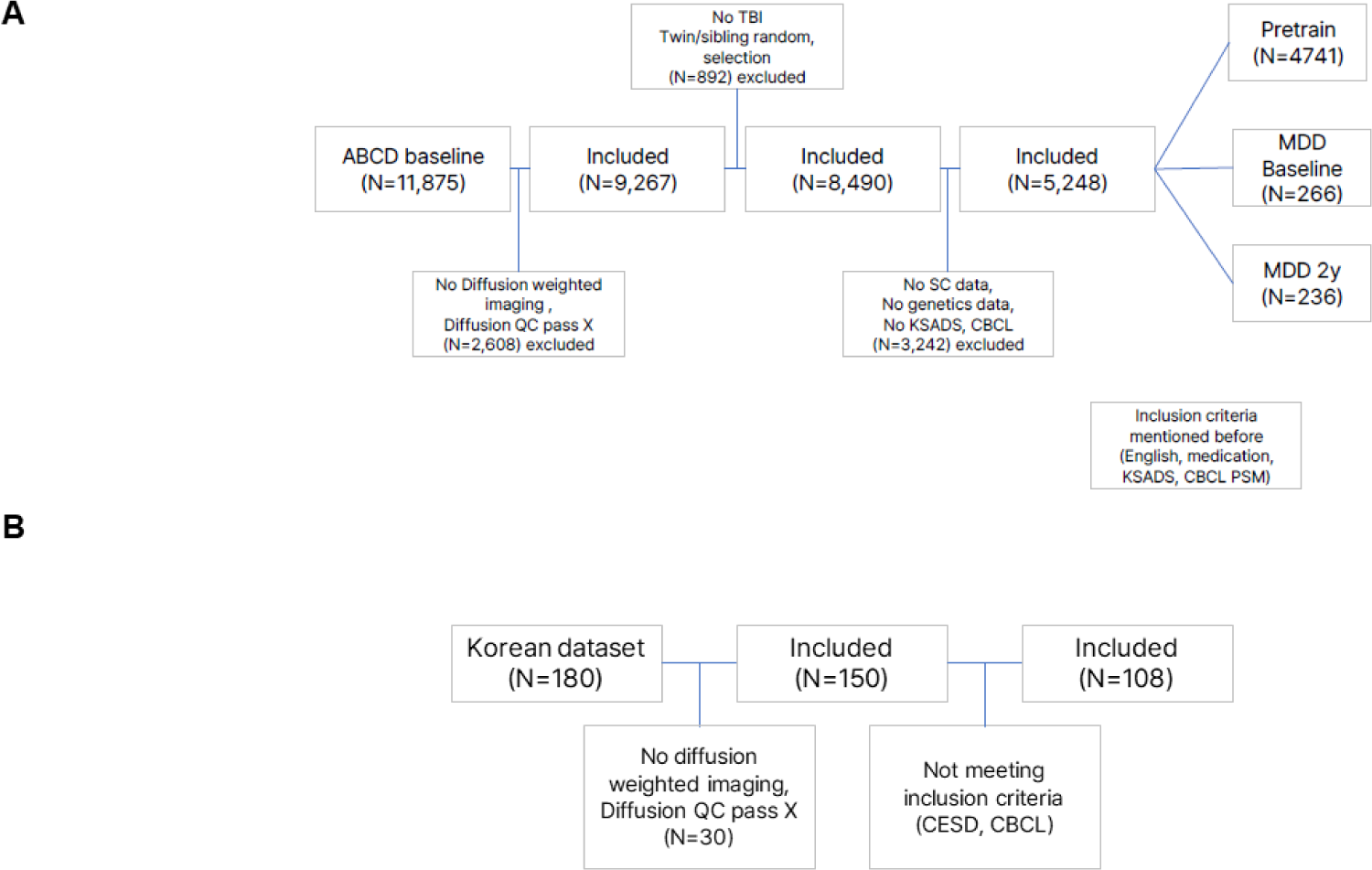
Participants inclusion / exclusion process (A) ABCD participants inclusion / exclusion process, (B) Korean dataset participants inclusion / exclusion process

**Supplementary figure 2.**
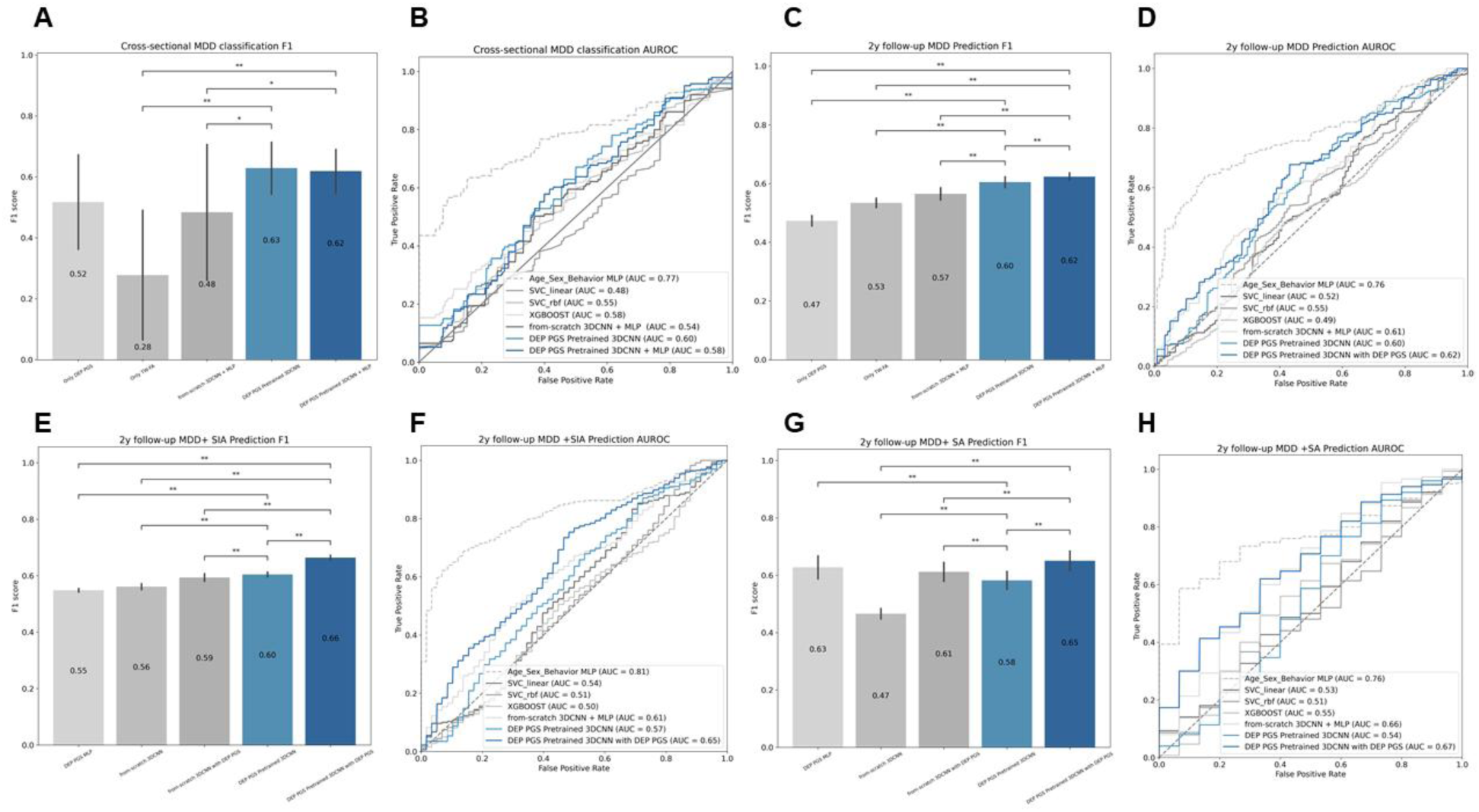
MDD classification task performance using Depression PGS (A) F1 score comparison for cross-sectional MDD classification, showing statistically significant performance differences across models. (B) Receiver operating characteristic (ROC) curves displaying AUROC values for cross-sectional MDD classification models. (C) F1 score comparison for 2-year follow-up MDD prediction, highlighting model performance differences. (D) ROC curves for 2-year follow-up MDD prediction. (E) F1 score comparison for 2-year follow-up MDD and suicidal active ideation prediction. (F) ROC curves for 2-year follow-up MDD and suicidal active ideation prediction. (G) F1 score comparison for 2-year follow-up MDD and suicidal attempt prediction (H) ROC curves for 2-year follow-up MDD and suicidal attempt prediction. (*: p-value < 0.05, **: p-value <0.01, ***p-value <0.0001)

**Supplementary figure 3.**
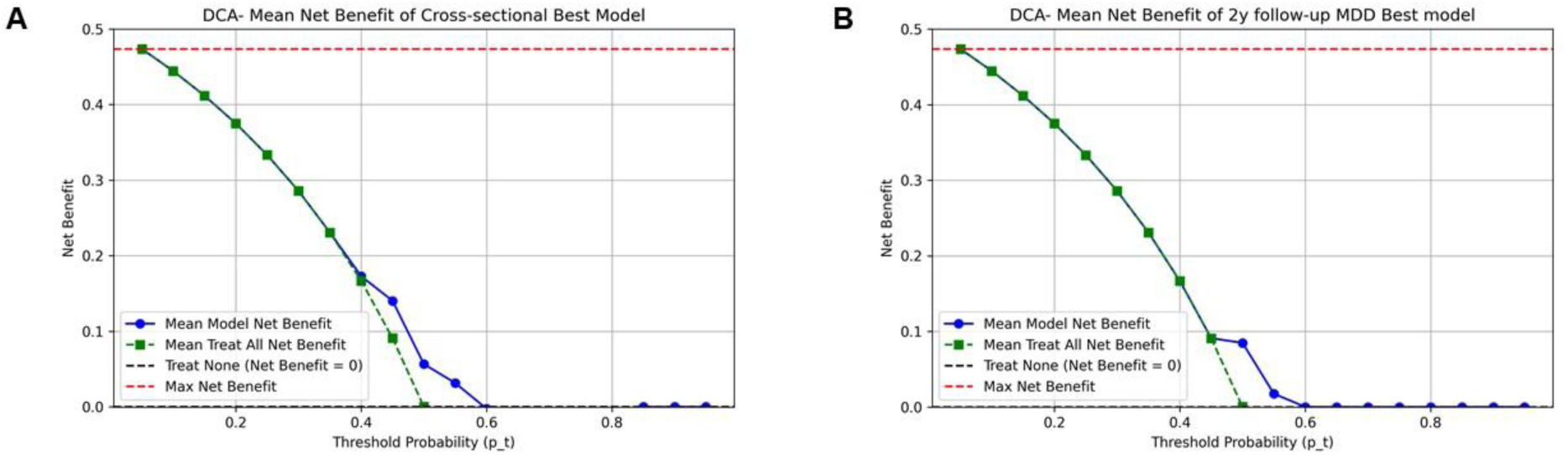
Decision curve analysis (A)Decision curve analysis for cross-sectional MDD Classification. (B) Decision curve analysis for cross-sectional 2y follow-up MDD Prediction.

**Supplementary figure 4.**
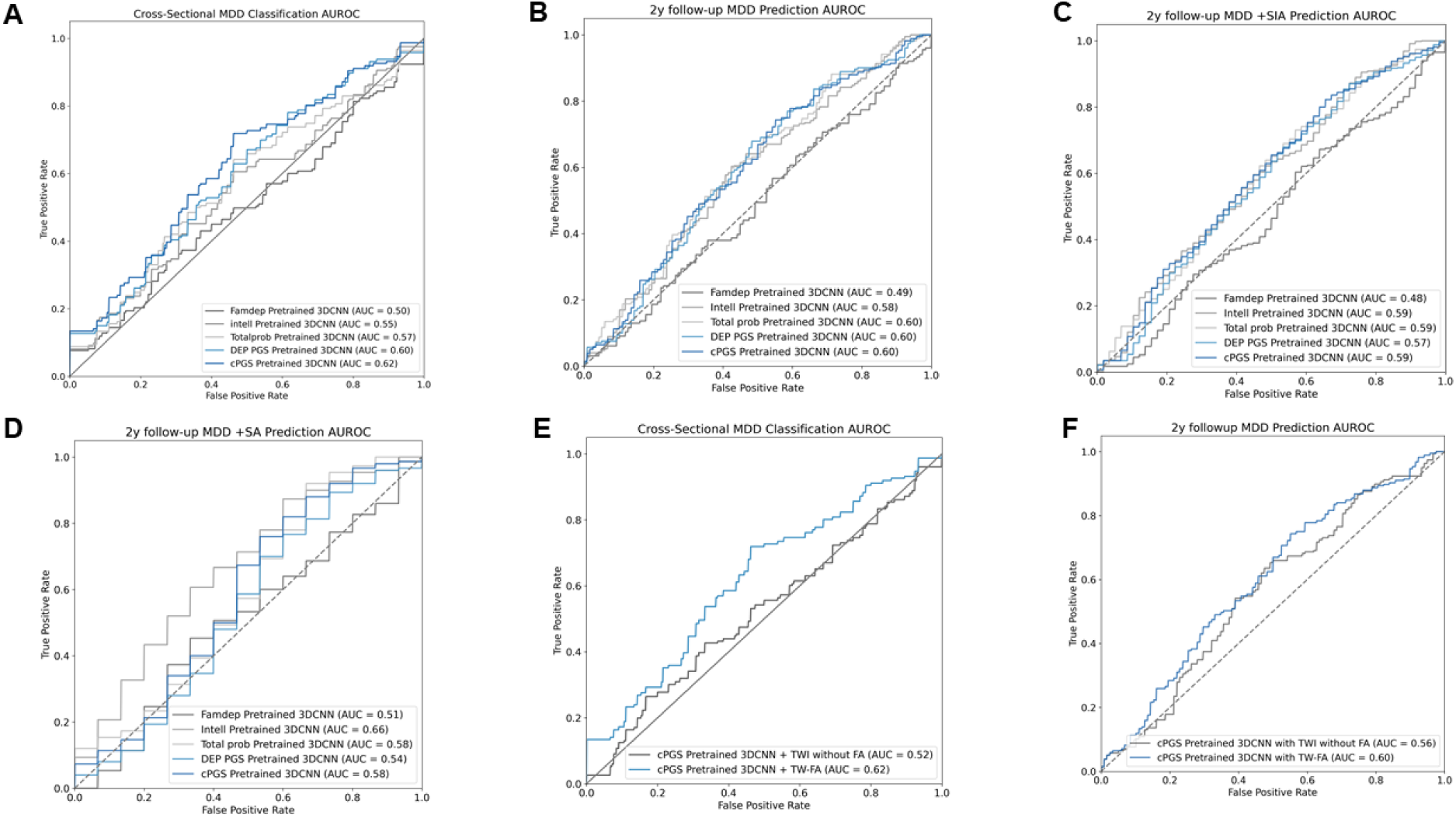
Pretraining with alternative variables (A–D) AUROC curves comparing pretraining with family history of depression (FamDep), intelligence (Intell), and total problem scores (CBCL) instead of PGS for MDD classification and prediction. (E–F) AUROC results for models trained using track-weighted imaging (TWI) features alone, excluding FA values.

**Supplementary figure 5.**
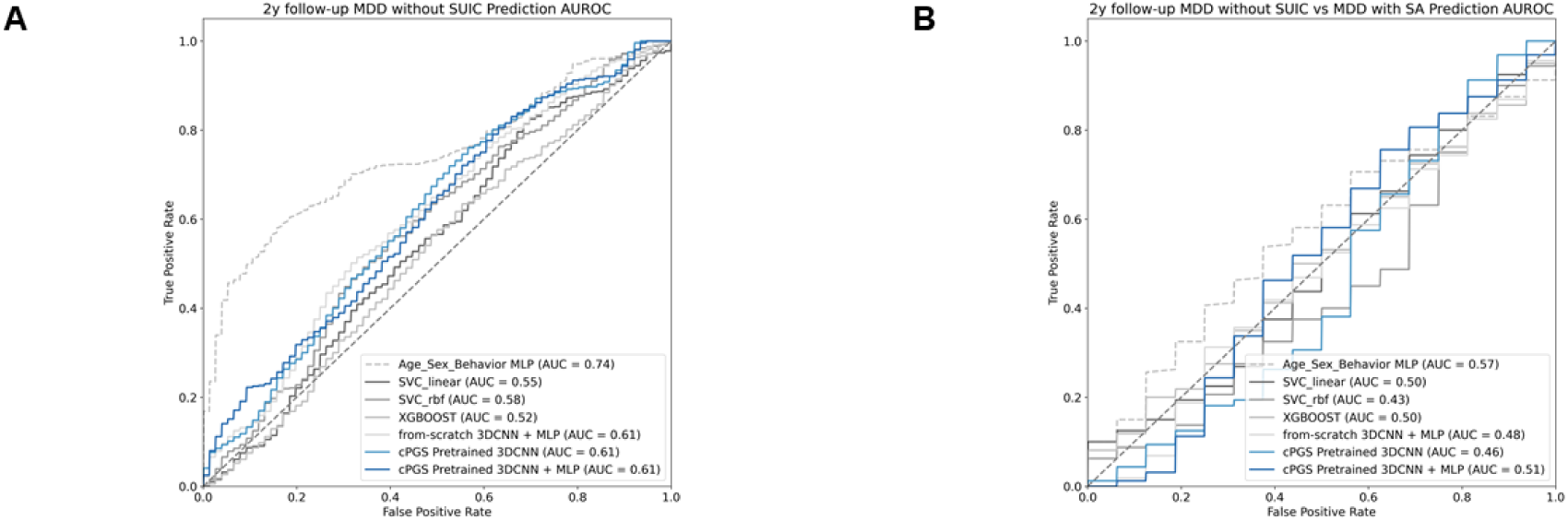
Classification performance for MDD with and without suicidal behavior (A) AUROC curves for distinguishing health controls from MDD without suicidal behavior. (B) AUROC curves for differentiating MDD with suicidal attempts from MDD without suicidal behavior.

**Supplementary figure 6.**
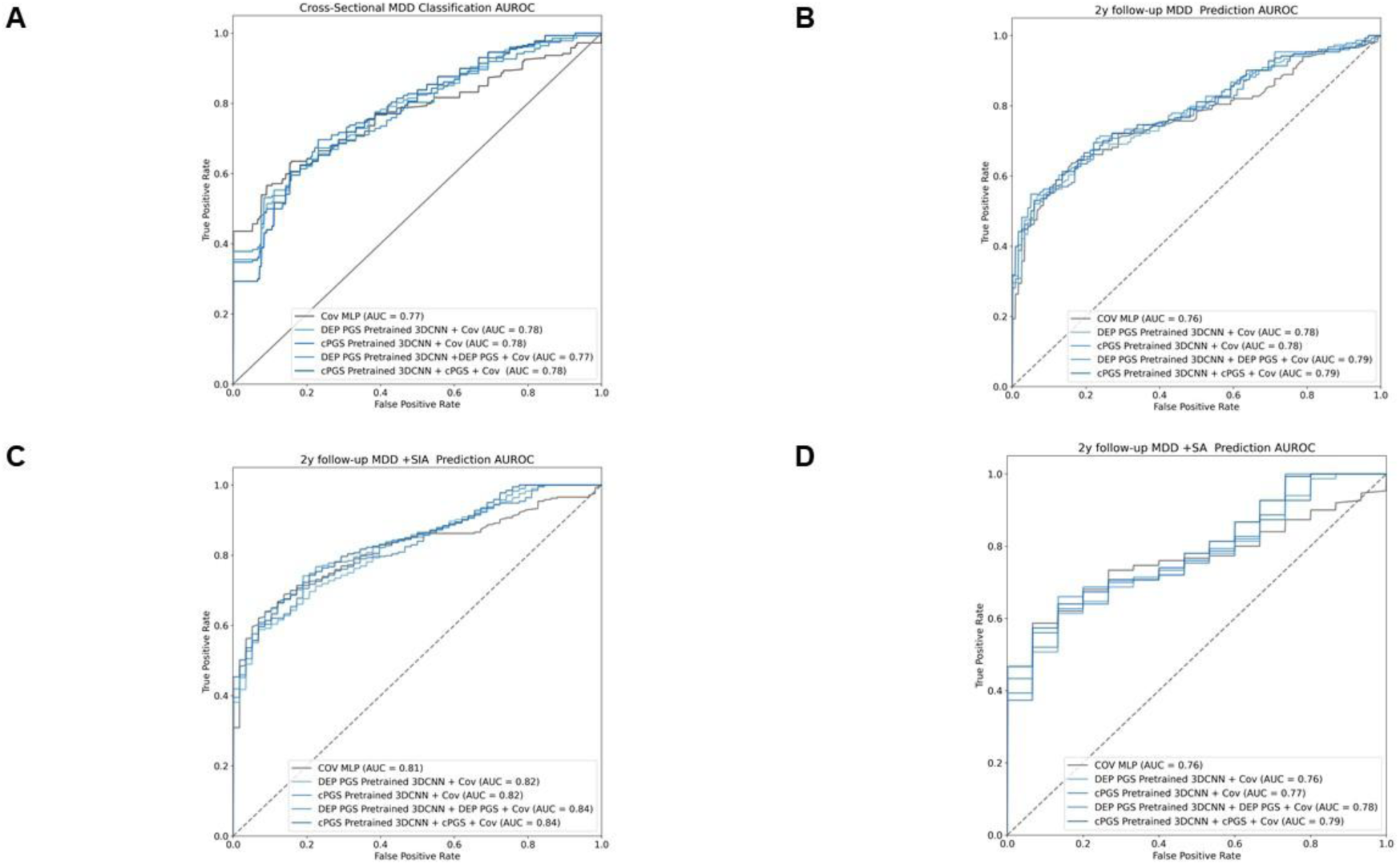
Classification performance for MDD including age, sex, behavioral variables (A) ROC curves displaying AUROC values for cross-sectional MDD classification models. (B) ROC curves for 2-year follow-up MDD prediction. (C) ROC curves for 2-year follow-up MDD and suicidal active ideation prediction. (D) ROC curves for 2-year follow-up MDD and suicidal attempt prediction.

**Supplementary figure 7.**
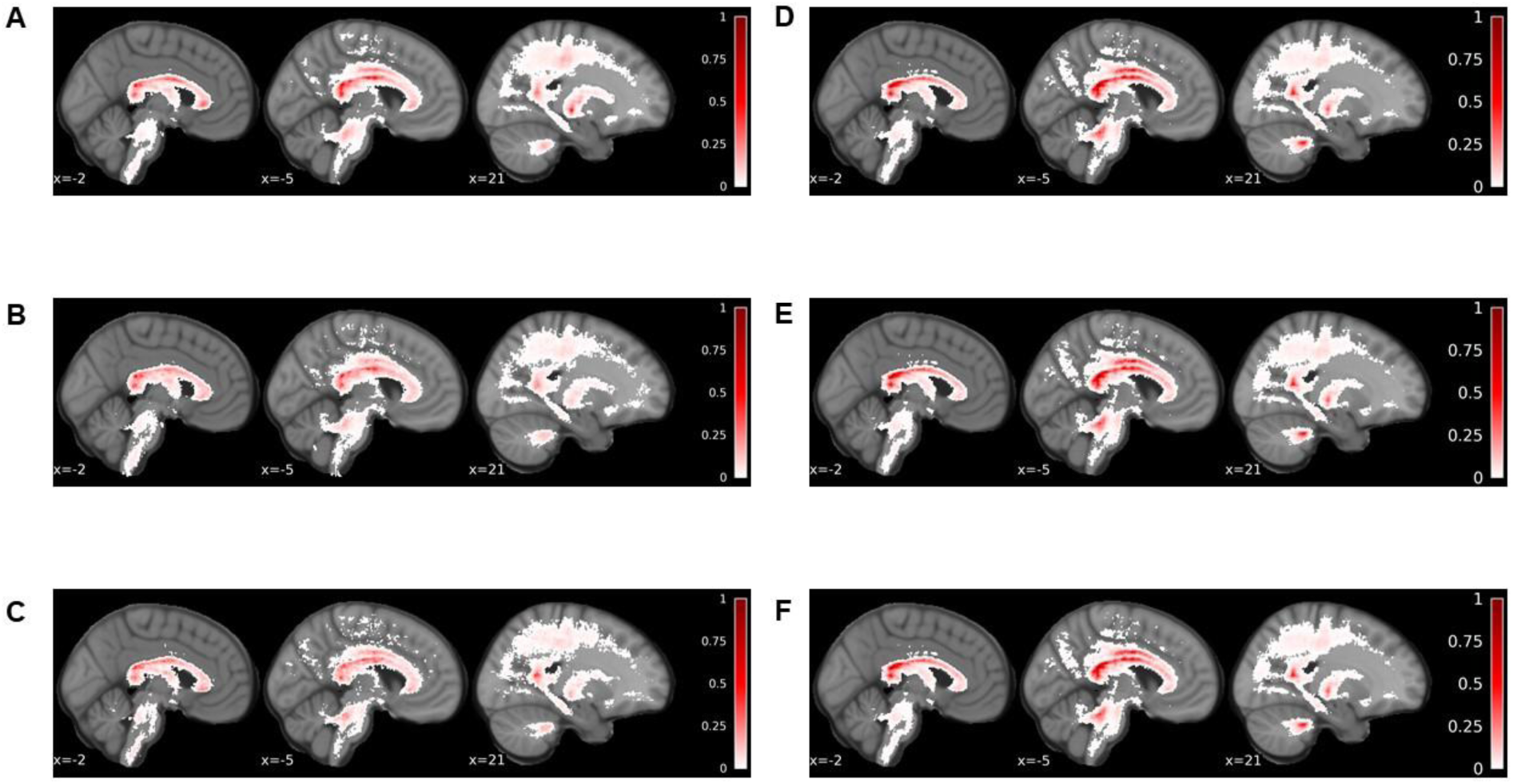
XAI variability (A-C) Cross-sectional MDD classification (D-F) 2-y Follow-up MDD + suicidal ideation Prediction (A) Voxels showing the top 5% variability in True Positive and True Negative individuals. (B) Voxels showing the top 5% variability in True Positive individuals. (C) Voxels showing the top 5% variability in True Negative individuals. (D) Voxels showing the top 5% variability in True Positive and True Negative individuals. (E) Voxels showing the top 5% variability in True Positive individuals. (F) Voxels showing the top 5% variability in True Negative individuals.

## Supplementary Tables

**Supplementary Table 1.** ABCD Demographics of each Datasets.

| ABCD |  | MDD (KSADS) |  |  |  |
| --- | --- | --- | --- | --- | --- |
|  | Covariate | 0 | 1 | p-value | statistics |
| Test | sex | M=251/W=250 | M=61/W=72 | 0.4408 | 0.594299 |
| Chi_square | abcd site (max N) | 16 | 4 | 0.6514 | 17.961729 |
| Chi_square | race_ethnicity |  |  | 0.1269 | 7.174994 |
| Chi_square | white (1) | 278 | 57 |  |  |
|  | black (2) | 58 | 22 |  |  |
|  | hispanic (3) | 115 | 39 |  |  |
|  | asian (4) | 6 | 2 |  |  |
|  | Other | 44 | 13 |  |  |
| T-test | age (mean) | 119.311377 | 119.744361 | 0.5464 | 0.604158 |
| ABCD |  | MDD_2y(KSADS) |  |  |  |
|  | Covariate | 0 | 1 | p-value | statistics |
| Test | sex | M=59/W=59 | M=62/W=92 | 0.1392 | 2.187298 |
| Chi_square | abcd site (max N) | 16 | 16 | 0.2823 | 23.132551 |
| Chi_square | race_ethnicity |  |  | 0.5494 | 3.050694 |
| Chi_square | white (1) | 66 | 76 |  |  |
|  | black (2) | 13 | 24 |  |  |
|  | hispanic (3) | 27 | 38 |  |  |
|  | asian (4) | 1 | 0 |  |  |
|  | Other | 11 | 16 |  |  |
| T-test | age (mean) | 144.466102 | 145.272727 | 0.4105 | 0.824294 |
| ABCD |  | MDD_2y_SIA(KSADS) |  |  |  |
|  | Covariate | 0 | 1 | p-value | statistics |
| Test | sex | M=37/W=44 | M=20/W=38 | 0.2507 | 1.319151 |
| Chi_square | abcd site (max N) | 16 | 16 | 0.4832 | 19.599870 |
| Chi_square | race_ethnicity |  |  | 0.5690 | 2.933352 |
| Chi_square | white (1) | 43 | 27 |  |  |
|  | black (2) | 9 | 10 |  |  |
|  | hispanic (3) | 21 | 13 |  |  |
|  | asian (4) | 1 | 0 |  |  |
|  | Other | 7 | 8 |  |  |
| **T-test** | age (mean) | 143.814815 | 146.948276 | 0.0280 | 2.224974 |
| **ABCD** |  | **MDD_2y_SA(KSADS)** |  |  |  |
|  | **Covariate** | **0** | **1** | **p-value** | **statistics** |
| **Test** | sex | M=32/W=39 | M=4/W=11 | 0.3055 | 1.050169 |
| **Chi_square** | abcd site (max N) | 20 | 20 | 0.5991 | 17.822658 |
| **Chi_square** | race_ethnicity |  |  | 0.1166 | 7.390975 |
| **Chi_square** | white (1) | 37 | 8 |  |  |
|  | black (2) | 7 | 5 |  |  |
|  | hispanic (3) | 19 | 1 |  |  |
|  | asian (4) | 1 | 0 |  |  |
|  | Other | 7 | 1 |  |  |
| **T-test** | age (mean) | 144.507042 | 145.333333 | 0.7214 | 0.361603 |

**Supplementary Table 2.** Korean Dataset Demographics.

| **Korean dataset** |  | **CES-D** |  |  |  |
| --- | --- | --- | --- | --- | --- |
|  | **Covariate** | **0** | **1** | **p-value** | **statistics** |
| **Chi_square** | sex | M=33/W=34 | M=17/W=24 | 0.5558 | 0.347062 |
| **t-test** | age (mean) | 13.164179 | 14.219512 | 0.0055 | 2.841068 |

**Supplementary Table 3.** Cross-sectional MDD classification 10-fold CV Average TEST Performance Comparison with Machine Learning Models.

| **model** | **input** | **ACC** | **AUROC** | **F1 score** | **stdACC** | **stdAUROC** | **std F1** |
| --- | --- | --- | --- | --- | --- | --- | --- |
| SCV_linear | SC + DEP<br>PGS | 0.533 | 0.482 | 0.515 | 0.065 | 0.067 | 0.094 |
| SCV_rbf | SC + DEP<br>PGS | 0.578 | 0.549 | 0.578 | 0.095 | 0.121 | 0.126 |
| XGBOOST | SC + DEP<br>PGS | 0.555 | 0.576 | 0.528 | 0.07 | 0.102 | 0.088 |
| from-scratch<br>3DCNN | TW-FA +<br>DEP PGS | 0.533 | 0.54 | 0.484 | 0.084 | 0.146 | 0.223 |
| DEP PGS<br>pretrained<br>3DCNN | TW-FA | **0.585** | **0.597** | **0.629** | 0.096 | 0.12 | 0.085 |
| DEP PGS<br>pretrained<br>3DCNN | TW-FA +<br>DEP PGS | 0.547 | 0.58 | 0.619 | 0.066 | 0.081 | 0.071 |

| model | input | ACC | AUROC | F1 score | stdACC | stdAUROC | std F1 |
| --- | --- | --- | --- | --- | --- | --- | --- |
| SCV_linear | SC + cPGS | 0.514 | 0.486 | 0.506 | 0.072 | 0.061 | 0.094 |
| SCV_rbf | SC + cPGS | 0.563 | 0.557 | 0.568 | 0.105 | 0.124 | 0.131 |
| XGBOOST | SC + cPGS | 0.566 | 0.588 | 0.536 | 0.073 | 0.092 | 0.093 |
| from-scratch 3DCNN | TW-FA + cPGS | 0.503 | 0.562 | 0.449 | 0.071 | 0.118 | 0.217 |
| cPGS pretrained 3DCNN | TW-FA | **0.577** | **0.62** | 0.636 | 0.094 | 0.121 | 0.072 |
| cPGS pretrained 3DCNN | TW-FA + cPGS | 0.564 | 0.6 | **0.638** | 0.033 | 0.127 | 0.052 |

**Supplementary Table 4.** 2-Year-Followup MDD Prediction 10-fold CV Average TEST Performance Comparison with Machine Learning Models.

| model | input | ACC | AUROC | F1 score | stdACC | stdAUROC | std F1 |
| --- | --- | --- | --- | --- | --- | --- | --- |
| SCV linear | SC + DEP PGS | 0.498 | 0.524 | 0.121 | 0.007 | 0.01 | 0.015 |
| SCV rbf | SC + DEP PGS | 0.497 | 0.548 | 0.348 | 0.009 | 0.01 | 0.019 |
| XGBOOST | SC + DEP PGS | 0.497 | 0.49 | 0.469 | 0.02 | 0.011 | 0.018 |
| from-scratch 3D-CNN | TW-FA + DEP PGS | **0.589** | 0.607 | 0.565 | 0.01 | 0.014 | 0.02 |
| DEP PGS pretrained 3D-CNN | TW-FA | 0.583 | 0.595 | 0.605 | 0.014 | 0.014 | 0.018 |
| DEP PGS pretrained 3D-CNN | TW-FA + DEP PGS | 0.581 | **0.619** | **0.623** | 0.01 | 0.014 | 0.012 |
| model | input | ACC | AUROC | F1 score | stdACC | stdAUROC | std F1 |
| SCV linear | SC + cPGS | 0.493 | 0.522 | 0.106 | 0.009 | 0.01 | 0.018 |
| SCV rbf | SC + cPGS | 0.498 | 0.548 | 0.348 | 0.007 | 0.01 | 0.017 |
| XGBOOST | SC + cPGS | 0.495 | 0.494 | 0.472 | 0.014 | 0.009 | 0.016 |
| from-scratch 3D-CNN | TW-FA + cPGS | 0.577 | 0.596 | 0.604 | 0.008 | 0.008 | 0.011 |
| cPGS pretrained 3D-CNN | TW-FA | 0.566 | 0.597 | 0.593 | 0.013 | 0.013 | 0.017 |
| cPGS pretrained 3D-CNN | TW-FA + cPGS | **0.585** | **0.61** | **0.652** | 0.006 | 0.011 | 0.007 |

**Supplementary Table 5.**
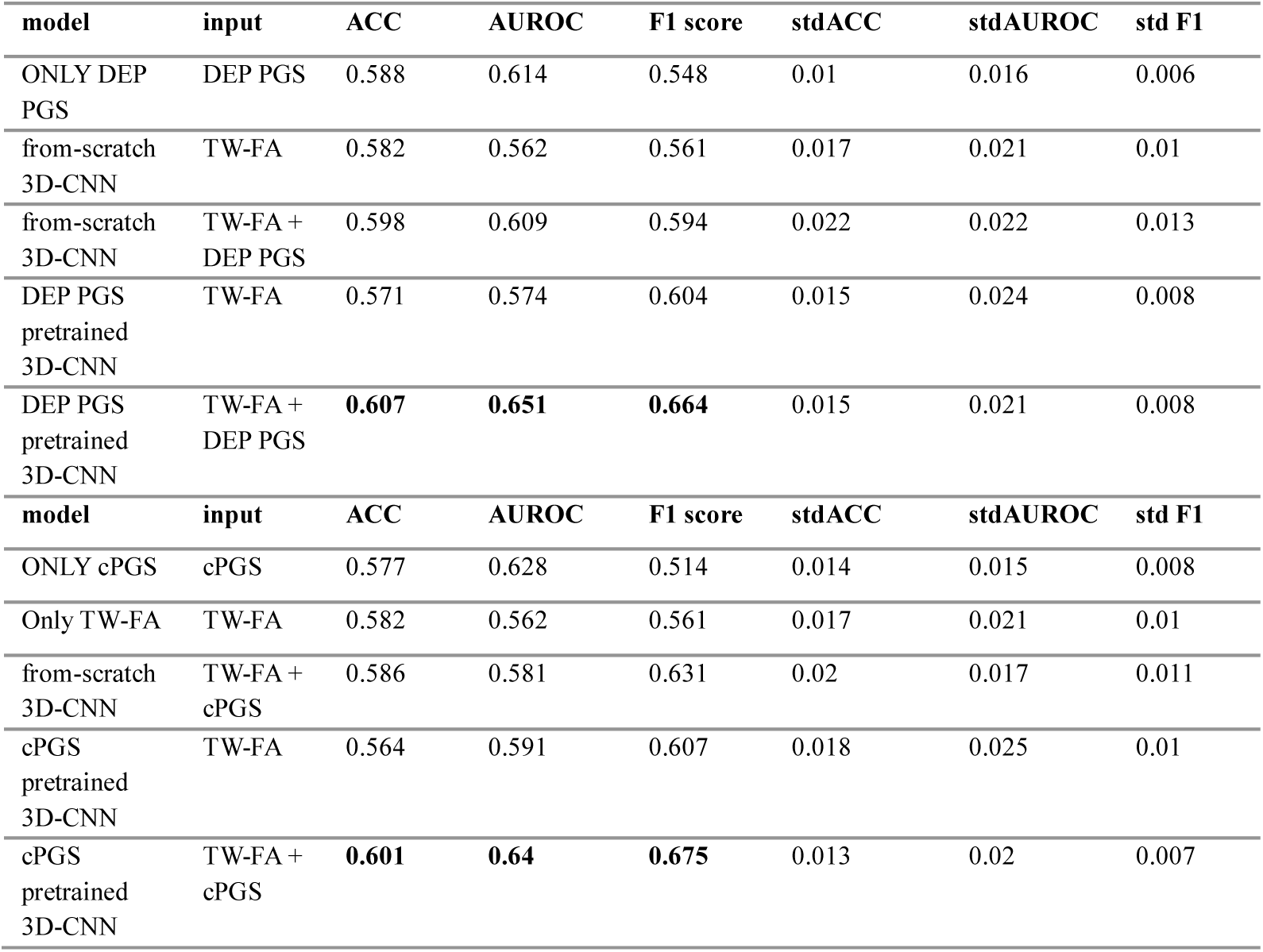
2-Year-Followup MDD + Suicidal Active Ideation Prediction 10-fold CV Average TEST Performance Comparison with Modalities.

**Supplementary Table 6.** 2-Year-Followup MDD + Suicidal Active Ideation Prediction 10-fold CV Average TEST Performance Comparison with Machine Learning Models.

| model | input | ACC | AUROC | F1 score | stdACC | stdAUROC | std F1 |
| --- | --- | --- | --- | --- | --- | --- | --- |
| SCV linear | SC + DEP PGS | 0.521 | 0.535 | 0.17 | 0.011 | 0.024 | 0.018 |
| SCV rbf | SC + DEP PGS | 0.487 | 0.51 | 0.352 | 0.01 | 0.017 | 0.006 |
| XGBOOST | SC + DEP PGS | 0.511 | 0.502 | 0.501 | 0.02 | 0.013 | 0.022 |
| from-scratch 3D-CNN | TW-FA + DEP PGS | 0.598 | 0.609 | 0.594 | 0.022 | 0.022 | 0.013 |
| DEP PGS pretrained 3D-CNN | TW-FA | 0.571 | 0.574 | 0.604 | 0.015 | 0.024 | 0.008 |
| DEP PGS pretrained 3D-CNN | TW-FA + DEP PGS | **0.607** | **0.651** | **0.664** | 0.015 | 0.021 | 0.008 |
| model | input | ACC | AUROC | F1 score | stdACC | stdAUROC | std F1 |
| SCV linear | SC + cPGS | 0.52 | 0.536 | 0.167 | 0.011 | 0.025 | 0.014 |
| SCV rbf | SC + cPGS | 0.486 | 0.507 | 0.352 | 0.014 | 0.018 | 0.009 |
| XGBOOST | SC + cPGS | 0.507 | 0.496 | 0.502 | 0.021 | 0.01 | 0.021 |
| from-scratch 3D-CNN | TW-FA + cPGS | 0.586 | 0.581 | 0.631 | 0.02 | 0.017 | 0.011 |
| cPGS<br>pretrained<br>3D-CNN | TW-FA | 0.564 | 0.591 | 0.607 | 0.018 | 0.025 | 0.01 |
| cPGS<br>pretrained<br>3D-CNN | TW-FA +<br>cPGS | **0.601** | **0.64** | **0.675** | 0.013 | 0.02 | 0.007 |

**Supplementary Table 7.** 2-Year-Followup MDD + Suicidal attempt Prediction 10-fold CV Average TEST Performance Comparison with Modalities.

| model | input | ACC | AUROC | F1 score | stdACC | stdAUROC | std F1 |
| --- | --- | --- | --- | --- | --- | --- | --- |
| ONLY DEP<br>PGS | DEP PGS | **0.64** | **0.716** | 0.628 | 0.063 | 0.057 | 0.04 |
| from-scratch<br>3D-CNN | TW-FA | 0.54 | 0.573 | 0.466 | 0.033 | 0.062 | 0.018 |
| from-scratch<br>3D-CNN | TW-FA +<br>DEP PGS | 0.617 | 0.661 | 0.612 | 0.052 | 0.074 | 0.033 |
| DEP PGS<br>pretrained<br>3D-CNN | TW-FA | 0.567 | 0.543 | 0.582 | 0.054 | 0.081 | 0.031 |
| DEP PGS<br>pretrained<br>3D-CNN | TW-FA +<br>DEP PGS | 0.603 | 0.67 | **0.651** | 0.055 | 0.076 | 0.034 |
| model | input | ACC | AUROC | F1 score | stdACC | stdAUROC | std F1 |
| ONLY cPGS | cPGS | **0.633** | 0.664 | 0.622 | 0.049 | 0.051 | 0.031 |
| Only TW-FA | TW-FA | 0.54 | 0.573 | 0.466 | 0.033 | 0.062 | 0.018 |
| from-scratch<br>3D-CNN | TW-FA +<br>cPGS | 0.607 | 0.611 | 0.646 | 0.049 | 0.061 | 0.026 |
| cPGS<br>pretrained<br>3D-CNN | TW-FA | 0.62 | 0.585 | 0.661 | 0.064 | 0.097 | 0.038 |
| cPGS<br>pretrained<br>3D-CNN | TW-FA +<br>cPGS | 0.63 | **0.665** | **0.702** | 0.06 | 0.071 | 0.034 |

**Supplementary Table 8.**
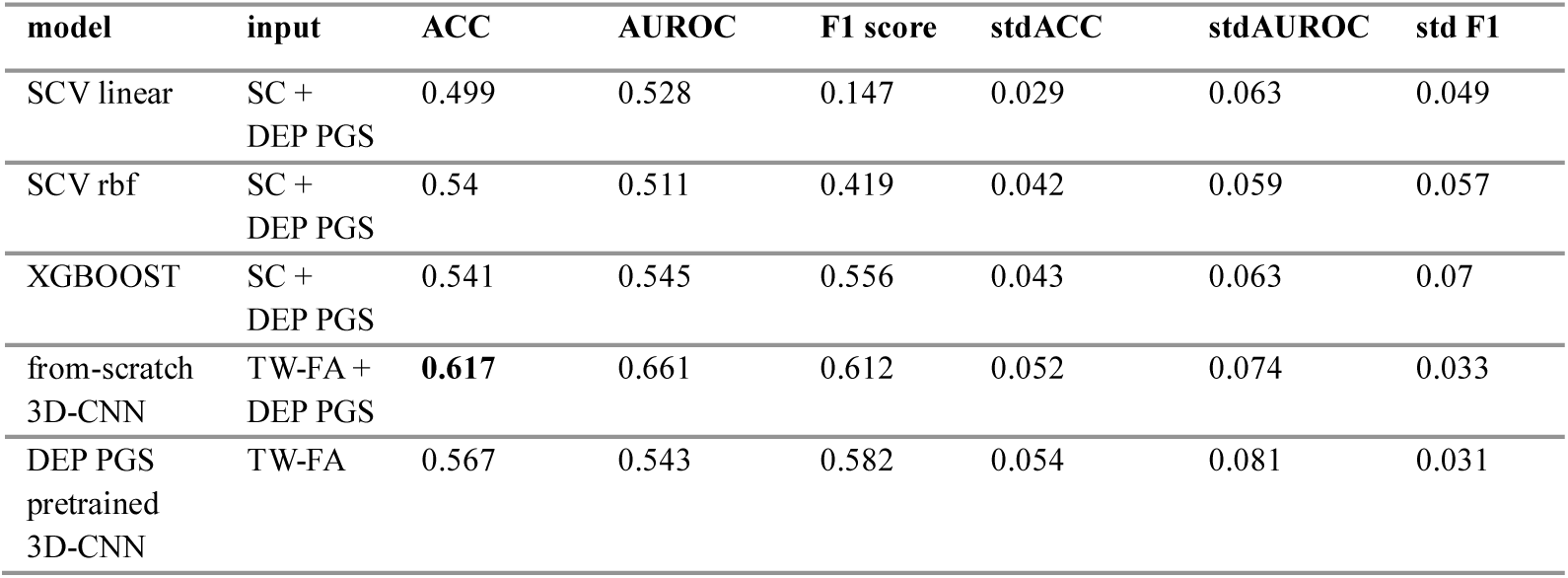
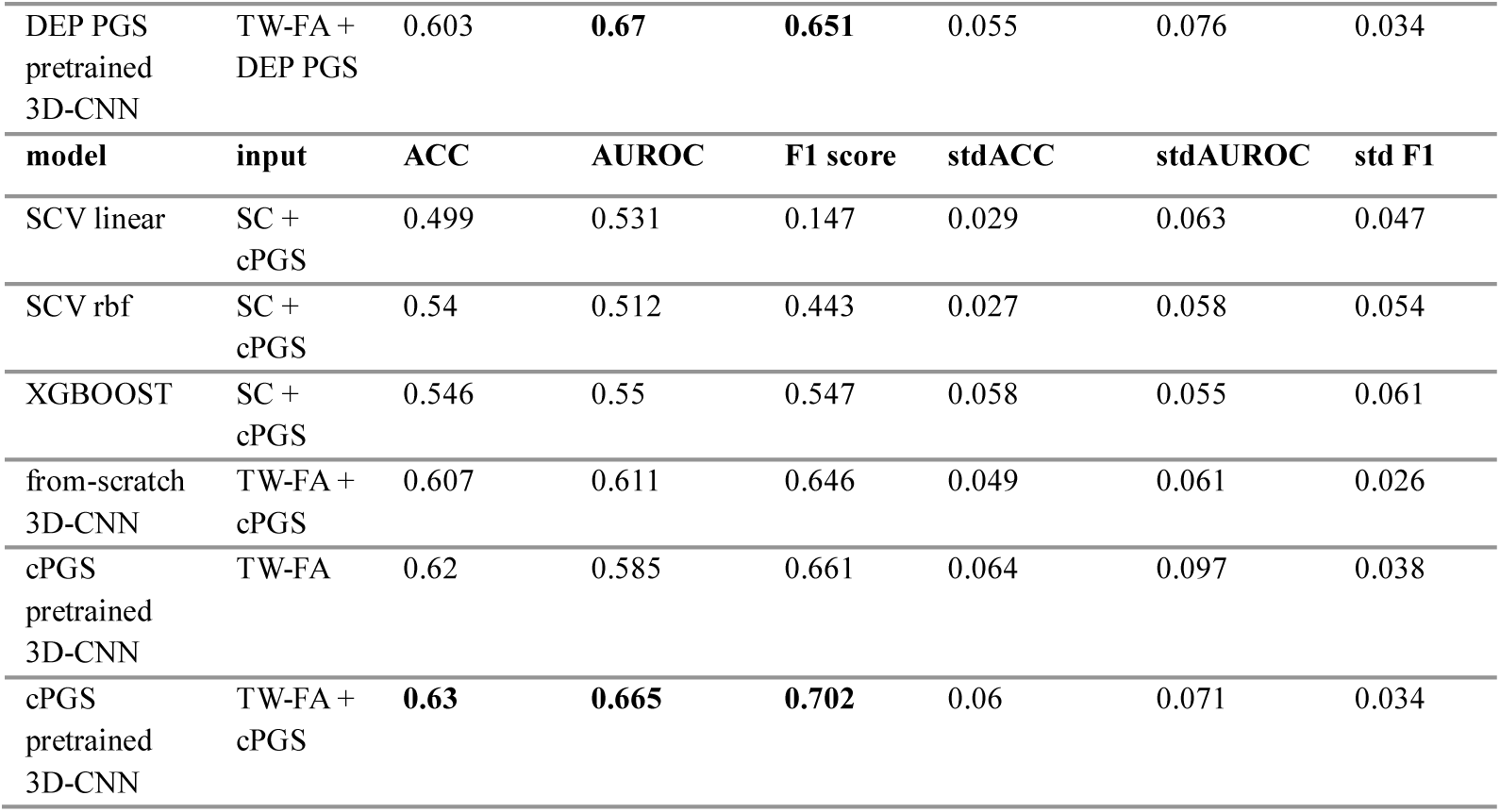
2-Year-Followup MDD + Suicidal attempt Prediction 10-fold CV Average TEST Performance Comparison with Machine Learning Models.

**Supplementary Table 9.** Control Analysis Performance.

| model | input | ACC | AUROC | F1 score | stdACC | stdAUROC | std F1 |
| --- | --- | --- | --- | --- | --- | --- | --- |
| cPGS<br>pretrained<br>3DCNN | TW-intensity<br>image<br>(tractogram) | 0.513 | 0.522 | 0.354 | 0.059 | 0.095 | 0.262 |
| CBCL total<br>behavioral<br>problems<br>pretrained<br>3DCNN | TW-FA | 0.555 | 0.572 | 0.488 | 0.096 | 0.137 | 0.187 |
| Total<br>intelligence<br>pretrained<br>3DCNN | TW-FA | 0.492 | 0.553 | 0.583 | 0.06 | 0.102 | 0.083 |
| Family<br>Depression<br>history<br>pretrained<br>3DCNN | TW-FA | 0.514 | 0.498 | 0.460 | 0.056 | 0.145 | 0.196 |

**Supplementary Table 10.** 2-Year-Followup MDD without Suicidal behavior Prediction 10-fold CV Average TEST Performance Comparison with Modalities.

Average TEST Performance Comparison with Modalities
| model | input | ACC | AUROC | F1 score | stdACC | stdAUROC | std F1 |
| --- | --- | --- | --- | --- | --- | --- | --- |
| ONLY_DEP<br>PGS | DEP PGS | 0.491 | 0.499 | 0.402 | 0.013 | 0.02 | 0.006 |

| from-scratch<br>3DCNN | TW-FA | 0.578 | 0.611 | 0.536 | 0.017 | 0.025 | 0.01 |
| --- | --- | --- | --- | --- | --- | --- | --- |
| from-scratch<br>3DCNN | TW-FA +<br>DEP PGS | 0.576 | **0.615** | 0.541 | 0.015 | 0.025 | 0.009 |
| DEP PGS<br>pretrained<br>3DCNN | TW-FA | **0.587** | 0.612 | **0.61** | 0.019 | 0.022 | 0.011 |
| DEP PGS<br>pretrained<br>3DCNN | TW-FA +<br>DEP PGS | 0.557 | 0.608 | 0.593 | **0.02** | **0.022** | **0.011** |
| model | input | ACC | AUROC | F1 score | stdACC | stdAUROC | std F1 |
| ONLY cPGS | cPGS | 0.506 | 0.532 | 0.427 | 0.015 | 0.021 | 0.008 |
| Only TW-FA | TW-FA | 0.578 | 0.611 | 0.536 | 0.017 | 0.025 | 0.01 |
| from-scratch<br>3DCNN | TW-FA +<br>cPGS | 0.577 | 0.607 | 0.599 | 0.02 | 0.024 | 0.011 |
| cPGS<br>pretrained<br>3DCNN | TW-FA | 0.575 | 0.611 | 0.598 | 0.024 | 0.025 | 0.013 |
| cPGS<br>pretrained<br>3DCNN | TW-FA +<br>cPGS | **0.593** | **0.612** | **0.66** | **0.022** | **0.024** | **0.012** |

**Supplementary Table 11.** 2-Year-Followup MDD without Suicidal behavior Prediction 10-fold CV Average TEST Performance Comparison with Machine Learning Models All the Wilcoxon rank sum test analysis were conducted through python, utilizing *statsmodels*.

| model | input | ACC | AUROC | F1 score | stdACC | stdAUROC | std F1 |
| --- | --- | --- | --- | --- | --- | --- | --- |
| SCV_linear | SC + DEP<br>PGS | 0.478 | 0.547 | 0.057 | 0.013 | 0.017 | 0.019 |
| SCV_rbf | SC + DEP<br>PGS | 0.505 | 0.581 | 0.34 | 0.016 | 0.017 | 0.026 |
| XGBOOST | SC + DEP<br>PGS | 0.526 | 0.521 | 0.5 | 0.019 | 0.019 | 0.027 |
| from-scratch<br>3DCNN | TW-FA +<br>DEP PGS | 0.576 | **0.615** | 0.541 | 0.015 | 0.025 | 0.009 |
| DEP PGS<br>pretrained<br>3DCNN | TW-FA | **0.587** | 0.612 | **0.61** | 0.019 | 0.022 | 0.011 |
| DEP PGS<br>pretrained<br>3DCNN | TW-FA +<br>DEP PGS | 0.557 | 0.608 | 0.593 | **0.02** | **0.022** | **0.011** |
| model | input | ACC | AUROC | F1 score | stdACC | stdAUROC | std F1 |
| SCV_linear | SC + cPGS | 0.483 | 0.545 | 0.064 | 0.012 | 0.018 | 0.024 |
| SCV_rbf | SC + cPGS | 0.506 | 0.582 | 0.341 | 0.017 | 0.017 | 0.027 |
| XGBOOST | SC + cPGS | 0.525 | 0.523 | 0.497 | 0.026 | 0.022 | 0.037 |
| from-scratch<br>3DCNN | TW-FA +<br>cPGS | 0.577 | 0.607 | 0.599 | 0.02 | 0.024 | 0.011 |
| cPGS<br>pretrained<br>3DCNN | TW-FA | 0.575 | **0.609** | 0.598 | 0.024 | 0.025 | 0.013 |
| cPGS<br>pretrained<br>3DCNN | TW-FA +<br>cPGS | **0.593** | 0.607 | **0.66** | **0.022** | **0.024** | **0.012** |

**Supplementary Table 12.**
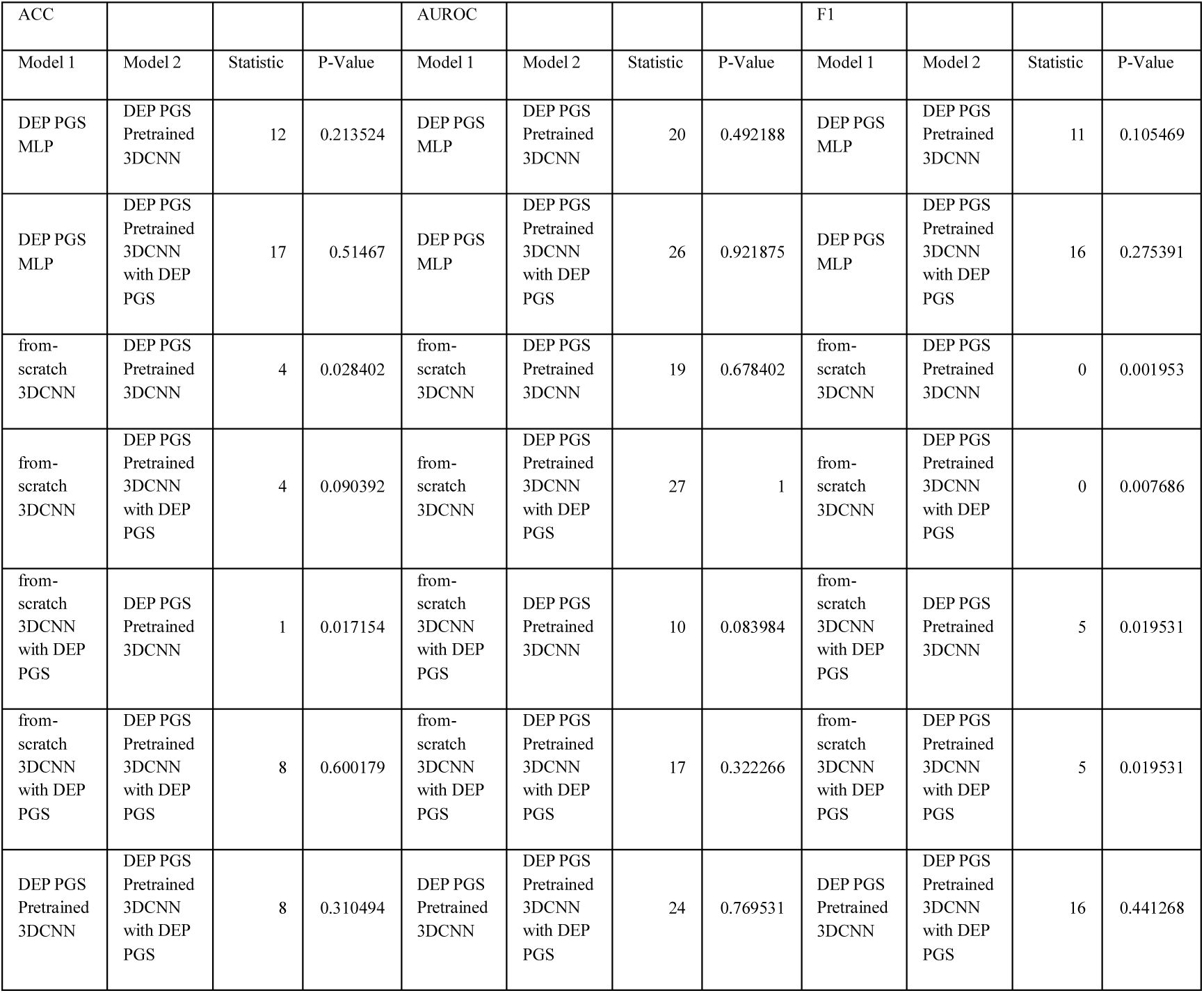
Cross-sectional MDD Classification Model Comparison Modalities (DEP PGS)

| ACC |  |  |  | AUROC |  |  |  | F1 |  |  |  |
| --- | --- | --- | --- | --- | --- | --- | --- | --- | --- | --- | --- |
| Model 1 | Model 2 | Statistic | P-Value | Model 1 | Model 2 | Statistic | P-Value | Model 1 | Model 2 | Statistic | P-Value |
| DEP PGS MLP | DEP PGS Pretrained 3DCNN | 12 | 0.213524 | DEP PGS MLP | DEP PGS Pretrained 3DCNN | 20 | 0.492188 | DEP PGS MLP | DEP PGS Pretrained 3DCNN | 11 | 0.105469 |
| DEP PGS MLP | DEP PGS Pretrained 3DCNN with DEP PGS | 17 | 0.51467 | DEP PGS MLP | DEP PGS Pretrained 3DCNN with DEP PGS | 26 | 0.921875 | DEP PGS MLP | DEP PGS Pretrained 3DCNN with DEP PGS | 16 | 0.275391 |
| from-scratch 3DCNN | DEP PGS Pretrained 3DCNN | 4 | 0.028402 | from-scratch 3DCNN | DEP PGS Pretrained 3DCNN | 19 | 0.678402 | from-scratch 3DCNN | DEP PGS Pretrained 3DCNN | 0 | 0.001953 |
| from-scratch 3DCNN | DEP PGS Pretrained 3DCNN with DEP PGS | 4 | 0.090392 | from-scratch 3DCNN | DEP PGS Pretrained 3DCNN with DEP PGS | 27 | 1 | from-scratch 3DCNN | DEP PGS Pretrained 3DCNN with DEP PGS | 0 | 0.007686 |
| from-scratch 3DCNN with DEP PGS | DEP PGS Pretrained 3DCNN | 1 | 0.017154 | from-scratch 3DCNN with DEP PGS | DEP PGS Pretrained 3DCNN | 10 | 0.083984 | from-scratch 3DCNN with DEP PGS | DEP PGS Pretrained 3DCNN | 5 | 0.019531 |
| from-scratch 3DCNN with DEP PGS | DEP PGS Pretrained 3DCNN with DEP PGS | 8 | 0.600179 | from-scratch 3DCNN with DEP PGS | DEP PGS Pretrained 3DCNN with DEP PGS | 17 | 0.322266 | from-scratch 3DCNN with DEP PGS | DEP PGS Pretrained 3DCNN with DEP PGS | 5 | 0.019531 |
| DEP PGS Pretrained 3DCNN | DEP PGS Pretrained 3DCNN with DEP PGS | 8 | 0.310494 | DEP PGS Pretrained 3DCNN | DEP PGS Pretrained 3DCNN with DEP PGS | 24 | 0.769531 | DEP PGS Pretrained 3DCNN | DEP PGS Pretrained 3DCNN with DEP PGS | 16 | 0.441268 |

**Supplementary Table 13.**
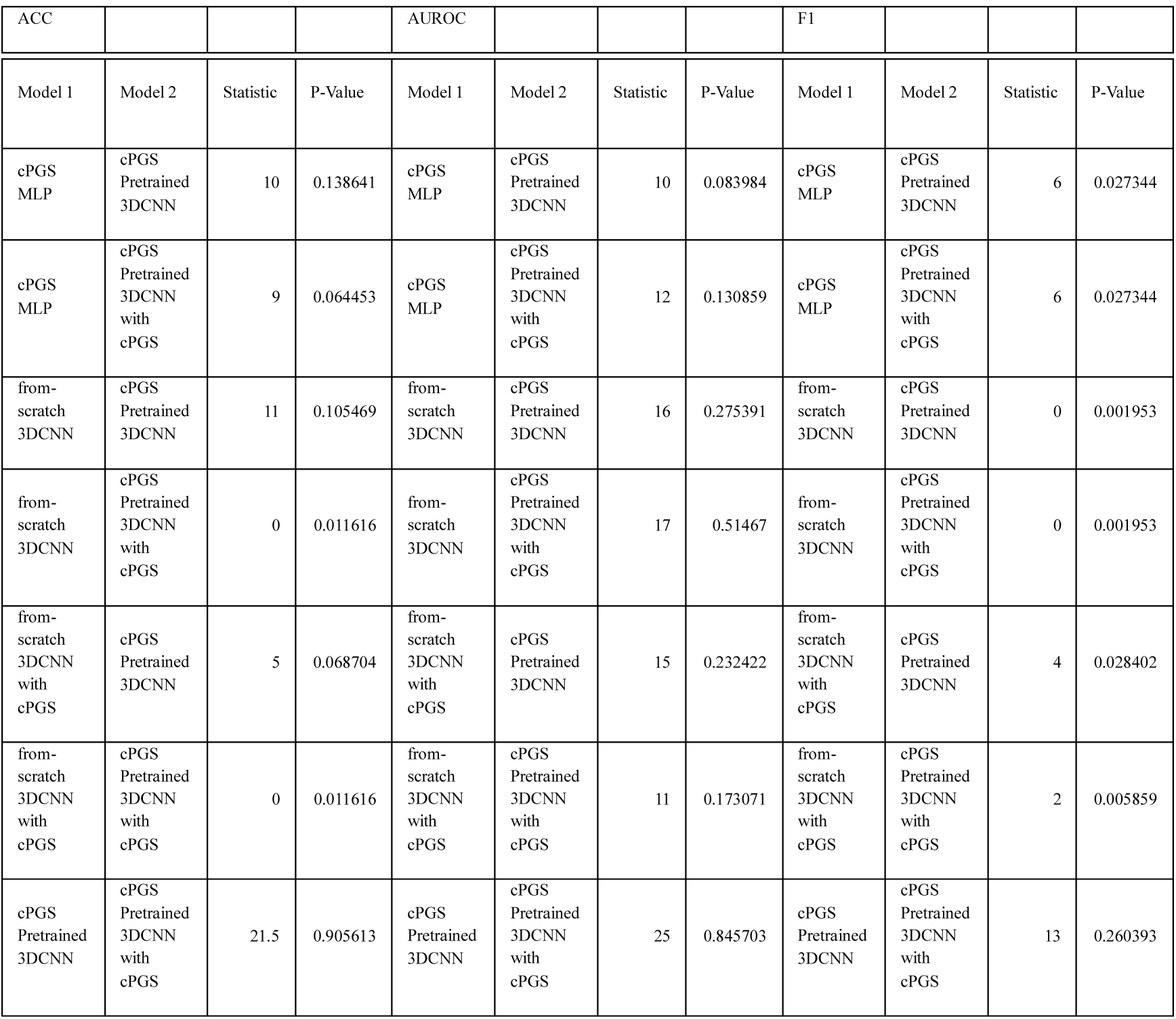
Cross-sectional MDD Classification Model Comparison Modalities (cPGS)

| Model 1 | Model 2 | Statistic | P-Value | Model 1 | Model 2 | Statistic | P-Value | Model 1 | Model 2 | Statistic | P-Value |
| --- | --- | --- | --- | --- | --- | --- | --- | --- | --- | --- | --- |
| cPGS MLP | cPGS Pretrained 3DCNN | 10 | 0.138641 | cPGS MLP | cPGS Pretrained 3DCNN | 10 | 0.083984 | cPGS MLP | cPGS Pretrained 3DCNN | 6 | 0.027344 |
| cPGS MLP | cPGS Pretrained 3DCNN with cPGS | 9 | 0.064453 | cPGS MLP | cPGS Pretrained 3DCNN with cPGS | 12 | 0.130859 | cPGS MLP | cPGS Pretrained 3DCNN with cPGS | 6 | 0.027344 |
| from-scratch 3DCNN | cPGS Pretrained 3DCNN | 11 | 0.105469 | from-scratch 3DCNN | cPGS Pretrained 3DCNN | 16 | 0.275391 | from-scratch 3DCNN | cPGS Pretrained 3DCNN | 0 | 0.001953 |
| from-scratch 3DCNN | cPGS Pretrained 3DCNN with cPGS | 0 | 0.011616 | from-scratch 3DCNN | cPGS Pretrained 3DCNN with cPGS | 17 | 0.51467 | from-scratch 3DCNN | cPGS Pretrained 3DCNN with cPGS | 0 | 0.001953 |
| from-scratch 3DCNN with cPGS | cPGS Pretrained 3DCNN | 5 | 0.068704 | from-scratch 3DCNN with cPGS | cPGS Pretrained 3DCNN | 15 | 0.232422 | from-scratch 3DCNN with cPGS | cPGS Pretrained 3DCNN | 4 | 0.028402 |
| from-scratch 3DCNN with cPGS | cPGS Pretrained 3DCNN with cPGS | 0 | 0.011616 | from-scratch 3DCNN with cPGS | cPGS Pretrained 3DCNN with cPGS | 11 | 0.173071 | from-scratch 3DCNN with cPGS | cPGS Pretrained 3DCNN with cPGS | 2 | 0.005859 |
| cPGS Pretrained 3DCNN | cPGS Pretrained 3DCNN with cPGS | 21.5 | 0.905613 | cPGS Pretrained 3DCNN | cPGS Pretrained 3DCNN with cPGS | 25 | 0.845703 | cPGS Pretrained 3DCNN | cPGS Pretrained 3DCNN with cPGS | 13 | 0.260393 |

**Supplementary Table 14.** Cross-sectional MDD Classification Model Comparison_Machine Learning (DEP PGS)

| ACC |  |  |  | AUROC |  |  |  | F1 |  |  |  |
| --- | --- | --- | --- | --- | --- | --- | --- | --- | --- | --- | --- |
| Model 1 | Model 2 | Statistic | P-Value | Model 1 | Model 2 | Statistic | P-Value | Model 1 | Model 2 | Statistic | P-Value |
| SVC_linear | DEP PGS Pretrained 3DCNN | 10 | 0.138641 | SVC_linear | DEP PGS Pretrained 3DCNN | 11 | 0.1054687 | SVC_linear | DEP PGS Pretrained 3DCNN | 5 | 0.019531 |
| SVC_linear | DEP PGS Pretrained 3DCNN with DEP PGS | 20 | 0.767097 | SVC_linear | DEP PGS Pretrained 3DCNN with DEP PGS | 2 | 0.015156 | SVC_linear | DEP PGS Pretrained 3DCNN with DEP PGS | 0 | 0.001953 |
| SVC_rbf | DEP PGS Pretrained 3DCNN | 24 | 0.769531 | SVC_rbf | DEP PGS Pretrained 3DCNN | 16 | 0.275391 | SVC_rbf | DEP PGS Pretrained 3DCNN | 15 | 0.232422 |
| SVC_rbf | DEP PGS Pretrained 3DCNN with DEP PGS | 18 | 0.375 | SVC_rbf | DEP PGS Pretrained 3DCNN with DEP PGS | 24.5 | 0.769531 | SVC_rbf | DEP PGS Pretrained 3DCNN with DEP PGS | 18 | 0.375 |
| XGBOOST | DEP PGS<br>Pretrained<br>3DCNN | 13 | 0.259974 | XGBOOST | DEP PGS<br>Pretrained<br>3DCNN | 16 | 0.2753906 | XGBOOST | DEP PGS<br>Pretrained<br>3DCNN | 6 | 0.027344 |
| XGBOOST | DEP PGS<br>Pretrained<br>3DCNN<br>with DEP<br>PGS | 17 | 0.888638 | XGBOOST | DEP PGS<br>Pretrained<br>3DCNN<br>with DEP<br>PGS | 19 | 0.678402 | XGBOOST | DEP PGS<br>Pretrained<br>3DCNN<br>with DEP<br>PGS | 7 | 0.037109 |
| scratch<br>3DCNN<br>with DEP<br>PGS | DEP PGS<br>Pretrained<br>3DCNN | 1 | 0.017154 | scratch<br>3DCNN<br>with DEP<br>PGS | DEP PGS<br>Pretrained<br>3DCNN | 14 | 0.193359 | scratch<br>3DCNN<br>with DEP<br>PGS | DEP PGS<br>Pretrained<br>3DCNN | 5 | 0.019531 |
| scratch<br>3DCNN<br>with DEP<br>PGS | DEP PGS<br>Pretrained<br>3DCNN<br>with DEP<br>PGS | 8 | 0.600179 | scratch<br>3DCNN<br>with DEP<br>PGS | DEP PGS<br>Pretrained<br>3DCNN<br>with DEP<br>PGS | 16 | 0.275391 | scratch<br>3DCNN<br>with DEP<br>PGS | DEP PGS<br>Pretrained<br>3DCNN<br>with DEP<br>PGS | 5 | 0.019531 |
| DEP PGS<br>Pretrained<br>3DCNN | DEP PGS<br>Pretrained<br>3DCNN<br>with DEP<br>PGS | 8 | 0.310494 | DEP PGS<br>Pretrained<br>3DCNN | DEP PGS<br>Pretrained<br>3DCNN<br>with DEP<br>PGS | 22 | 0.952765 | DEP PGS<br>Pretrained<br>3DCNN | DEP PGS<br>Pretrained<br>3DCNN<br>with DEP<br>PGS | 16 | 0.441268 |

**Supplementary Table 15.**
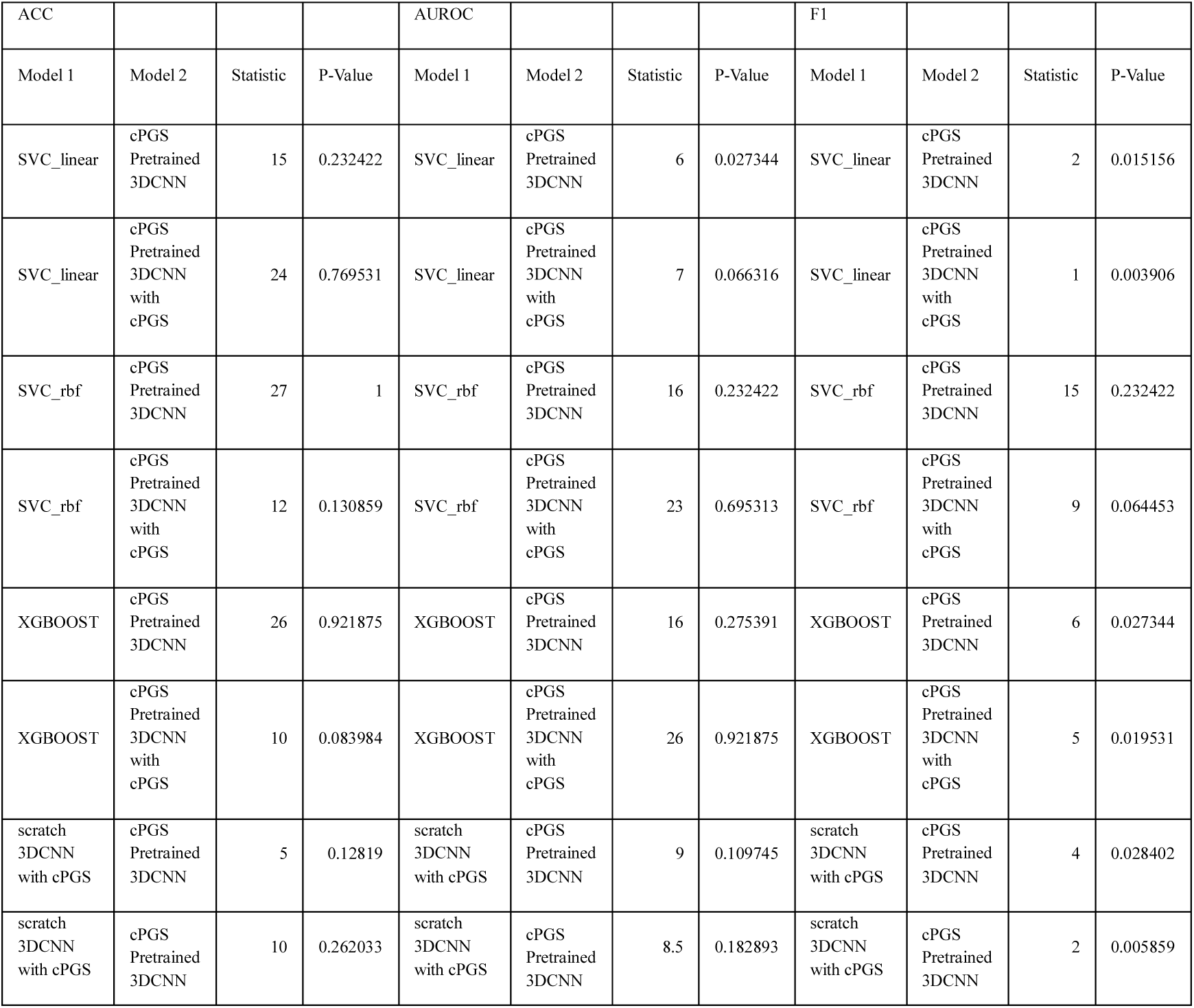
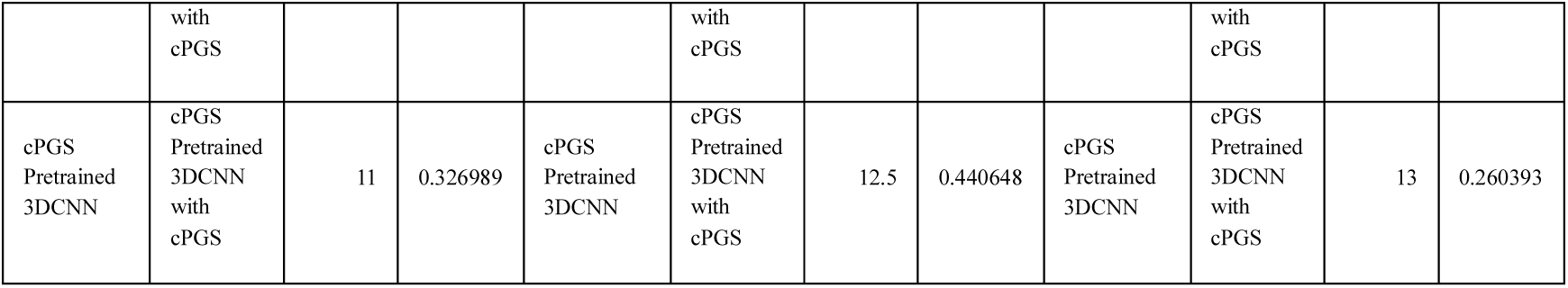
Cross-sectional MDD Classification Model Comparison_Machine Learning (cPGS)

**Supplementary Table 16.**
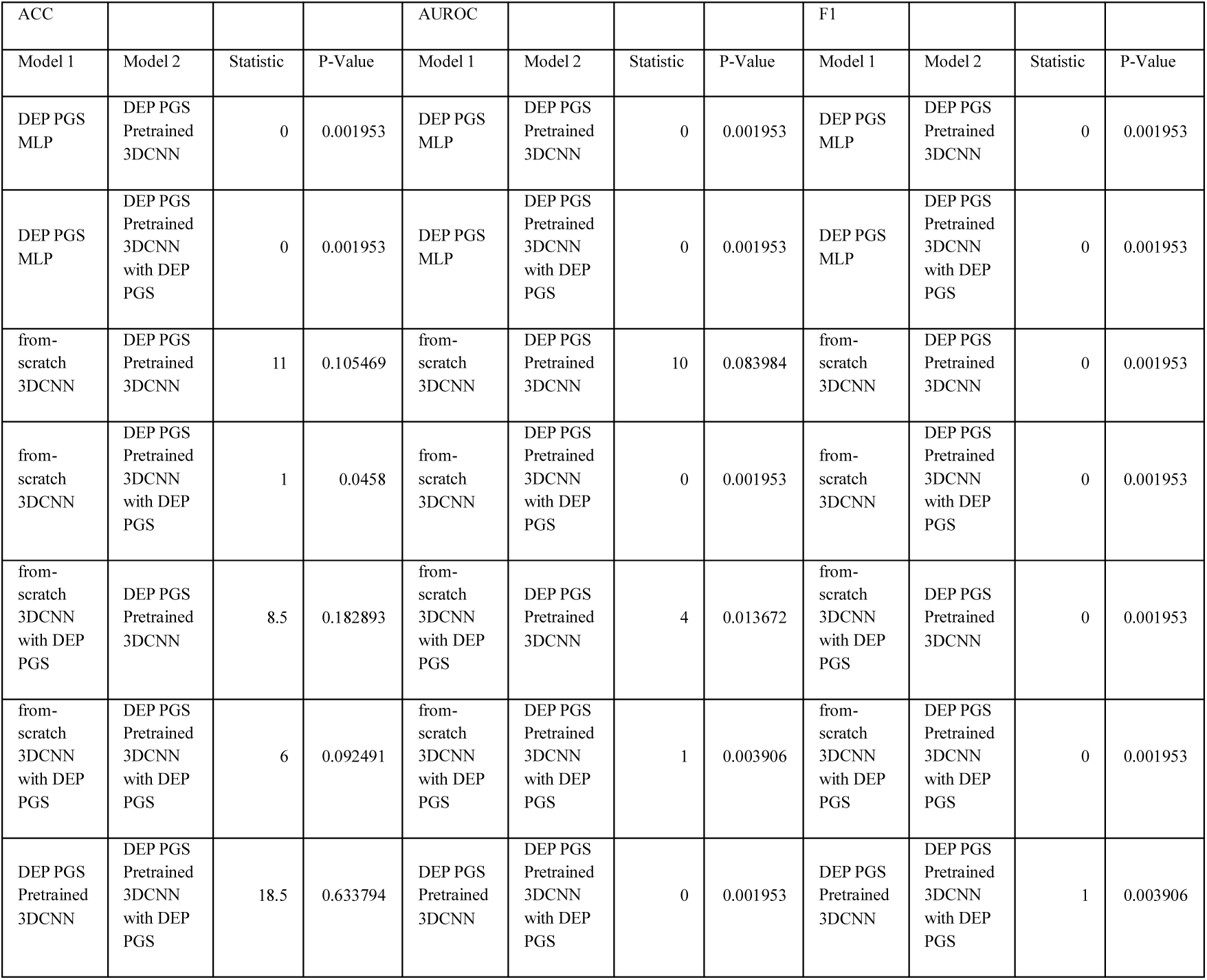
2y MDD Prediction Model Comparison_Modalities (DEP PGS)

**Supplementary Table 17.** 2y MDD Prediction Model Comparison_Modalities (cPGS)

| ACC |  |  |  | AUROC |  |  |  | F1 |  |  |  |
| --- | --- | --- | --- | --- | --- | --- | --- | --- | --- | --- | --- |
| Model 1 | Model 2 | Statistic | P-Value | Model 1 | Model 2 | Statistic | P-Value | Model 1 | Model 2 | Statistic | P-Value |
| cPGS<br>MLP | cPGS<br>Pretrained<br>3DCNN | 0 | 0.001953 | cPGS<br>MLP | cPGS<br>Pretrained<br>3DCNN | 0 | 0.001953 | cPGS<br>MLP | cPGS<br>Pretrained<br>3DCNN | 0 | 0.001953 |
| cPGS<br>MLP | cPGS<br>Pretrained<br>3DCNN<br>with cPGS | 0 | 0.001953 | cPGS<br>MLP | cPGS<br>Pretrained<br>3DCNN<br>with cPGS | 0 | 0.001953 | cPGS<br>MLP | cPGS<br>Pretrained<br>3DCNN<br>with cPGS | 0 | 0.001953 |
| from-scratch<br>3DCNN | cPGS<br>Pretrained<br>3DCNN | 3 | 0.062509 | from-scratch<br>3DCNN | cPGS<br>Pretrained<br>3DCNN | 4 | 0.013672 | from-scratch<br>3DCNN | cPGS<br>Pretrained<br>3DCNN | 0 | 0.001953 |
| from-scratch<br>3DCNN | cPGS<br>Pretrained<br>3DCNN<br>with cPGS | 2.5 | 0.017518 | from-scratch<br>3DCNN | cPGS<br>Pretrained<br>3DCNN<br>with cPGS | 0 | 0.001953 | from-scratch<br>3DCNN | cPGS<br>Pretrained<br>3DCNN<br>with cPGS | 0 | 0.001953 |
| from-scratch<br>3DCNN<br>with cPGS | cPGS<br>Pretrained<br>3DCNN | 3 | 0.020433 | from-scratch<br>3DCNN<br>with cPGS | cPGS<br>Pretrained<br>3DCNN | 24 | 0.769531 | from-scratch<br>3DCNN<br>with cPGS | cPGS<br>Pretrained<br>3DCNN | 8 | 0.048828 |
| from-scratch<br>3DCNN<br>with cPGS | cPGS<br>Pretrained<br>3DCNN<br>with cPGS | 8.5 | 0.048828 | from-scratch<br>3DCNN<br>with cPGS | cPGS<br>Pretrained<br>3DCNN<br>with cPGS | 0 | 0.001953 | from-scratch<br>3DCNN<br>with cPGS | cPGS<br>Pretrained<br>3DCNN<br>with cPGS | 0 | 0.001953 |
| cPGS<br>Pretrained<br>3DCNN | cPGS<br>Pretrained<br>3DCNN<br>with cPGS | 0 | 0.001953 | cPGS<br>Pretrained<br>3DCNN | cPGS<br>Pretrained<br>3DCNN<br>with cPGS | 0 | 0.001953 | cPGS<br>Pretrained<br>3DCNN | cPGS<br>Pretrained<br>3DCNN<br>with cPGS | 0 | 0.001953 |

**Supplementary Table 18.**
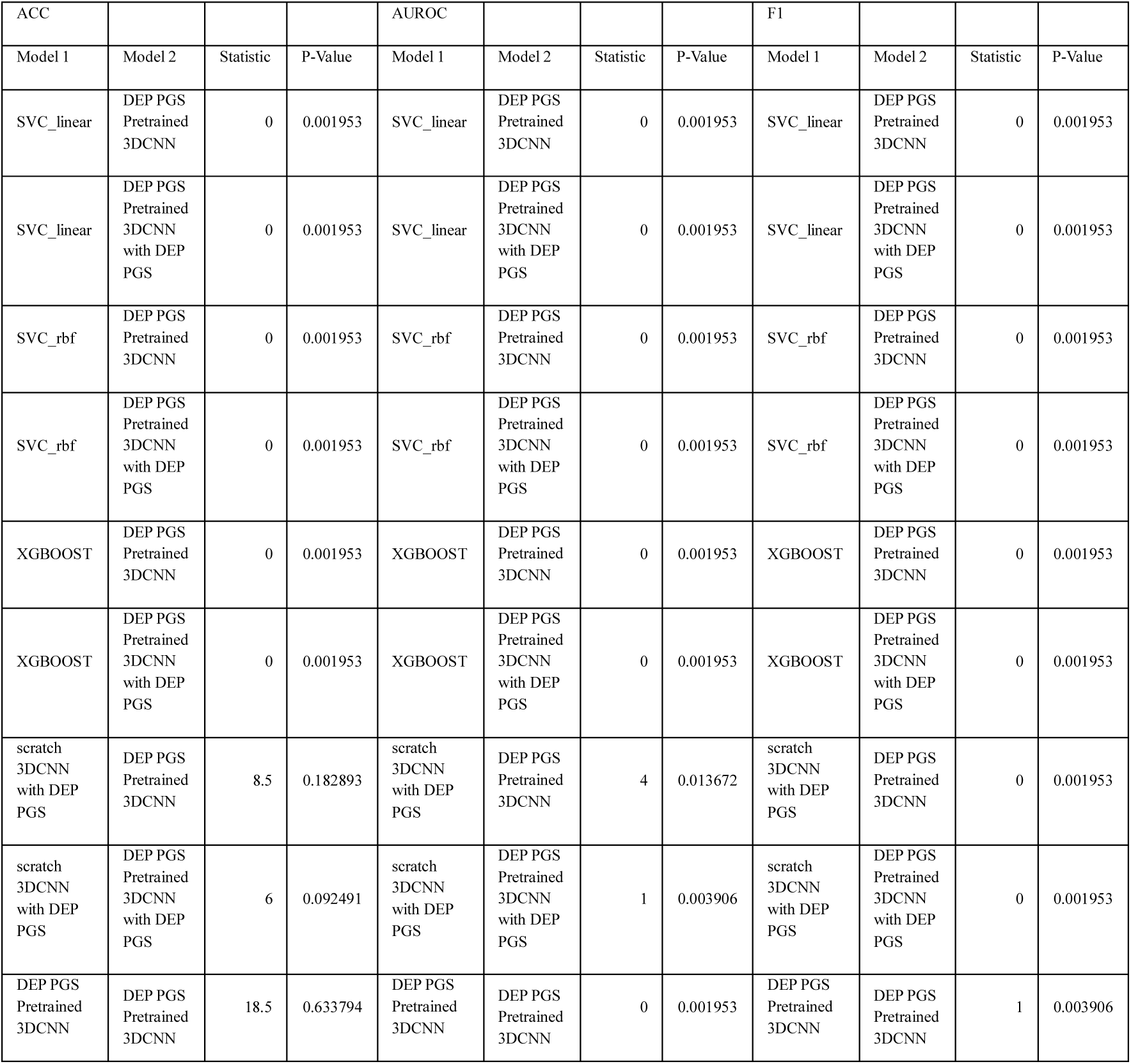
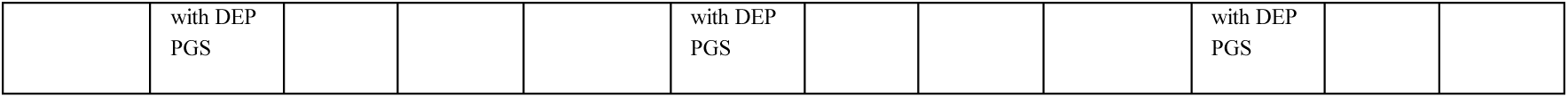
2y MDD Prediction Model Comparison_Machine Learning (DEP PGS)

**Supplementary Table 19.**
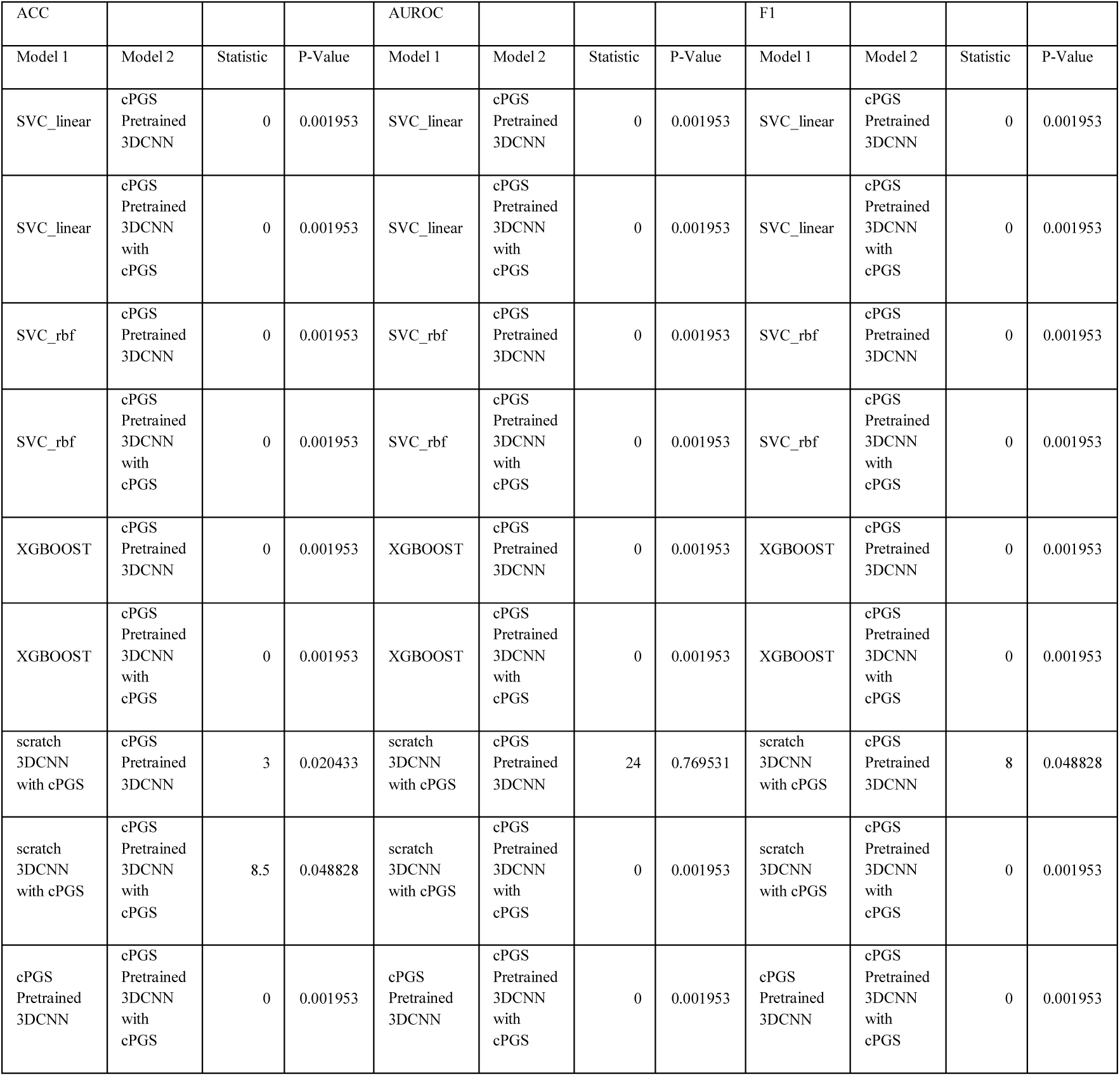
2y MDD Prediction Model Comparison_Machine Learning (cPGS)

**Supplementary Table 20.** 2y MDD + SIA Prediction Model Comparison_Modalities (DEP PGS)

| ACC |  |  |  | AUROC |  |  |  | F1 |  |  |  |
| --- | --- | --- | --- | --- | --- | --- | --- | --- | --- | --- | --- |
| Model 1 | Model 2 | Statistic | P-Value | Model 1 | Model 2 | Statistic | P-Value | Model 1 | Model 2 | Statistic | P-Value |
| DEP PGS<br>MLP | DEP PGS<br>Pretrained<br>3DCNN | 1 | 0.003906 | DEP PGS<br>MLP | DEP PGS<br>Pretrained<br>3DCNN | 1 | 0.003906 | DEP PGS<br>MLP | DEP PGS<br>Pretrained<br>3DCNN | 0 | 0.001953 |
| DEP PGS<br>MLP | DEP PGS<br>Pretrained<br>3DCNN<br>with DEP<br>PGS | 1 | 0.003906 | DEP PGS<br>MLP | DEP PGS<br>Pretrained<br>3DCNN<br>with DEP<br>PGS | 1 | 0.003906 | DEP PGS<br>MLP | DEP PGS<br>Pretrained<br>3DCNN<br>with DEP<br>PGS | 0 | 0.001953 |
| from-scratch<br>3DCNN | DEP PGS<br>Pretrained<br>3DCNN | 7.5 | 0.075304 | from-scratch<br>3DCNN | DEP PGS<br>Pretrained<br>3DCNN | 8 | 0.048828 | from-scratch<br>3DCNN | DEP PGS<br>Pretrained<br>3DCNN | 0 | 0.001953 |
| from-scratch<br>3DCNN | DEP PGS<br>Pretrained<br>3DCNN<br>with DEP<br>PGS | 0 | 0.001953 | from-scratch<br>3DCNN | DEP PGS<br>Pretrained<br>3DCNN<br>with DEP<br>PGS | 0 | 0.001953 | from-scratch<br>3DCNN | DEP PGS<br>Pretrained<br>3DCNN<br>with DEP<br>PGS | 0 | 0.001953 |
| from-scratch<br>3DCNN<br>with DEP<br>PGS | DEP PGS<br>Pretrained<br>3DCNN | 0 | 0.007474 | from-scratch<br>3DCNN<br>with DEP<br>PGS | DEP PGS<br>Pretrained<br>3DCNN | 0 | 0.001953 | from-scratch<br>3DCNN<br>with DEP<br>PGS | DEP PGS<br>Pretrained<br>3DCNN | 2 | 0.005859 |
| from-scratch<br>3DCNN<br>with DEP<br>PGS | DEP PGS<br>Pretrained<br>3DCNN<br>with DEP<br>PGS | 9 | 0.108497 | from-scratch<br>3DCNN<br>with DEP<br>PGS | DEP PGS<br>Pretrained<br>3DCNN<br>with DEP<br>PGS | 0 | 0.001953 | from-scratch<br>3DCNN<br>with DEP<br>PGS | DEP PGS<br>Pretrained<br>3DCNN<br>with DEP<br>PGS | 0 | 0.001953 |
| DEP PGS<br>Pretrained<br>3DCNN | DEP PGS<br>Pretrained<br>3DCNN<br>with DEP<br>PGS | 0 | 0.001953 | DEP PGS<br>Pretrained<br>3DCNN | DEP PGS<br>Pretrained<br>3DCNN<br>with DEP<br>PGS | 0 | 0.001953 | DEP PGS<br>Pretrained<br>3DCNN | DEP PGS<br>Pretrained<br>3DCNN<br>with DEP<br>PGS | 0 | 0.001953 |

**Supplementary Table 21.**
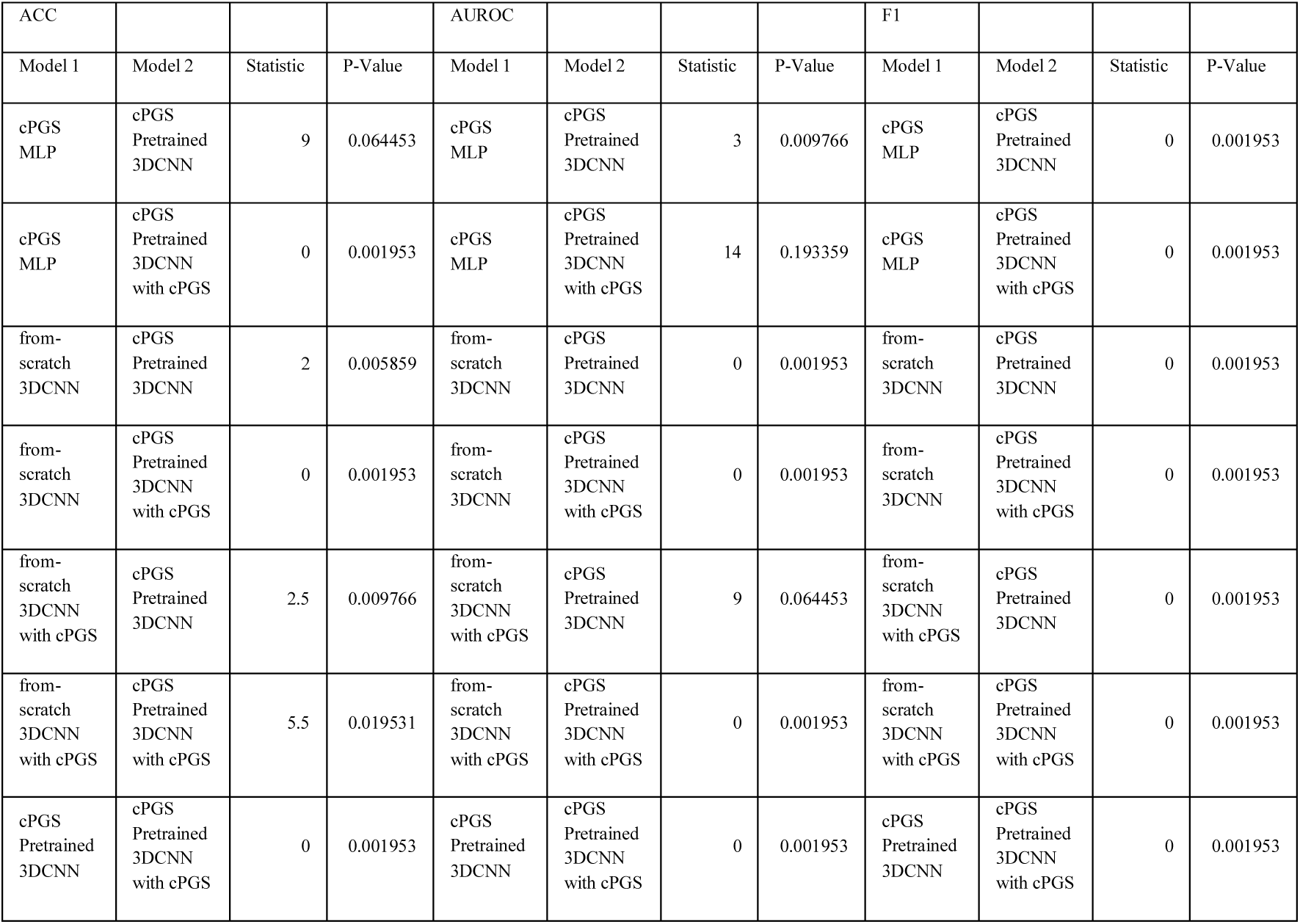
2y MDD + SIA Prediction Model Comparison_Modalities (cPGS)

| ACC |  |  |  | AUROC |  |  |  | F1 |  |  |  |
| --- | --- | --- | --- | --- | --- | --- | --- | --- | --- | --- | --- |
| Model 1 | Model 2 | Statistic | P-Value | Model 1 | Model 2 | Statistic | P-Value | Model 1 | Model 2 | Statistic | P-Value |
| cPGS MLP | cPGS Pretrained 3DCNN | 9 | 0.064453 | cPGS MLP | cPGS Pretrained 3DCNN | 3 | 0.009766 | cPGS MLP | cPGS Pretrained 3DCNN | 0 | 0.001953 |
| cPGS MLP | cPGS Pretrained 3DCNN with cPGS | 0 | 0.001953 | cPGS MLP | cPGS Pretrained 3DCNN with cPGS | 14 | 0.193359 | cPGS MLP | cPGS Pretrained 3DCNN with cPGS | 0 | 0.001953 |
| from-scratch 3DCNN | cPGS Pretrained 3DCNN | 2 | 0.005859 | from-scratch 3DCNN | cPGS Pretrained 3DCNN | 0 | 0.001953 | from-scratch 3DCNN | cPGS Pretrained 3DCNN | 0 | 0.001953 |
| from-scratch 3DCNN | cPGS Pretrained 3DCNN with cPGS | 0 | 0.001953 | from-scratch 3DCNN | cPGS Pretrained 3DCNN with cPGS | 0 | 0.001953 | from-scratch 3DCNN | cPGS Pretrained 3DCNN with cPGS | 0 | 0.001953 |
| from-scratch 3DCNN with cPGS | cPGS Pretrained 3DCNN | 2.5 | 0.009766 | from-scratch 3DCNN with cPGS | cPGS Pretrained 3DCNN | 9 | 0.064453 | from-scratch 3DCNN with cPGS | cPGS Pretrained 3DCNN | 0 | 0.001953 |
| from-scratch 3DCNN with cPGS | cPGS Pretrained 3DCNN with cPGS | 5.5 | 0.019531 | from-scratch 3DCNN with cPGS | cPGS Pretrained 3DCNN with cPGS | 0 | 0.001953 | from-scratch 3DCNN with cPGS | cPGS Pretrained 3DCNN with cPGS | 0 | 0.001953 |
| cPGS Pretrained 3DCNN | cPGS Pretrained 3DCNN with cPGS | 0 | 0.001953 | cPGS Pretrained 3DCNN | cPGS Pretrained 3DCNN with cPGS | 0 | 0.001953 | cPGS Pretrained 3DCNN | cPGS Pretrained 3DCNN with cPGS | 0 | 0.001953 |

**Supplementary Table 22.**
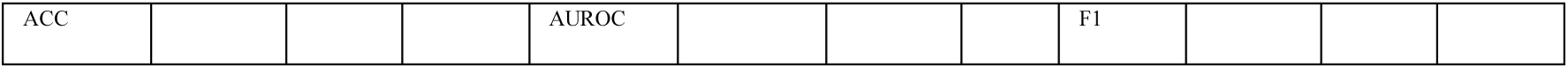
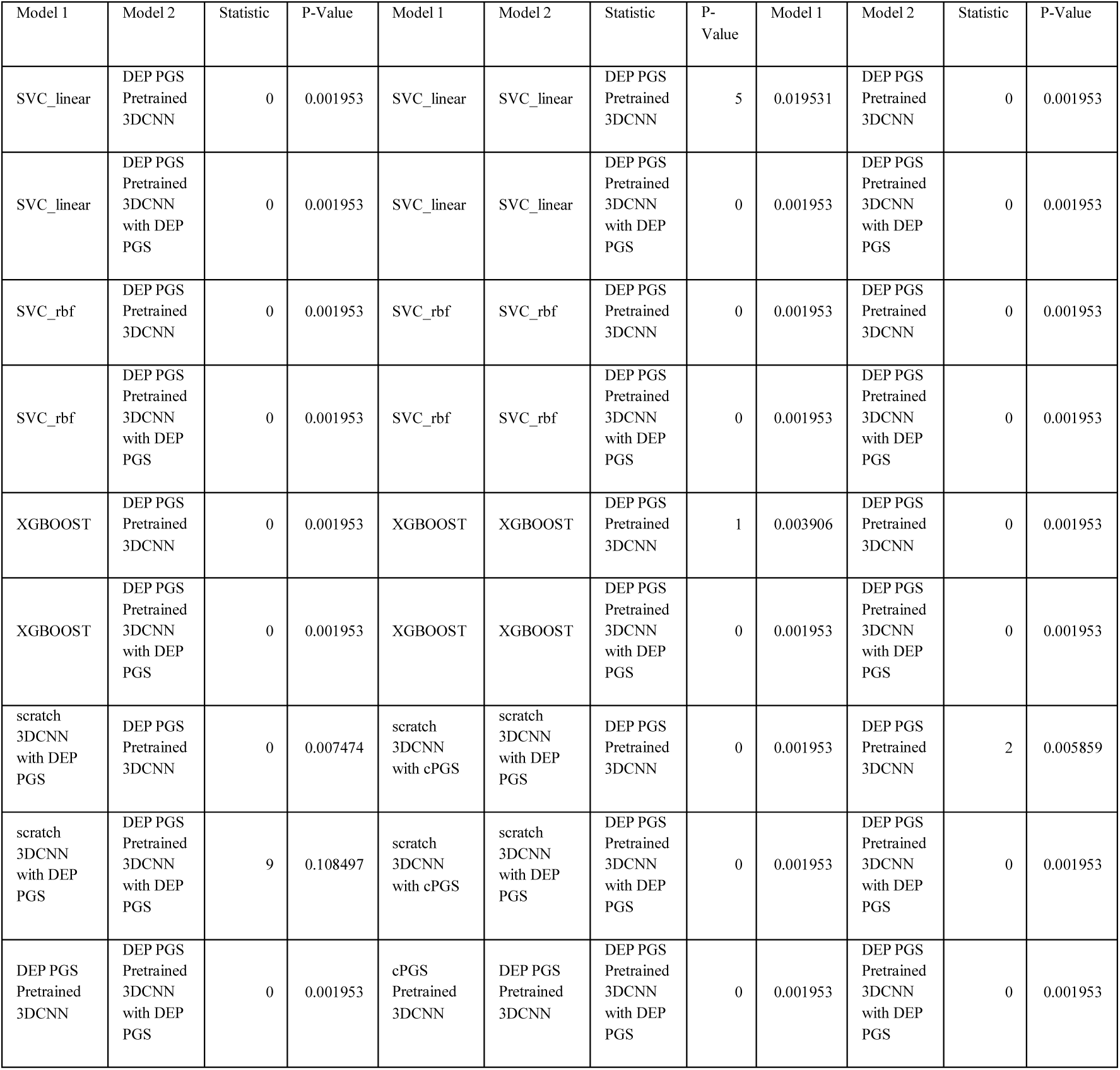
2y MDD +SIA prediction model Comparison_Machine Learning (DEP PGS)

**Supplementary Table 23.**
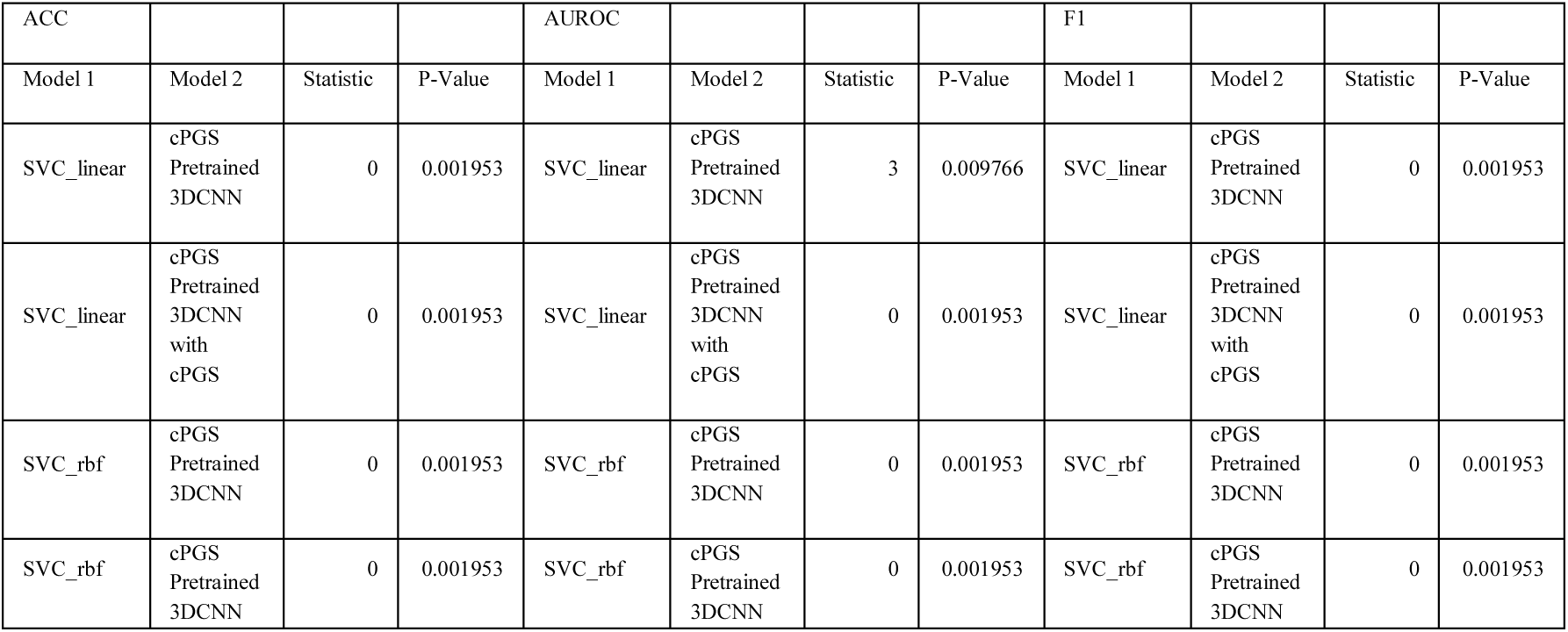
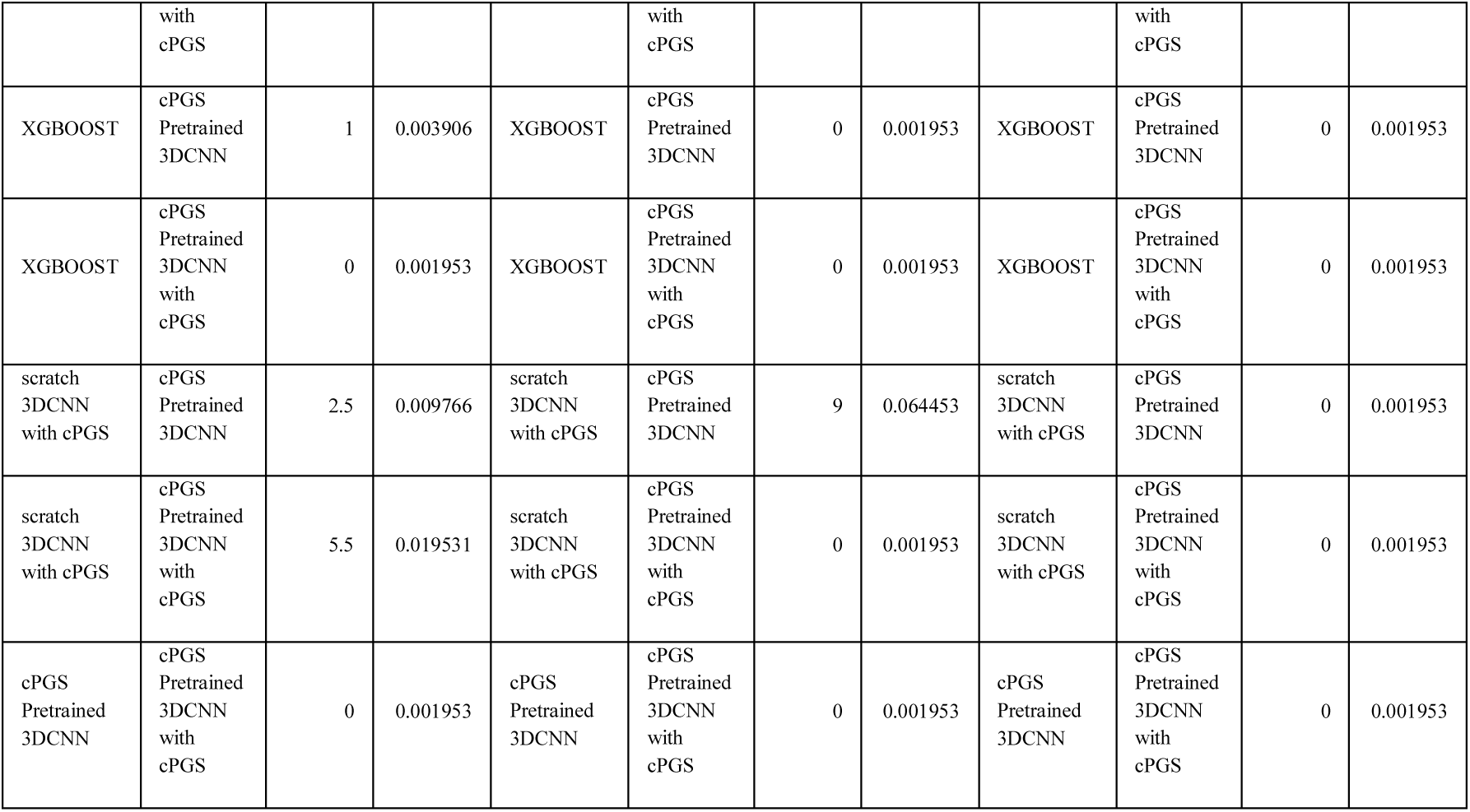
2y MDD +SIA prediction model Comparison_Machine Learning (cPGS)

**Supplementary Table 24.**
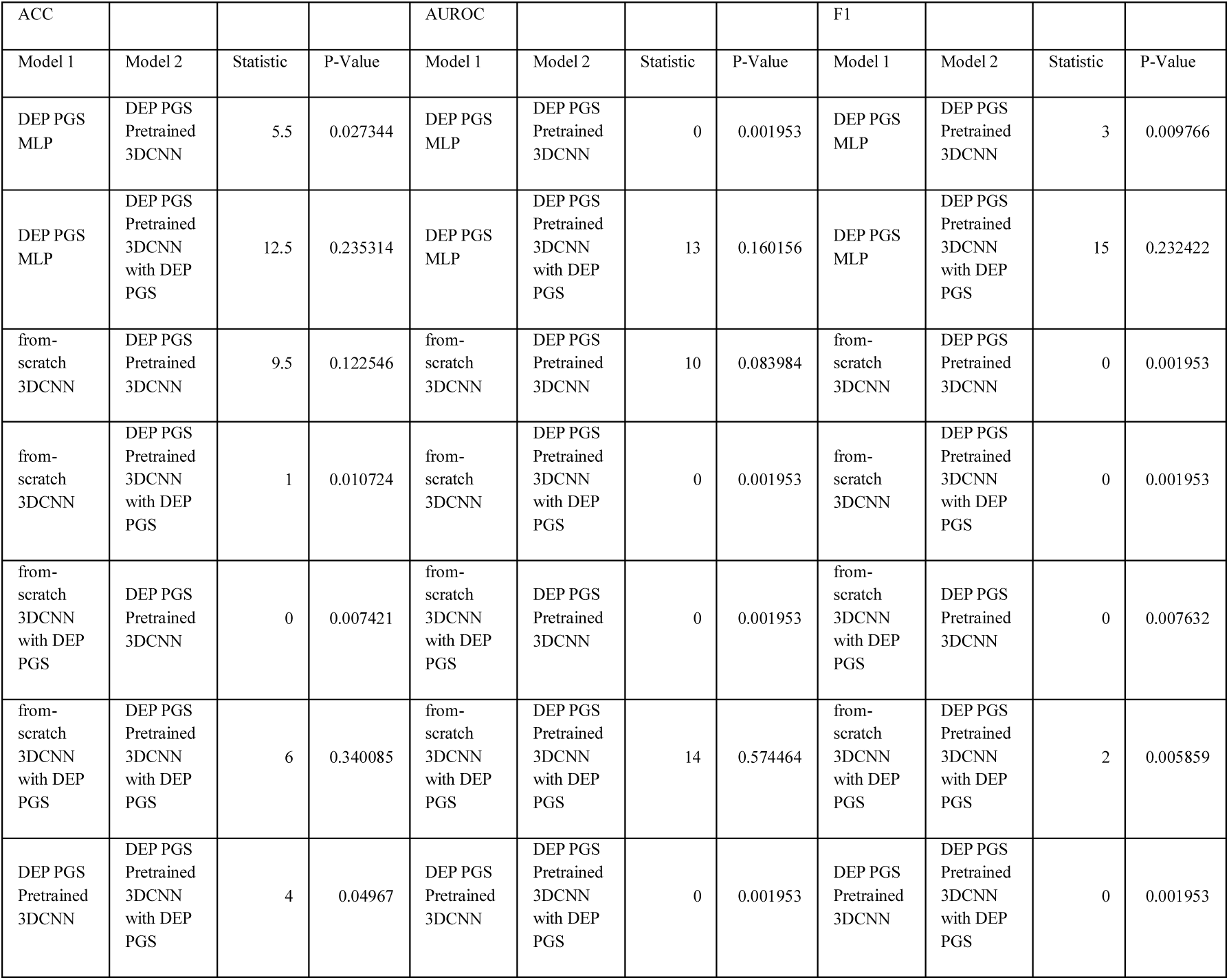
2y MDD + SA Prediction Model Comparison_Modalities (DEP PGS)

**Supplementary Table 25.** 2y MDD + SA Prediction Model Comparison_Modalities (cPGS)

| ACC |  |  |  | AUROC |  |  |  | F1 |  |  |  |
| --- | --- | --- | --- | --- | --- | --- | --- | --- | --- | --- | --- |
| Model 1 | Model 2 | Statistic | P-Value | Model 1 | Model 2 | Statistic | P-Value | Model 1 | Model 2 | Statistic | P-Value |
| cPGS MLP | cPGS Pretrained 3DCNN | 13 | 0.482765 | cPGS MLP | cPGS Pretrained 3DCNN | 7 | 0.037109 | cPGS MLP | cPGS Pretrained 3DCNN | 1 | 0.003906 |
| cPGS MLP | cPGS Pretrained 3DCNN with cPGS | 20 | 0.767097 | cPGS MLP | cPGS Pretrained 3DCNN with cPGS | 26 | 0.921875 | cPGS MLP | cPGS Pretrained 3DCNN with cPGS | 0 | 0.001953 |
| from-scratch 3DCNN | cPGS Pretrained 3DCNN | 1 | 0.010724 | from-scratch 3DCNN | cPGS Pretrained 3DCNN | 27.5 | 1 | from-scratch 3DCNN | cPGS Pretrained 3DCNN | 0 | 0.001953 |
| from-scratch 3DCNN | cPGS Pretrained 3DCNN with cPGS | 1 | 0.010586 | from-scratch 3DCNN | cPGS Pretrained 3DCNN with cPGS | 0 | 0.001953 | from-scratch 3DCNN | cPGS Pretrained 3DCNN with cPGS | 0 | 0.001953 |
| from-scratch 3DCNN with cPGS | cPGS Pretrained 3DCNN | 13.5 | 0.526066 | from-scratch 3DCNN with cPGS | cPGS Pretrained 3DCNN | 19 | 0.431641 | from-scratch 3DCNN with cPGS | cPGS Pretrained 3DCNN | 9.5 | 0.123205 |
| from-scratch 3DCNN with cPGS | cPGS Pretrained 3DCNN with cPGS | 6.5 | 0.203272 | from-scratch 3DCNN with cPGS | cPGS Pretrained 3DCNN with cPGS | 5 | 0.019531 | from-scratch 3DCNN with cPGS | cPGS Pretrained 3DCNN with cPGS | 0 | 0.007686 |
| cPGS Pretrained 3DCNN | cPGS Pretrained 3DCNN with cPGS | 3 | 0.113846 | cPGS Pretrained 3DCNN | cPGS Pretrained 3DCNN with cPGS | 0 | 0.001953 | cPGS Pretrained 3DCNN | cPGS Pretrained 3DCNN with cPGS | 0 | 0.001953 |

**Supplementary Table 26.**
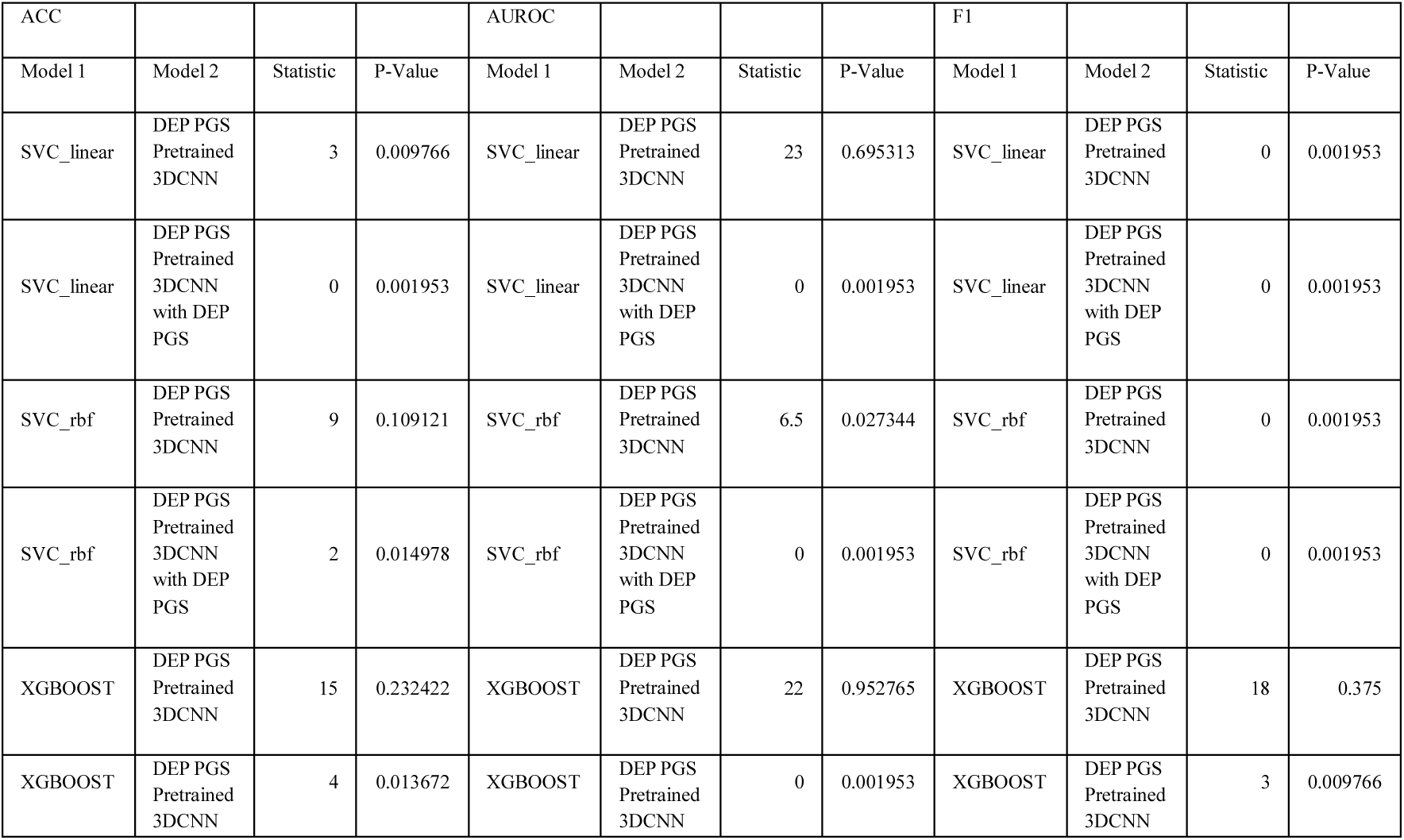
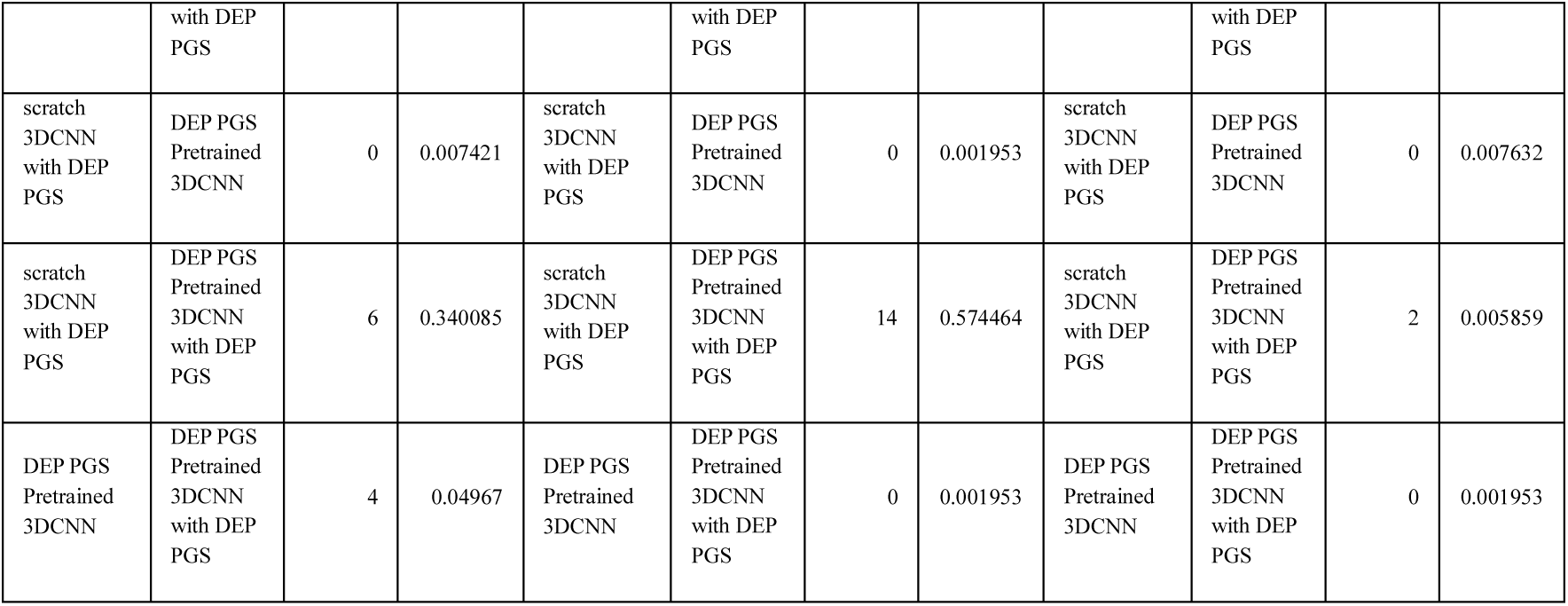
2y MDD +SA Prediction Model Comparison_Machine Learning (DEP PGS)

**Supplementary Table 27.** 2y MDD +SA Prediction Model Comparison_Machine Learning (cPGS)

| ACC |  |  |  | AUROC |  |  |  | F1 |  |  |  |
| --- | --- | --- | --- | --- | --- | --- | --- | --- | --- | --- | --- |
| Model 1 | Model 2 | Statistic | P-Value | Model 1 | Model 2 | Statistic | P-Value | Model 1 | Model 2 | Statistic | P-Value |
| SVC_linear | cPGS Pretrained 3DCNN | 0 | 0.007579 | SVC_linear | cPGS Pretrained 3DCNN | 13 | 0.160156 | SVC_linear | cPGS Pretrained 3DCNN | 0 | 0.001953 |
| SVC_linear | cPGS Pretrained 3DCNN with cPGS | 0 | 0.007632 | SVC_linear | cPGS Pretrained 3DCNN with cPGS | 0 | 0.001953 | SVC_linear | cPGS Pretrained 3DCNN with cPGS | 0 | 0.001953 |
| SVC_rbf | cPGS Pretrained 3DCNN | 1 | 0.003906 | SVC_rbf | cPGS Pretrained 3DCNN | 0 | 0.001953 | SVC_rbf | cPGS Pretrained 3DCNN | 0 | 0.001953 |
| SVC_rbf | cPGS Pretrained 3DCNN with cPGS | 1 | 0.003906 | SVC_rbf | cPGS Pretrained 3DCNN with cPGS | 0 | 0.001953 | SVC_rbf | cPGS Pretrained 3DCNN with cPGS | 0 | 0.001953 |
| XGBOOST | cPGS Pretrained 3DCNN | 9 | 0.064453 | XGBOOST | cPGS Pretrained 3DCNN | 13 | 0.160156 | XGBOOST | cPGS Pretrained 3DCNN | 2 | 0.005859 |
| XGBOOST | cPGS Pretrained 3DCNN with cPGS | 9 | 0.064453 | XGBOOST | cPGS Pretrained 3DCNN with cPGS | 2 | 0.005859 | XGBOOST | cPGS Pretrained 3DCNN with cPGS | 1 | 0.003906 |
| scratch 3DCNN with cPGS | cPGS Pretrained 3DCNN | 13.5 | 0.526066 | scratch 3DCNN with cPGS | cPGS Pretrained 3DCNN | 19 | 0.431641 | scratch 3DCNN with cPGS | cPGS Pretrained 3DCNN | 9.5 | 0.123205 |
| scratch 3DCNN with cPGS | cPGS Pretrained 3DCNN with cPGS | 6.5 | 0.203272 | scratch 3DCNN with cPGS | cPGS Pretrained 3DCNN with cPGS | 5 | 0.019531 | scratch 3DCNN with cPGS | cPGS Pretrained 3DCNN with cPGS | 0 | 0.007686 |
| cPGS Pretrained 3DCNN | cPGS Pretrained 3DCNN with cPGS | 3 | 0.113846 | cPGS Pretrained 3DCNN | cPGS Pretrained 3DCNN with cPGS | 0 | 0.001953 | cPGS Pretrained 3DCNN | cPGS Pretrained 3DCNN with cPGS | 0 | 0.001953 |

**Supplementary Table 28.**
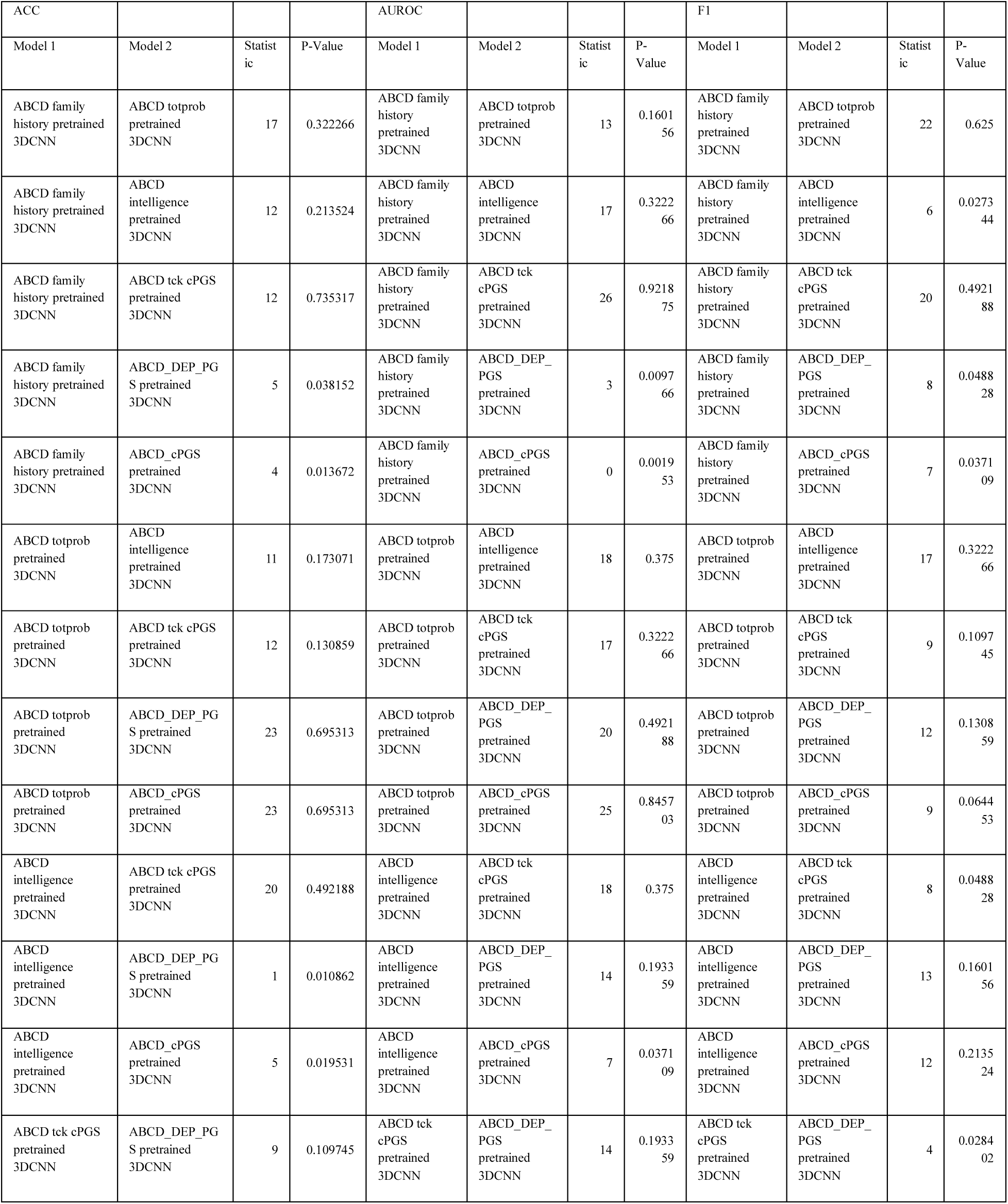
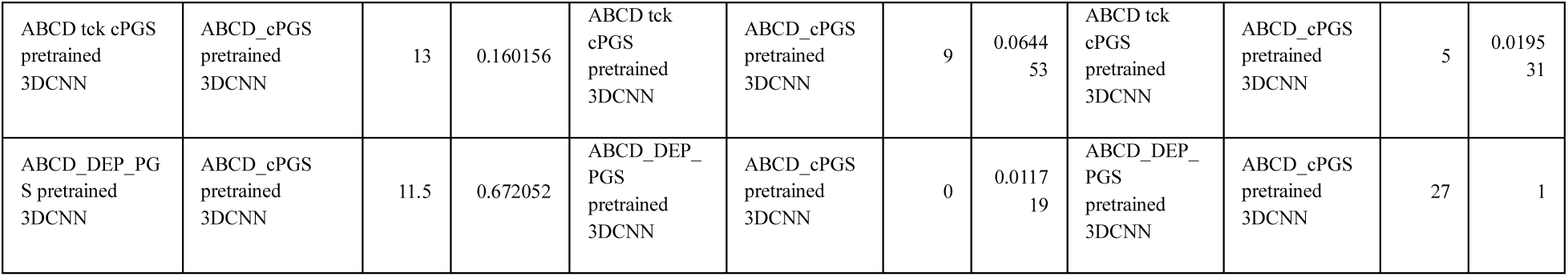
Control Analysis Performance Comparison.

**Supplementary Table 29.** 2y MDD without Suicidal Behavior Prediction Model Comparison_Modalities (DEP PGS)

| ACC |  |  |  | AUROC |  |  |  | F1 |  |  |  |
| --- | --- | --- | --- | --- | --- | --- | --- | --- | --- | --- | --- |
| Model 1 | Model 2 | Statistic | P-Value | Model 1 | Model 2 | Statistic | P-Value | Model 1 | Model 2 | Statistic | P-Value |
| DEP PGS<br>MLP | DEP PGS<br>Pretrained<br>3DCNN | 0 | 0.001953 | DEP PGS<br>MLP | DEP PGS<br>Pretrained<br>3DCNN | 0 | 0.001953 | DEP PGS<br>MLP | DEP PGS<br>Pretrained<br>3DCNN | 0 | 0.001953 |
| DEP PGS<br>MLP | DEP PGS<br>Pretrained<br>3DCNN<br>with DEP<br>PGS | 0 | 0.001953 | DEP PGS<br>MLP | DEP PGS<br>Pretrained<br>3DCNN<br>with DEP<br>PGS | 0 | 0.001953 | DEP PGS<br>MLP | DEP PGS<br>Pretrained<br>3DCNN<br>with DEP<br>PGS | 0 | 0.001953 |
| from-scratch<br>3DCNN | DEP PGS<br>Pretrained<br>3DCNN | 3 | 0.035015 | from-scratch<br>3DCNN | DEP PGS<br>Pretrained<br>3DCNN | 22.5 | 0.625 | from-scratch<br>3DCNN | DEP PGS<br>Pretrained<br>3DCNN | 0 | 0.001953 |
| from-scratch<br>3DCNN | DEP PGS<br>Pretrained<br>3DCNN<br>with DEP<br>PGS | 1 | 0.003906 | from-scratch<br>3DCNN | DEP PGS<br>Pretrained<br>3DCNN<br>with DEP<br>PGS | 16 | 0.441268 | from-scratch<br>3DCNN | DEP PGS<br>Pretrained<br>3DCNN<br>with DEP<br>PGS | 0 | 0.001953 |
| from-scratch<br>3DCNN<br>with DEP<br>PGS | DEP PGS<br>Pretrained<br>3DCNN | 0 | 0.027281 | from-scratch<br>3DCNN<br>with DEP<br>PGS | DEP PGS<br>Pretrained<br>3DCNN | 20 | 0.492188 | from-scratch<br>3DCNN<br>with DEP<br>PGS | DEP PGS<br>Pretrained<br>3DCNN | 0 | 0.001953 |
| from-scratch<br>3DCNN<br>with DEP<br>PGS | DEP PGS<br>Pretrained<br>3DCNN<br>with DEP<br>PGS | 1 | 0.003906 | from-scratch<br>3DCNN<br>with DEP<br>PGS | DEP PGS<br>Pretrained<br>3DCNN<br>with DEP<br>PGS | 7 | 0.037109 | from-scratch<br>3DCNN<br>with DEP<br>PGS | DEP PGS<br>Pretrained<br>3DCNN<br>with DEP<br>PGS | 0 | 0.001953 |
| DEP PGS<br>Pretrained<br>3DCNN | DEP PGS<br>Pretrained<br>3DCNN<br>with DEP<br>PGS | 1 | 0.003906 | DEP PGS<br>Pretrained<br>3DCNN | DEP PGS<br>Pretrained<br>3DCNN<br>with DEP<br>PGS | 18 | 0.375 | DEP PGS<br>Pretrained<br>3DCNN | DEP PGS<br>Pretrained<br>3DCNN<br>with DEP<br>PGS | 1 | 0.003906 |

**Supplementary Table 30.**
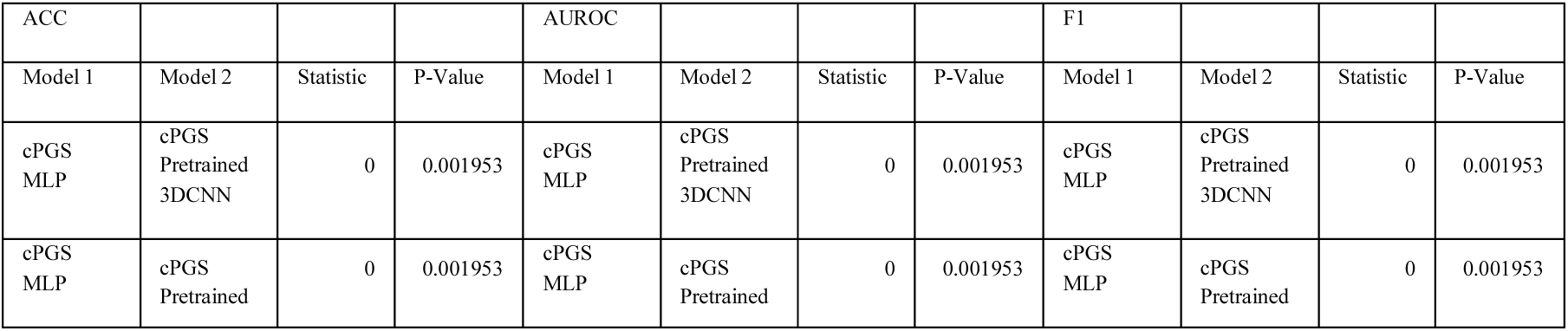
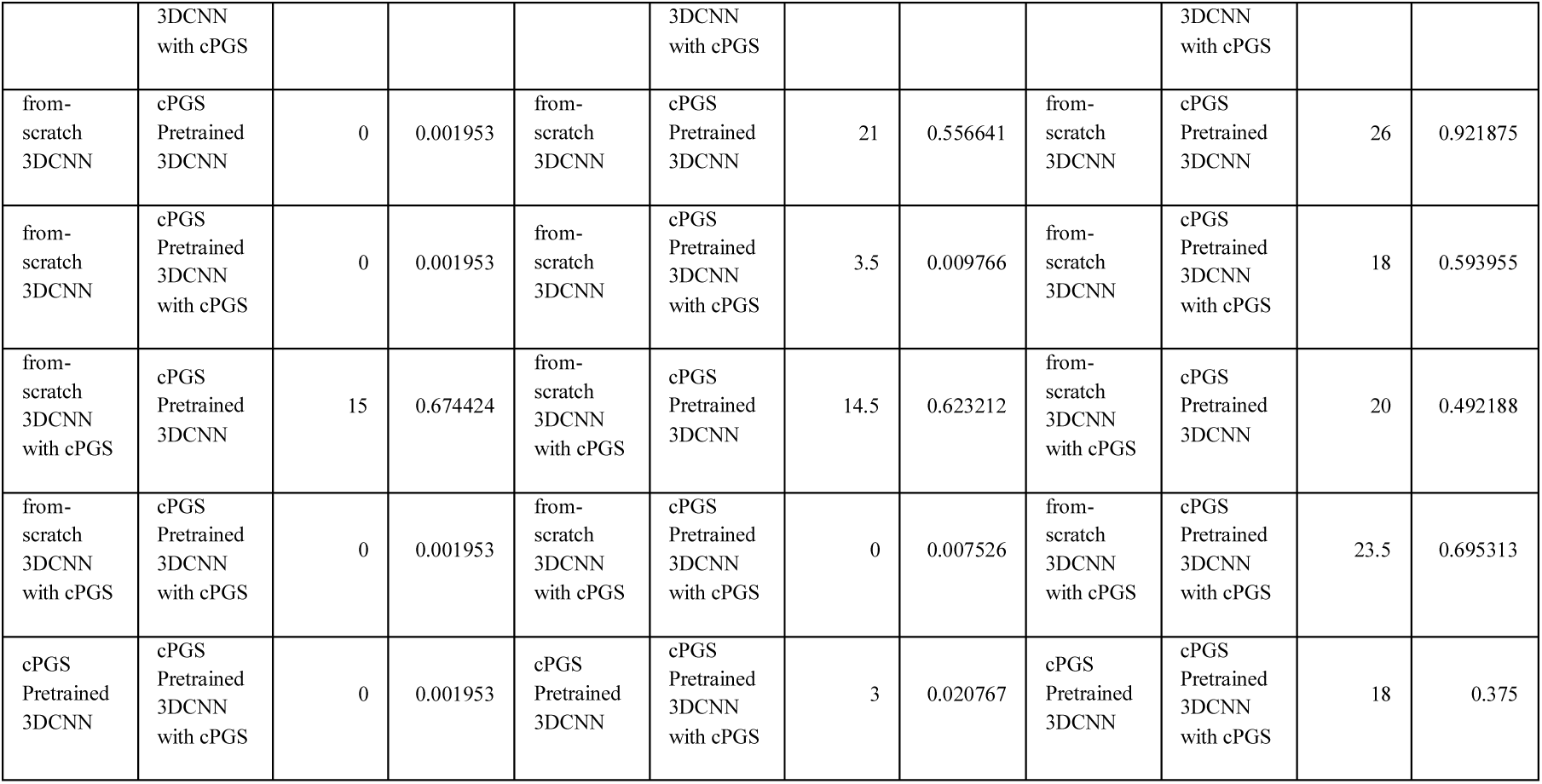
2y MDD without Suicidal Behavior Prediction Model Comparison_Modalities (cPGS)

**Supplementary Table 31.**
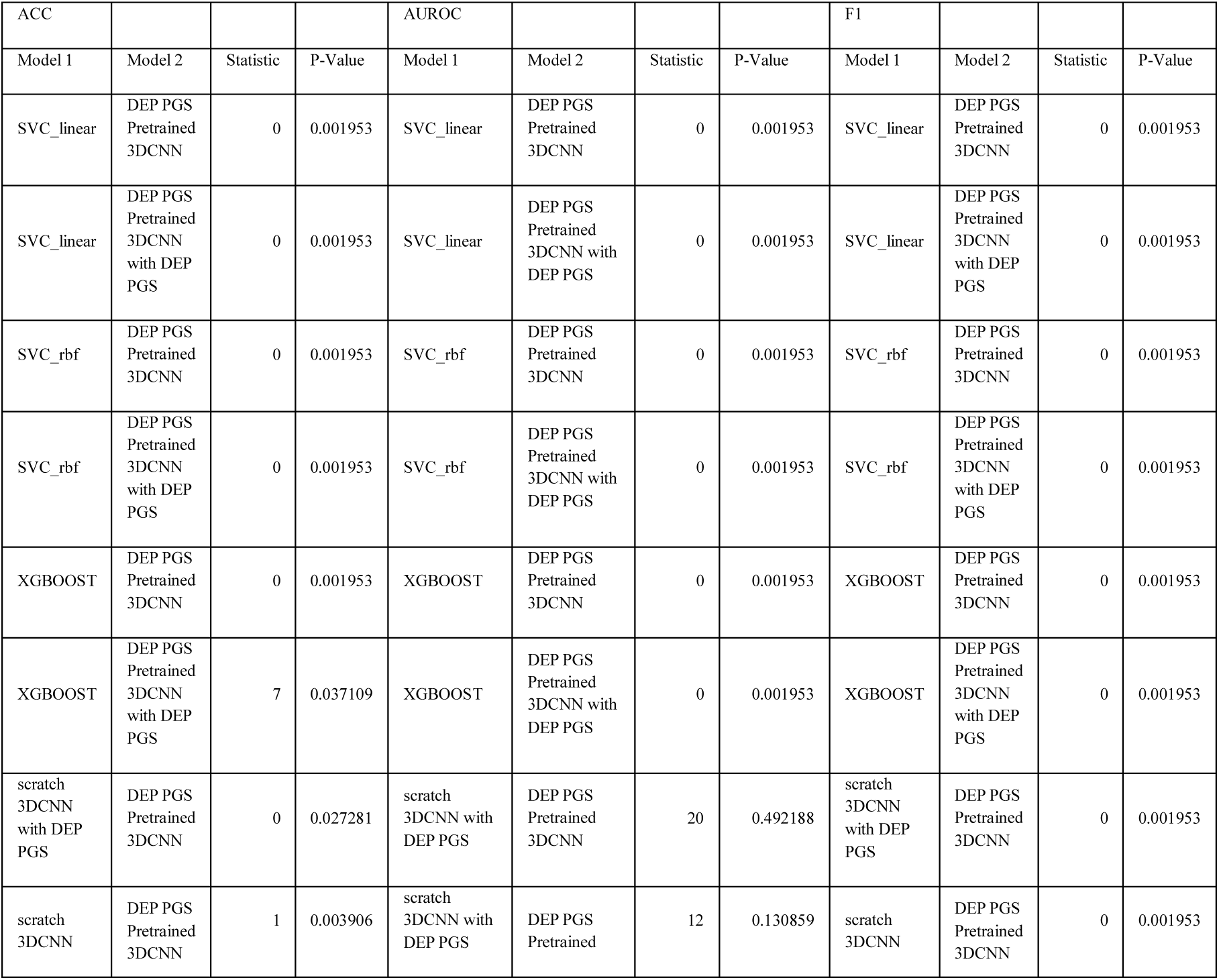
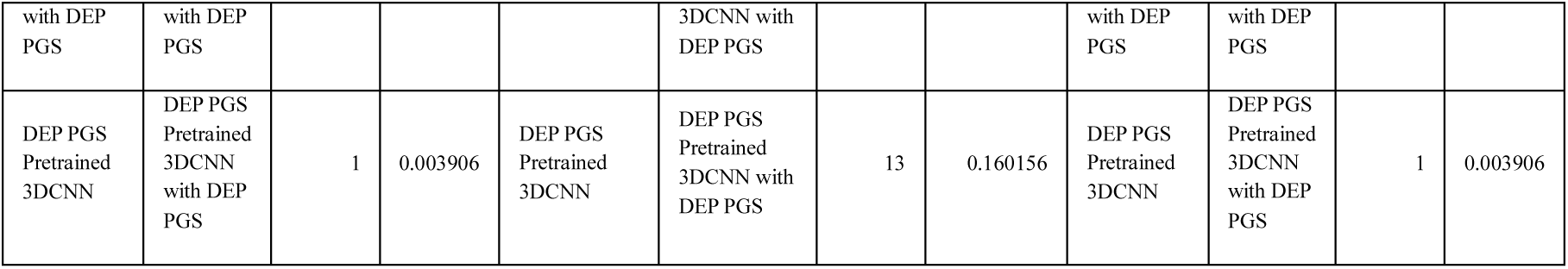
2y MDD without Suicidal Behavior Prediction Model Comparison_Machine Learning (DEP PGS)

**Supplementary Table 32.** 2y MDD without Suicidal Behavior Prediction Model Comparison_Machine Learning (cPGS)

| ACC |  |  |  | AUROC |  |  |  | F1 |  |  |  |
| --- | --- | --- | --- | --- | --- | --- | --- | --- | --- | --- | --- |
| Model 1 | Model 2 | Statistic | P-Value | Model 1 | Model 2 | Statistic | P-Value | Model 1 | Model 2 | Statistic | P-Value |
| SVC_linear | cPGS Pretrained 3DCNN | 0 | 0.001953 | SVC_linear | cPGS Pretrained 3DCNN | 0 | 0.001953 | SVC_linear | cPGS Pretrained 3DCNN | 0 | 0.001953 |
| SVC_linear | cPGS Pretrained 3DCNN with cPGS | 0 | 0.001953 | SVC_linear | cPGS Pretrained 3DCNN with cPGS | 0 | 0.001953 | SVC_linear | cPGS Pretrained 3DCNN with cPGS | 0 | 0.001953 |
| SVC_rbf | cPGS Pretrained 3DCNN | 0 | 0.001953 | SVC_rbf | cPGS Pretrained 3DCNN | 1 | 0.003906 | SVC_rbf | cPGS Pretrained 3DCNN | 0 | 0.001953 |
| SVC_rbf | cPGS Pretrained 3DCNN with cPGS | 0 | 0.001953 | SVC_rbf | cPGS Pretrained 3DCNN with cPGS | 1 | 0.003906 | SVC_rbf | cPGS Pretrained 3DCNN with cPGS | 0 | 0.001953 |
| XGBOOST | cPGS Pretrained 3DCNN | 1 | 0.003906 | XGBOOST | cPGS Pretrained 3DCNN | 0 | 0.001953 | XGBOOST | cPGS Pretrained 3DCNN | 0 | 0.001953 |
| XGBOOST | cPGS Pretrained 3DCNN with cPGS | 0 | 0.001953 | XGBOOST | cPGS Pretrained 3DCNN with cPGS | 0 | 0.001953 | XGBOOST | cPGS Pretrained 3DCNN with cPGS | 0 | 0.001953 |
| scratch 3DCNN with cPGS | cPGS Pretrained 3DCNN | 14.5 | 0.623212 | scratch 3DCNN with cPGS | cPGS Pretrained 3DCNN | 20 | 0.492188 | scratch 3DCNN with cPGS | cPGS Pretrained 3DCNN | 15 | 0.674424 |
| scratch 3DCNN with cPGS | cPGS Pretrained 3DCNN with cPGS | 0 | 0.007526 | scratch 3DCNN with cPGS | cPGS Pretrained 3DCNN with cPGS | 23.5 | 0.695313 | scratch 3DCNN with cPGS | cPGS Pretrained 3DCNN with cPGS | 0 | 0.001953 |
| cPGS Pretrained 3DCNN | cPGS Pretrained 3DCNN with cPGS | 3 | 0.020767 | cPGS Pretrained 3DCNN | cPGS Pretrained 3DCNN with cPGS | 18 | 0.375 | cPGS Pretrained 3DCNN | cPGS Pretrained 3DCNN with cPGS | 0 | 0.001953 |

**Supplementary Table 33.**
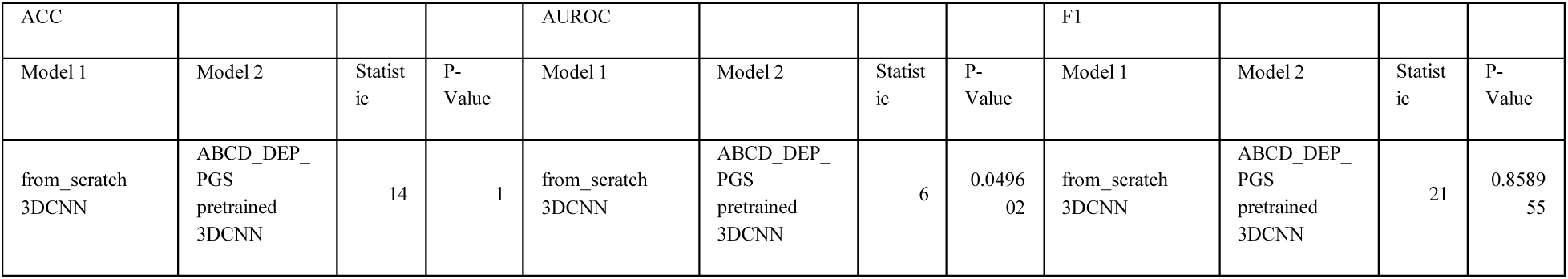
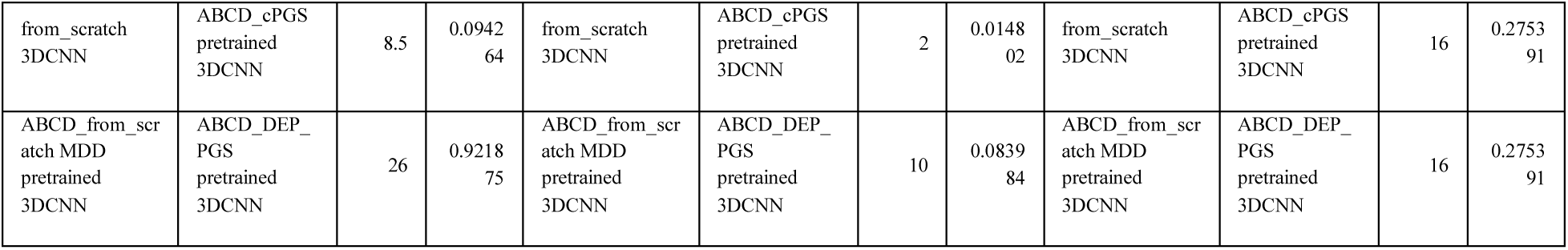
Korean dataset Depression Classification model Comparison.

